# Combination of single cell sequencing data and GWAS summary statistics reveals genetically-influenced liver cell types for primary biliary cholangitis

**DOI:** 10.1101/2021.08.18.21262250

**Authors:** Bingyu Xiang, Chunyu Deng, Jingjing Li, Shanshan Li, Huifang Zhang, Xiuli Lin, Mingqin Lu, Yunlong Ma

**Affiliations:** Department of Infectious Diseases, the First Affiliated Hospital of Wenzhou Medical University, Wenzhou 325000, Zhejiang, China; Institute of Biomedical Big Data, School of Ophthalmology & Optometry and Eye Hospital, School of Biomedical Engineering, Wenzhou Medical University, Wenzhou 325027, Zhejiang, China; State Key Laboratory for Diagnosis and Treatment of Infectious Diseases, The First Affiliated Hospital, Collaborative Innovation Center for Diagnosis and Treatment of Infectious Diseases, Zhejiang University School of Medicine, Hangzhou 310003, Zhejiang, China

## Abstract

**Importance:** Primary biliary cholangitis (PBC) is a classical autoimmune disease, which is highly influenced by genetic determinants. Many genome-wide association studies (GWAS) have reported that numerous genetic loci were significantly associated with PBC susceptibility. However, the effects of genetic determinants on liver cells and its immune microenvironment for PBC remain unclear.

**Objective:** To identify genetics-modulated functional liver cell subsets involved in the pathogenesis of PBC.

**Design, Setting, and Participants:** In this present study, 13,239 European participants were collected from IEU open GWAS project on PBC. There were 1,124,241 qualified SNPs used for GWAS analysis. Expression quantitative trait loci (eQTL) data across 49 tissues were downloaded from the GTEx database. Two single cell RNA sequencing (scRNA-seq) profiles and two bulk-based RNA transcriptomes were downloaded from the GEO database. Data collection and analyses were performed from August 2020 to June 2021.

**Main outcomes and measures:** We constructed a powerful computational framework to integrate GWAS summary statistics with scRNA-seq data to uncover genetics-modulated liver cell subpopulations.

**Results:** Based on our multi-omics integrative analysis, we found that 29 risk genes including *ORMDL3, GSNK2B*, and *DDAH2* were significantly associated with PBC susceptibility. Gene-property analysis revealed that four immune cell types, including Cst3^+^ dendritic cell, Chil3^+^ macrophage, Trbc2^+^ T cell, and Gzma^+^ T cell, were significantly enriched by PBC-risk genes. By combining GWAS summary statistics with scRNA-seq data, we found that cholangiocytes exhibited a notable enrichment by PBC-related genetic association signals (Permuted P < 0.05). The risk gene of *ORMDL3* showed the highest expression proportion in cholangiocytes than other liver cells (22.38%). Compared with *ORMDL3*^+^ cholangiocytes, there were 71 significantly highly-expressed genes among *ORMDL3*^-^ cholangiocytes (FDR < 0.05), such as inflammatory cytokine genes *CXCL8, CCL3, IFI16*, and *IRF1*. These highly-expressed genes were significantly enriched in numerous biological pathways and functional terms associated with autoimmune diseases (FDR < 0.05).

**Conclusions and relevance:** To the best of our knowledge, this is the first study to integrate genetic information with single cell sequencing data for parsing genetics-influenced liver cells for PBC risk. We identified that *ORMDL3*^-^ cholangiocytes play important immune-modulatory roles in the etiology of PBC.

**Key points:** *Question:* Are genetics factors influenced liver cell subpopulations and its immune microenvironment for PBC?

*Findings:* In this comprehensive genomics study based on multi-omics data, genetic determinants were significantly enriched in cholangiocytes and immune cells including subsets of macrophage, dendritic cells, and T cells. *ORMDL3*^-^ cholangiocytes have crucial immune-modulatory roles in developing PBC.

*Meaning:* Findings suggest that integration of single cell sequencing data with GWAS summary statistics contribute to pinpoint PBC-relevant cell types and risk genes.

## Introduction

Primary biliary cholangitis (PBC), which is formally known as primary biliary cirrhosis until 2016 ^1^, is a rare chronic cholestatic liver disease characterized by progressive autoimmune-mediated destruction of the small intrahepatic biliary epithelial cells ^2,3^. PBC patients suffering from chronic cholestasis can eventually lead to cirrhosis and hepatic failure without effective treatments ^2^. Although ursodeoxycholic acid has been used as the first-line therapeutic agent for PBC, there exist 10% to 20% of PBC patients resistant to ursodeoxycholic acid and developing to advanced-stage liver disease ^2^. Previous studies ^4^ have reported that a combination of genetic and environmental risk factors have an important influence on the aetiology of PBC. Hence, understanding the genetic mechanisms of PBC is becoming a great interest, which may promote the development of individualized therapeutic strategy for PBC.

Over the past decade, a growing number of genome-wide association studies (GWAS) and Immunochip studies based on East Asian and European populations have been performed to uncover the genetic susceptibility loci associated with PBC ^4^. To date, more than 40 genetic loci with numerous risk genes have been reported ^5-10^, such as *SLC19A3/CCL20, IRF8/FOXF1, NFKB1/MANBA*, and *PDGFB/RPL3*. Nevertheless, the GWAS approach has generally focused on examining the genetic associations of millions of single nucleotide polymorphisms (SNPs) and only a handful of SNPs with a genome-wide significance (P ≤ 5×10^−8^) are reported. There exist many common SNPs with small marginal effects were neglected ^11,12^. Moreover, the vast majority of reported SNPs were mapped within non-coding genomic regions^12^. It is plausible to infer that these non-coding SNPs may modulate the expression levels of corresponding risk genes rather than change the functions of their proteins. Thus, combination of GWAS summary statistics and other different types of data that characterize tissue- and cell-type-specific activity, including expression quantitative trait loci ^11^, DNase I-hypersensitive sites (DHS) ^13^, and histone marks ^14^, contributes to highlight disease-related risk genes and cell types.

With the advance of single cell sequencing techniques, researchers have an effective avenue to discover more refined and novel cell populations for complex diseases ^15^. An accruing and large number of single cell RNA sequencing (scRNA-seq) studies on autoimmune diseases, including rheumatoid arthritis ^16^, inflammatory bowel disease ^17^, and systemic lupus erythematosus ^18^, have been reported to parse the heterogeneity of cellular subpopulations at unprecedented resolution. In view of no scRNA-seq study was conducted for uncovering human liver cell types implicated in PBC, we constructed a computational framework to identify risk genes whose genetically expressions associated with PBC and pinpoint cell subpopulations implicated in the etiology of PBC.

## Methods

### Datasets

#### 1. Single-cell transcriptomes of PBC

We downloaded two independent single cell RNA sequencing profiles (Accession number: GSE93170 and GSE115469) from the GEO database. With regard to the dataset of GSE93170, there were clinically and pathologically diagnosed six healthy controls and six PBC patients enrolled with written informed consent. Peripheral CD4+T cells were used to extract total RNA. The Agilent microarray of SurePrint G3 human GE 8×60K microarray kit was leveraged to produce gene expression profiles according to manufacturer’s protocols. The GSE115469 dataset contained five samples from primary liver patients, which were used for scRNA sequencing based on the 10× Genomics Chromium Single Cell Kits. A total of 8,444 parenchymal and non-parenchymal cells have obtained the transcriptional profiles based on the CellRanger analysis pipeline. The raw digital matrix of gene expression (namely UMI counts per gene per cell) was filtered, normalized and clustered. Cell was omitted if it has a very high (>0.5) mitochondrial genome transcript ratio or a very small library size (<1500). Using the standard Seurat package ^19^, there were 20 discrete clustered for the scRNA-seq dataset. Using well-known marker genes, these clusters were assigned into 13 distinct cell subpopulations, including portal endothelial cells, cholangiocytes, non-inflammatory macrophages, T cells, γδT cells, inflammatory monocytes/macrophages, natural killer (NK)-like cells, red blood cells (RBCs), sinusoidal endothelial cells, mature B cells, stellate cells, plasma cells, and hepatocytes.

#### 2. Bulk-based expression profiles of PBC

Furthermore, we also downloaded two independent bulk-based expression datasets based on liver tissue (Accession number: GSE159676) and blood (Accession number: GSE119600) from the Gene Expression Omnibus (GEO) database. The dataset of GSE159676 contained six healthy controls and three PBC cases based on fresh frozen liver tissue, which were obtained from explanted livers or diagnostic liver biopsies. The Affymetrix Human Gene 1.0-st array was leveraged to produce bulk-based liver expression profiles with 17,046 probes. With respect to the dataset of GSE119600, there were 47 healthy controls and 90 PBC patients with whole blood samples. The Illumina HumanHT-12 V4.0 expression beadchip was leveraged to produce bulk-based blood transcriptomes with 47,230 probes.

#### 3. GWAS summary statistics on PBC

We downloaded a GWAS summary dataset on PBC from the IEU open GWAS project (https://gwas.mrcieu.ac.uk/) ^5^. There were 2,764 PBC patients and 10,475 healthy controls based on European ancestry in this dataset used for performing a meta-analysis of genome-wide association signals. This GWAS dataset was approved by the University Health Network Research Ethics Board, the Mayo Clinic Institutional Review Board, Etico Indipendente IRCCS Istituto Clinico Humanitas, UC Davis Institutional Review Board and the Oxford Research Ethics Committee ^5^. A standard quality control (QC) pipeline was applied to remove low-quality SNPs. The software package of MaCH ^20^ with the reference of HapMap3 CEU+TSI samples was implemented to perform a genome-wide imputation analysis. There were 1,124,241 SNPs with minor allele frequency > 0.005 and imputation quality score R^2^ > 0.5 included in follow-up analyses.

### Data Processing and Analysis

#### 1. Combination of GWAS summary statistics with scRNA-seq data for PBC

We leveraged a widely-used method of RolyRoly ^21^, which was designed to gain the effects of SNPs near protein-coding genes on cell types contributing to complex traits, to explore genetics-influenced liver cell types for PBC. The regression-based polygenic model was used in the RolyRoly to incorporate GWAS summary data with scRNA-seq data (i.e., GSE115469) for identifying PBC-associated liver cell subpopulations. Let *g*(*i*) represents a given gene relevant to SNP *i, S*_*j*_ = {*i* : *g*(*i*) = *j*} represents a given set with multiple SNPs relevant to the gene *j*, and 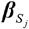 represents a PBC-GWAS-derived effect-size vector of *S*_*j*_ with a *priori* assumption that 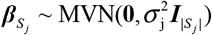. Under the assumption, RolyPoly offers a polygenic linear model for 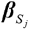:

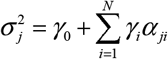

where *γ*_0_ represents an intercept term, α_*ji*_ (*i* = 1,2,…, *N*) represents a group of annotations, for example, cell-type-specific gene expressions, and *γ*_*i*_ is the annotation’s coefficient for α_*ji*_. To fit the observed and expected sum squared SNP effect sizes relevant to each gene, RolyPoly applies the method-of-moments estimators to estimate *γ*_*i*_ by the following formula:

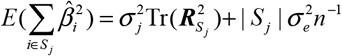

where 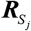 represents the LD matrix of *S*_*j*_. The 1,000 Genome Project European Phase 3 panel ^22^ was used to calculate the SNP-SNP LD information. The major histocompatibility complex (MHC) region was removed due to the highly extensive LD in this region. RolyPoly utilized the 1,000 iterations of block bootstrap to assess standard errors for calculating t-statistic and corresponding P value. P value ≤ 0.05 was interpreted to be of significance.

#### 2. Systematical computational analyses

To prioritize candidate risk genes and functional categories for PBC, we also performed systematical bioinformatics analyses (see Supplemental Methods S1-S7 for details). For using the converging effects of multiple SNPs in a given gene, we applied a multi-variant converging regression model to conduct a gene-level association analysis ^23,24^ of the GWAS summary statistics (Supplemental Methods S1). The disease- and GO-terms enrichment analyses were conducted to reveal the functions of these genetics-risk genes (Supplemental Methods S2). The Multidimensional scaling analysis was used to construct a functional module for identified pathways (Supplemental Methods S3) ^11^, and the GeneMANIA tool ^25^ was used to establish a protein-protein interaction network of these genetics-risk genes (Supplemental Methods S4). To highlight the functional risk genes whose expressions were significantly associated with PBC, we leveraged both S-PrediXcan^26^ and S-MultiXcan ^27^ to combine GWAS summary statistics on PBC with GTEx eQTL data^28^ (Supplemental Methods S5). Furthermore, We referenced the method used in previous studies ^11,24,29,30^ to perform an *in silico* permutation analysis of 100,000 times of random selections *(N* _*Total*_) for validating the consistency of results from S-MultiXcan analysis with other results from three distinct analyses: MAGMA analysis, and S-PrediXcan on liver and blood (Supplemental Methods S6). We also performed a gene-property analysis using the FUMA tool ^31^ to integrate gene-level association signals from GWAS summary statistics on PBC with scRNA-seq data based on liver tissue from the Mouse Cell Atlas ^32^.

### Statistical analysis

Differential gene expression (DGE) analyses between controls and PBC patients of three RNA expression datasets (i.e., GSE93170, GSE159676, and GSE119600) were examined by using the Student’s T-test. P value ≤ 0.05 was of significance. We also performed a co-expression pattern analysis in the dataset of GSE93170 for genetics-risk genes among PBC and healthy controls to evaluate whether the co-expression patterns were changed due to the disease status. The PLINK (v1.90) ^33^ was used to calculate the LD values among SNPs. The hypergeometric test was used to evaluate the significant enrichment for the disease- and GO-term enrichment analysis. The Jaccard distance algorithm ^34^ was used to assess the similarities among pathways.

## Results

### Workflow for the current genomics analysis

To explore the genetics-influenced cell types for PBC, we constructed a computational framework by integrating GWAS summary statistics, eQTL data, and single cell RNA sequencing data. The whole procedure of this framework is shown in Figure 1. There were 8,444 parenchymal and non-parenchymal cells obtained from five samples’ transcriptome profiles in the GSE115469 dataset, and we yielded 13 distinct human liver cell subpopulations (see Methods). We also collected a large-scale GWAS summary statistics on PBC (N = 13,239) for extracting useful genetic information and statistical values. Using a regression-based polygenic model ^21^, we combined the single cell transcriptomes with genetic association signals from the GWAS summary statistics for genetically mapping the single cell landscape of liver tissue implicated in PBC (Figure 1a, and Supplemental Table S1). Additionally, using the comprehensive bioinformatics analyses, including MAGMA gene-level association analysis, S-MultiXcan-based integrative analysis, S-PrediXcan-based integrative analysis, *in silico* permutation analysis, MDS analysis, network-based enrichment analysis, DEG analysis, drug-gene interaction analysis, pathway enrichment analysis, and functional annotation analysis, we highlighted novel genetics-risk genes associated with PBC susceptibility (Figure 1b). Finally, we used genetic association signals to determine the functional subsets of liver- and immune-based cell types relevant to PBC (Figure 1c).

**Figure 1.**
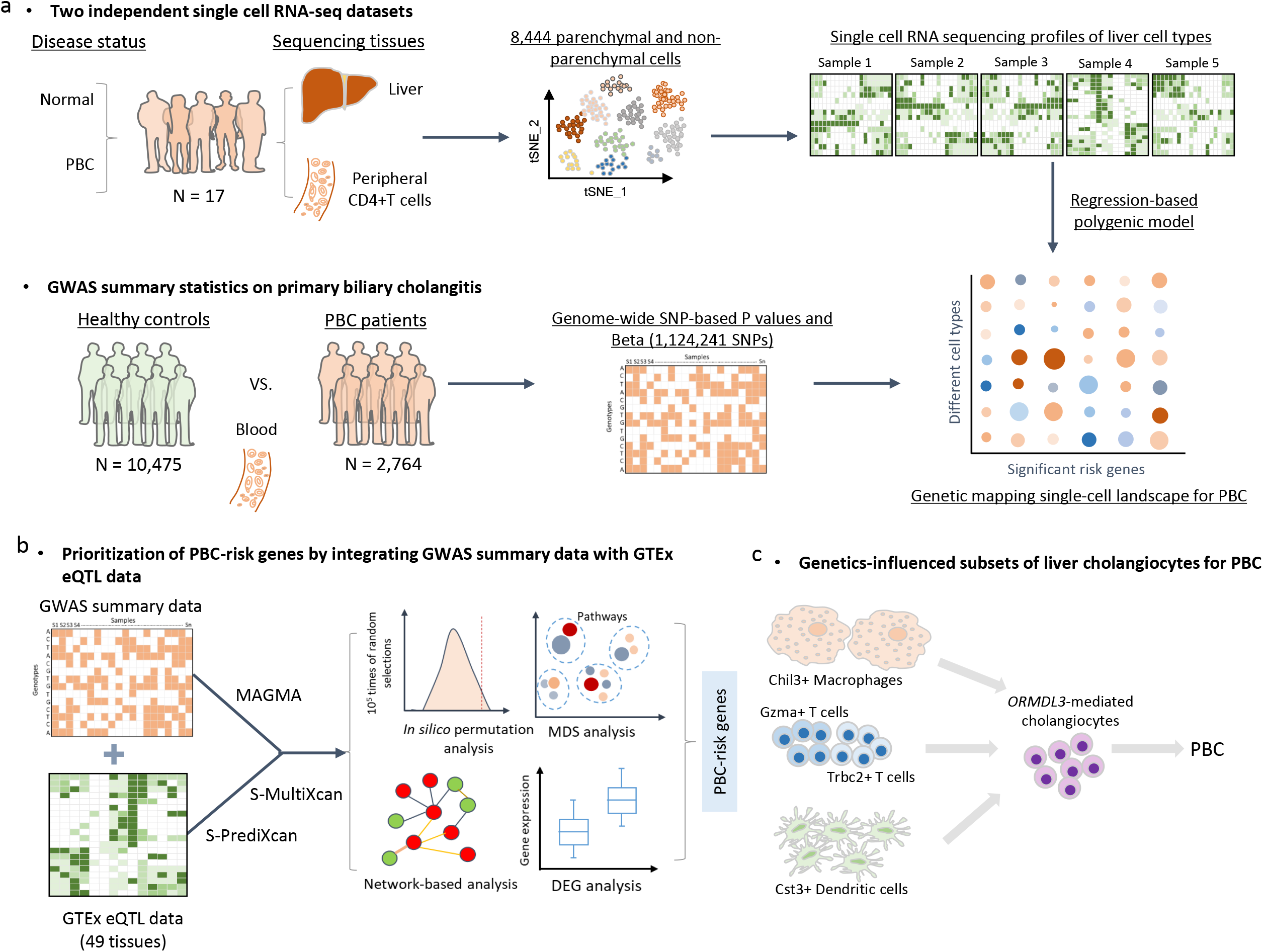
The workflow of current integrative genomics analysis. a) Integrating single cell RNA-sequencing data with GWAS summary statistics on PBC based on a regression-based polygenic model; b) Prioritization of PBC-risk genes by integrating GWAS summary data with GTEx eQTL data; c) Genetics-influenced liver cell subpopulations and its immune microenvironment for PBC.

### Identification of gene-level genetic associations for PBC

To identify the aggregated effects of SNPs in a given gene on PBC, we carried out a gene-level genetic association analysis and found that 563 genes were significantly associated with PBC (FDR ≤ 0.05, Supplemental Figure S1 and Table S2). For example, the top-ranked genes of *HLA-DPA1* (P = 2.69×10^−24^), *IL12A* (P = 6.63×10^−22^), *HLA-DPB1* (P = 1.11×10^−21^), *BTNL2* (P = 1.65×10^−17^), *RXRB* (P = 8.88×10^−17^), *SLC39A7* (P = 8.83×10^−17^), *HLA-DRB1* (P = 3.51×10^−13^), *HLA-DQB1* (P = 6.39×10^−13^), *IRF5* (P = 7.18×10^−11^), *IL12RB2* (P = 3.03×10^−10^), *C6ORF10* (P = 5.58×10^−10^), *HLA-DRA* (P = 5.58×10^−10^), and *CLEC16A* (P = 6.78×10^−10^). Among these significant genes, there were 64 genes, including *IL12A, HLA-DPB1, BTNL2, HLA-DRB1, HLA-DQB1, IRF5*, and *IL12RB2*, reported to be associated with PBC documented in the GWAS catalog database ^5-9^.

Furthermore, we conducted a genome-wide pathway enrichment analysis, and observed that 41 biological pathways were significantly enriched (FDR ≤ 0.05, Supplemental Table S3). Interestingly, the top-ranked pathways were relevant to autoimmune diseases (Supplemental Figure S2a), including Th1 and Th2 cell differentiation (FDR = 3.27×10^−13^), allograft rejection (FDR = 5.18×10^−7^), inflammatory bowel disease (FDR = 1.82×10^−6^), type I diabetes mellitus (FDR = 5.27×10^−6^), and intestinal immune network for IgA production (FDR = 5.27×10^−6^). Based on the MDS analysis, these 41 significant pathways were grouped into five clusters: Th1 and Th2 cell differentiation, allograft rejection, Th17 cell differentiation, cell adhesion molecules, and cytokine-cytokine receptor interactions (Supplemental Figure S2b). These results suggested that these identified genes have a more likelihood to be risk genes for PBC.

### Integrative analysis of GWAS summary statistics with eQTL data for PBC

To further highlight the functional genes whose expressions are associated with PBC, we leveraged the S-MultiXcan software ^27^ to meta-analyze tissue-specific associations across 49 GTEx tissues. There were 268 risk genes whose genetically-associated expression showing notable associations with PBC (FDR < 0.05, Figure 2 and Supplemental Table S4). For example, the top-ranked PBC-associated genes: *HLA-DRB1* (P = 4.95×10^−69^), *HLA-DQA1* (P = 1.03×10^−43^), *HLA-DRB5* (P = 1.17×10^−42^), *BTNL2* (P = 3.26×10^−41^), *EGFL8* (P = 1.37×10^−31^), and *IL12A* (P = 5.42×10^−29^). Among these significant genes, there were 52 risk genes, including *HLA-DRB1, BTNL2, HLA-DPB1, IL12A, HLA-DQB1, IRF5* and *IL12RB2*, having been documented in the GWAS Catalog database (Supplemental Table S4). There was a high consistency of results between MAGMA and S-MultiXcan analysis (232/268 = 86.6%, Supplemental Figure S3).

**Figure 2.**
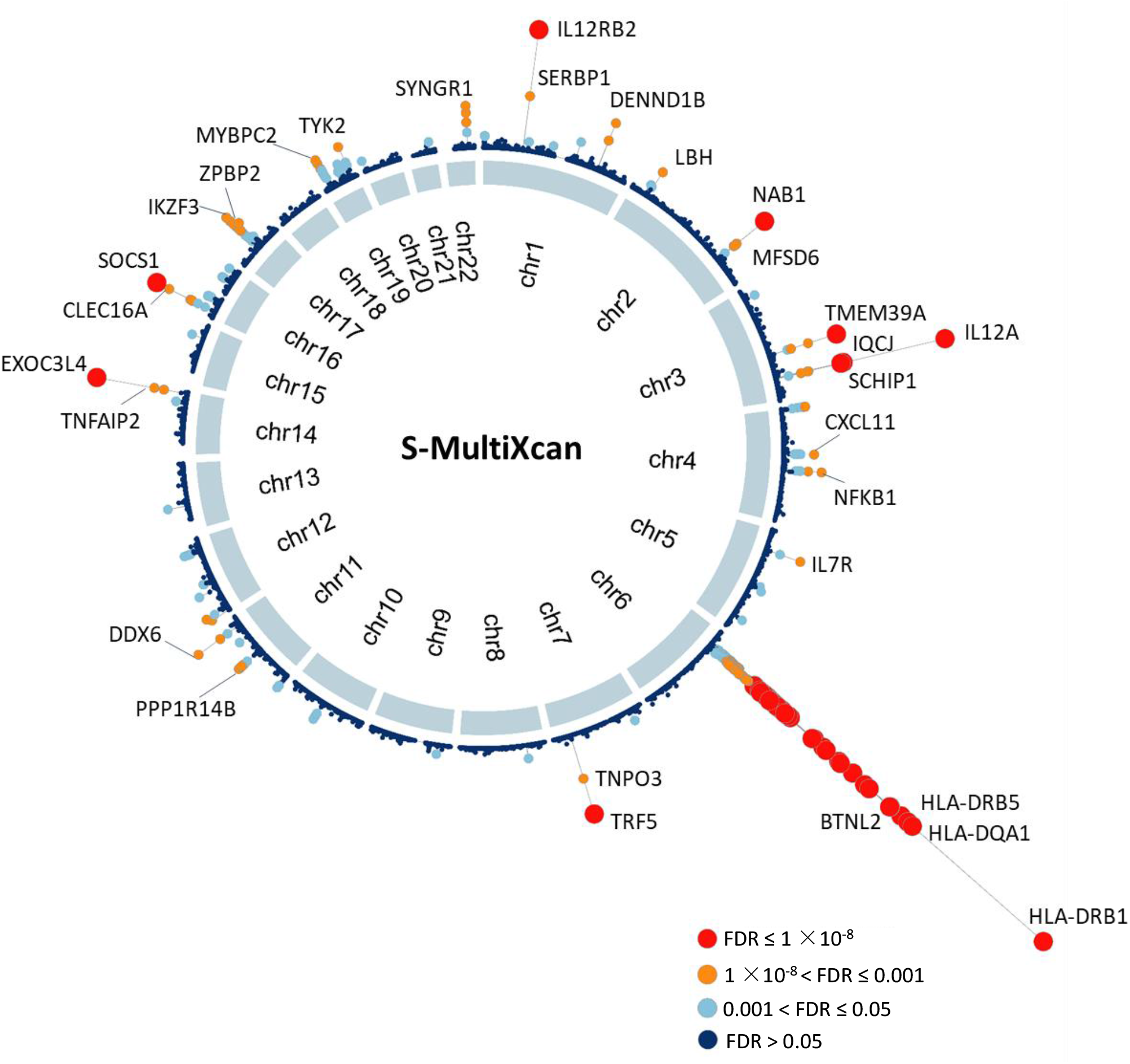
Circus plot showing the results of S-MultiXcan integrative genomics analysis. Note: A circular symbol in the outer ring represents a given gene. Color represents the statistical significance of the gene, where red color marks significant genes with FDR ≤ 1×10^−8^, orange color marks significant genes with FDR is between1×10^−8^ and 0.001, light blue indicates significant genes with FDR ranging from 0.001 to 0.05, and dark blue indicates non-significant genes with FDR > 0.05.

We further validated these risk genes in two PBC-relevant tissues (i.e., liver and blood) using the S-PrediXcan method. 76 and 115 genes were demonstrated to be significantly associated with PBC in liver and blood tissue, respectively (FDR < 0.05, Supplemental Figure S4a-b and Supplemental Tables S5-S6). Notably, 139 genes identified from MAGMA and S-MultiXcan analysis were replicated by using the S-PrediXcan analysis based on liver and blood (Supplemental Figure S4c). Using the Pearson correlation analysis, we observed that significant genes from S-MultiXcan analysis showed remarkable correlations with that from MAGMA and S-PrediXcan analysis on liver and blood (r = 0.14∼0.64, P < 0.05, Supplemental Figure S4d-f). Moreover, by performing three independent *in silico* permutation analyses (see Methods), we found that the number of observed overlapped genes between S-MultiXcan and MAGMA and S-PrediXcan were significantly higher than random events (Empirical P < 1×10^−5^, Supplemental Figures S4g-i and S5a-c). Overall, based on aforementioned integrative genomics analyses, we identified that there were 29 genes showing supportive evidence to involve in PBC susceptibility (Table 1).

**Table 1.** Identification of 29 significant risk genes by using integrative genomics analysis based on GWAS summary statistics and eQTL data.

| Gene name | Z score | P value (S-MultiXcan) | FDR (S-MultiXcan) | FDR (S-PrediXcan on liver) | FDR (S-PrediXcan on blood) | FDR (MAGMA analysis of GWAS) | P value (Differential gene expression analysis) | GWAS_Catalog database or PubMed database |
| --- | --- | --- | --- | --- | --- | --- | --- | --- |
| *CSNK2B* | 1.40 | $7.17 \times 10^{-19}$ | $8.41 \times 10^{-16}$ | $3.22 \times 10^{-11}$ | $2.24 \times 10^{-6}$ | $1.67 \times 10^{-11}$ | $4.96 \times 10^{-4}$ (Blood), $2 \times 10^{-2}$ (CD14+T cell) | Novel gene |
| *LY6G5B* | -7.92 | $2.43 \times 10^{-16}$ | $1.93 \times 10^{-13}$ | $5.52 \times 10^{-11}$ | $1.54 \times 10^{-10}$ | $1.19 \times 10^{-11}$ | Non-significant | Novel gene |
| *DDAH2* | -5.57 | $1.39 \times 10^{-15}$ | $9.99 \times 10^{-13}$ | $2.65 \times 10^{-9}$ | $1.56 \times 10^{-11}$ | $1.69 \times 10^{-9}$ | $2.70 \times 10^{-3}$ (CD14+T cell) | Novel gene |
| *LY6G5C* | -7.82 | $2.40 \times 10^{-15}$ | $1.62 \times 10^{-12}$ | $8.64 \times 10^{-13}$ | $9.05 \times 10^{-12}$ | $1.41 \times 10^{-11}$ | $2.80 \times 10^{-2}$ (Blood) | Novel gene |
| *IRF5* | 7.11 | $2.62 \times 10^{-15}$ | $1.72 \times 10^{-12}$ | $4.29 \times 10^{-13}$ | $1.12 \times 10^{-12}$ | $7.18 \times 10^{-11}$ | $7.10 \times 10^{-3}$ (Liver) | Reported gene |
| *SOCS1* | -3.26 | $9.33 \times 10^{-13}$ | $4.16 \times 10^{-10}$ | $5.25 \times 10^{-10}$ | $7.34 \times 10^{-10}$ | $1.06 \times 10^{-8}$ | $3.50 \times 10^{-2}$ (CD14+T cell) | Reported gene |
| *SYNGR1* | -1.30 | $2.83 \times 10^{-9}$ | $8.88 \times 10^{-7}$ | $7.97 \times 10^{-4}$ | $1.30 \times 10^{-7}$ | $4.13 \times 10^{-2}$ | $1.40 \times 10^{-2}$ (CD14+T cell) | Reported gene |
| *C6orf48* | -4.46 | $9.57 \times 10^{-9}$ | $2.66 \times 10^{-6}$ | $7.83 \times 10^{-4}$ | $6.19 \times 10^{-4}$ | $4.83 \times 10^{-11}$ | $4.16 \times 10^{-6}$ (Blood) | Novel gene |
| *HLA-DMA* | -3.99 | $8.87 \times 10^{-8}$ | $1.90 \times 10^{-5}$ | $1.27 \times 10^{-3}$ | $9.16 \times 10^{-4}$ | $1.62 \times 10^{-8}$ | $2.10 \times 10^{-3}$ (Liver) | Novel gene |
| *SMC4* | -1.34 | $1.00 \times 10^{-7}$ | $2.10 \times 10^{-5}$ | $9.38 \times 10^{-5}$ | $2.25 \times 10^{-6}$ | $2.07 \times 10^{-4}$ | $4.80 \times 10^{-4}$ (Blood) | Novel gene |
| *TCF19* | 4.55 | $1.49 \times 10^{-7}$ | $2.99 \times 10^{-5}$ | $9.46 \times 10^{-5}$ | $2.24 \times 10^{-6}$ | $1.98 \times 10^{-4}$ | Non-significant | Novel gene |
| *ORMDL3* | -2.92 | $7.07 \times 10^{-7}$ | $1.22 \times 10^{-4}$ | $2.35 \times 10^{-3}$ | $7.38 \times 10^{-5}$ | $2.33 \times 10^{-6}$ | Non-significant | Reported gene |
| *KPNA4* | -1.22 | $1.07 \times 10^{-6}$ | $1.82 \times 10^{-4}$ | $2.10 \times 10^{-2}$ | $3.58 \times 10^{-3}$ | $4.67 \times 10^{-4}$ | $3.40 \times 10^{-3}$ (CD14+T cell) | Novel gene |
| *MANBA* | -1.45 | $1.48 \times 10^{-6}$ | $2.35 \times 10^{-4}$ | $2.46 \times 10^{-6}$ | $2.39 \times 10^{-2}$ | $8.30 \times 10^{-8}$ | $9.67 \times 10^{-5}$ (Blood) | Reported gene |
| *MFSD6* | -0.32 | $1.61 \times 10^{-6}$ | $2.51 \times 10^{-4}$ | $2.55 \times 10^{-2}$ | $4.57 \times 10^{-2}$ | $2.06 \times 10^{-3}$ | $3.60 \times 10^{-2}$ (Liver) | Novel gene |
| *MED1* | -2.13 | $1.76 \times 10^{-6}$ | $2.70 \times 10^{-4}$ | $1.65 \times 10^{-2}$ | $1.53 \times 10^{-2}$ | $4.42 \times 10^{-5}$ | Non-significant | Novel gene |
| *IDUA* | 3.95 | $2.37 \times 10^{-6}$ | $3.47 \times 10^{-4}$ | $3.94 \times 10^{-2}$ | $3.87 \times 10^{-2}$ | $8.95 \times 10^{-4}$ | $8.10 \times 10^{-3}$ (Blood) | Reported gene |
| *DGKQ* | 3.24 | $3.86 \times 10^{-6}$ | $5.37 \times 10^{-4}$ | $4.90 \times 10^{-4}$ | $3.36 \times 10^{-4}$ | $2.35 \times 10^{-4}$ | $8.34 \times 10^{-7}$ (Blood) | Reported gene |
| *FCRL3* | -4.40 | $8.14 \times 10^{-6}$ | $1.07 \times 10^{-3}$ | $2.84 \times 10^{-3}$ | $2.84 \times 10^{-3}$ | $5.81 \times 10^{-4}$ | Non-significant | Reported gene |
| *ZCRB1* | 3.22 | $9.34 \times 10^{-6}$ | $1.20 \times 10^{-3}$ | $4.83 \times 10^{-3}$ | $5.02 \times 10^{-3}$ | $1.52 \times 10^{-3}$ | $5.90 \times 10^{-3}$ (Liver); $2.8 \times 10^{-3}$<br>(CD14+T cell); | Novel gene |
| *AGAP5* | -4.28 | $1.29 \times 10^{-5}$ | $1.59 \times 10^{-3}$ | $4.50 \times 10^{-3}$ | $3.40 \times 10^{-3}$ | $3.65 \times 10^{-2}$ | Non-significant | Novel gene |
| *CARM1* | -2.90 | $2.11 \times 10^{-5}$ | $2.49 \times 10^{-3}$ | $1.14 \times 10^{-3}$ | $8.92 \times 10^{-4}$ | $1.86 \times 10^{-4}$ | $2.60 \times 10^{-2}$ (Blood) | Novel gene |
| *UBE2D3* | -4.18 | $2.83 \times 10^{-5}$ | $3.23 \times 10^{-3}$ | $2.76 \times 10^{-3}$ | $5.08 \times 10^{-3}$ | $5.81 \times 10^{-4}$ | $3.40 \times 10^{-2}$ (Blood); $1.6 \times 10^{-4}$<br>(CD14+T cell) | Novel gene |
| *NAAA* | 4.08 | $6.73 \times 10^{-5}$ | $6.88 \times 10^{-3}$ | $1.25 \times 10^{-2}$ | $6.85 \times 10^{-3}$ | $7.41 \times 10^{-4}$ | $3.20 \times 10^{-2}$ (Liver) | Reported gene |
| *SH2B3* | -1.69 | $1.20 \times 10^{-4}$ | $1.17 \times 10^{-2}$ | $2.35 \times 10^{-2}$ | $1.87 \times 10^{-2}$ | $2.44 \times 10^{-3}$ | $1.40 \times 10^{-2}$ (Blood) | Reported gene |
| *CTSH* | 1.75 | $2.05 \times 10^{-4}$ | $1.84 \times 10^{-2}$ | $8.53 \times 10^{-3}$ | $3.46 \times 10^{-2}$ | $2.19 \times 10^{-5}$ | Non-significant | Novel gene |
| *TTC34* | 3.56 | $2.82 \times 10^{-4}$ | $2.39 \times 10^{-2}$ | $5.86 \times 10^{-3}$ | $5.02 \times 10^{-3}$ | $4.98 \times 10^{-3}$ | $9.80 \times 10^{-3}$ (Blood) | Novel gene |
| *METTL1* | 2.19 | $3.11 \times 10^{-4}$ | $2.59 \times 10^{-2}$ | $4.95 \times 10^{-2}$ | $3.81 \times 10^{-2}$ | $1.68 \times 10^{-2}$ | $5.86 \times 10^{-7}$ (Blood) | Novel gene |
| *TSFM* | 3.45 | $3.14 \times 10^{-4}$ | $2.61 \times 10^{-2}$ | $4.87 \times 10^{-2}$ | $3.87 \times 10^{-2}$ | $1.54 \times 10^{-2}$ | $2.80 \times 10^{-4}$ (Blood); $1.5 \times 10^{-3}$<br>(CD14+T cell) | Novel gene |

### Functional analysis of these 29 PBC-associated risk genes

Through mining the PubMed literature and GWAS Catalog database, we found that 10 of 29 identified risk genes have been reported to be associated with PBC in previous GWAS studies, and there were 19 novel genes, including *GSNK2B, LY6G5B, DDAH2*, and *LY6G5C*, newly identified (Supplemental Figure S6a). Among these novel identified genes, several have been demonstrated to be associated with other autoimmune-related diseases, such as *LY6G5B* and *DDAH2* exhibited significant associations with rheumatoid arthritis ^35^ and type 1 diabetes ^36^. Consistently, we performed a phenotype-based enrichment analysis, and found that these 29 risk genes were remarkably enriched in several phenotypes relevant to autoimmune diseases (Supplemental Figure S6b and Table S7), such as autoimmune disease (P = 8.47×10^−8^), type I diabetes mellitus (P = 2.04×10^−4^), immune system diseases (P = 1.41×10^−3^), and juvenile rheumatoid arthritis (P = 2.41×10^−3^).

By conducting a PPI network enrichment analysis, we observed that these risk genes were significantly interacted with each other in a subnetwork (PPI enrichment P = 0.0016, Supplemental Figure S6c), suggesting that these identified genes may have collectively biological functions on PBC risk ^34,37^. Furthermore, we also performed a GO-term enrichment analysis according to three different categories (i.e., CC, MF, and BP), and found that several GO-terms were notably overrepresented (Supplemental Figures S6d, and S7-S9), such as interferon-gamma-mediated signaling pathway (P = 2.8×10^−4^) and neutrophil activation involved in immune response (P =7.3×10^−4^).

Based on three independent expression profiles of liver (GSE159676), blood (GSE119600), and peripheral CD4+T cells (GSE93170), we conducted differential gene expression analysis for these 29 risk genes between PBC and matched control group. We found that 22 of 29 genes (75.86%) showed significantly differential expressions among PBC patients compared with controls (Table 1, Supplemental Figures S10-S12). Based on the dataset of peripheral CD4+T cells, the co-expression patterns among these 29 genes were prominently altered according to the PBC status (Supplemental Figure S12a). These results further support these identified genes have functional effects on the development of PBC.

### Drug-gene interaction analysis for 29 risk genes

Based on the drug-gene interaction analysis, we identified that 20 of 29 genes (68.96%) were enriched in ten potential “druggable” gene categories, including enzyme, druggable genome, kinase, and clinically actionable (Supplemental Figure S6e and Supplemental Table S8). The gene of *SOCS1* was found to be targeted by insulin and aldesleukin (Supplemental Figure S13), of which both have been applied to treat autoimmune diseases, including systemic lupus erythematosus ^38^, type 1 diabetes mellitus ^39^, and HIV ^40^. Additionally, *TCF19* gene was targeted by nevirapine, *NAAA* was targeted by cyclopentyl palmitate, *CARM1* was targeted by BIIB021 and eosin, and *SH2B3* was targeted by ruxolitinib and candesartan. Previous studies ^41^ have demonstrated that the Janus kinase (JAK)-inhibitor ruxolitinib significantly influenced dendritic cell differentiation and function resulting in impaired T-cell activation, which could be used for the treatment of autoimmune diseases. These results provide a good drug repurposing resource to develop effective therapeutics for PBC.

### Identification of genetically-influenced liver cell subpopulations for PBC

We performed a gene-property analysis based on mouse liver tissue with immune cells, and found that PBC-associated genes were enriched in several immune-related cell types (Figure 3a and Supplemental Table S9), including Cst3^+^ dendritic cell (P = 5.8×10^−3^), Trbc2^+^ T cell (P = 7.1×10^−3^), Chil3^+^ macrophage (P = 0.0167), and Gzma^+^ T cell (P = 0.0304), which were in line with the results in previous studies ^42,43^. To uncover genetics-regulatory cell subpopulations associated with PBC, we used a regression-based polygenic model to combine GWAS summary statistics on PBC with scRNA-seq data with 13 distinct human liver cell types. We found that cholangiocytes showed a notable enrichment by PBC-relevant genetic association signals (Permuted P < 0.05, Figure 3b and Supplemental Table S10). These results are consistent with previous evidence that an immune-mediated injury of cholangiocytes contributes risk to PBC ^44,45^.

**Figure 3.**
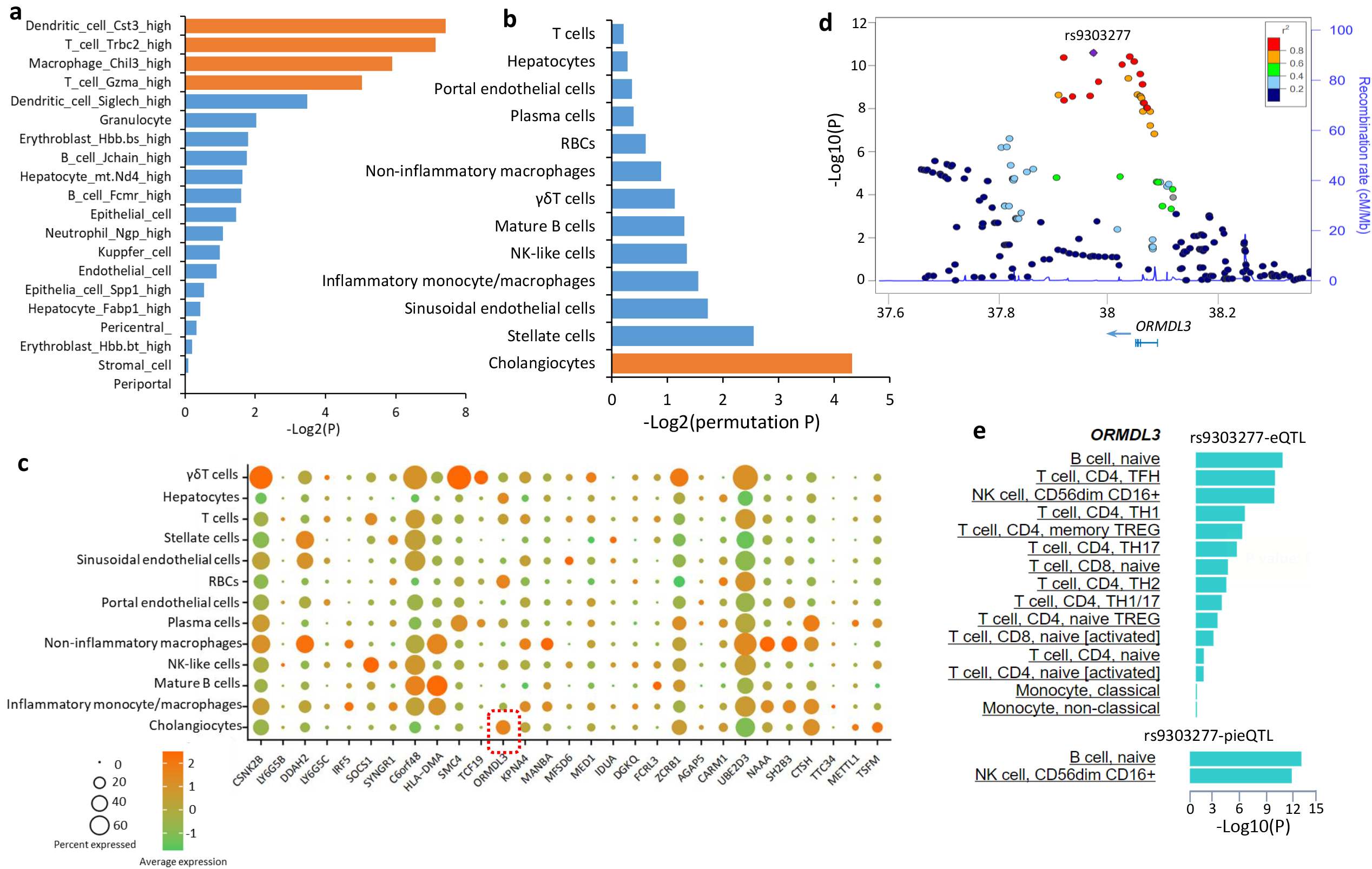
Combination of GWAS summary statistics on PBC with scRNA-seq data on human liver tissue. a) Results of MAGMA gene-property analysis based on the web-access tool of FUMA. Gray color means non-significant enrichment. Orange color stands for significant enrichment. b) Bar graph showing genetics-influenced liver cell subpopulations for PBC. Orange color represents PBC-relevant genetic association signals showing a significant enrichment in cholangiocytes. c) Dot plot showing the expression percentage of 29 PBC-risk genes for each cell type from human liver tissue. The color stands for the average expression of each gene in each cell type, and the size of circular symbol indicates the percentage of a given gene expressed in each cell type. d) Regional association plot for *ORMDL3* gene of GWAS summary statistics using the *LocusZoom* software. The color bar illustrates SNPs’ LD information with rs9303277 as shown in the color legend. e) Immune cell-type specific eQTL and promoter-interacting eQTL (pieQTL) of *ORMDL3* for rs9303277 using the DICE database.

Using the specificity algorithm in the MAGMA method ^23^, we noticed that the risk gene of *ORMDL3* exhibited the highest expression in cholangiocytes than other cell types (Figure 3c and Supplemental Table S11). The majority of *ORMDL3*-expressing cells were cholangiocytes with a relative high percentage of 22.38%, reminiscing that *ORMDL3* was demonstrated to be significantly associated with PBC in earlier studies ^7,9,46^. The top-ranked risk SNP associated with *ORMDL3* is rs9303277 (P = 2.57×10^−11^, Figure 3d). This SNP showed significant cell-specific eQTL of *ORMDL3* among naïve B cell (P = 5.8×10^−10^), TFH^+^CD4^+^T cell (P = 3.8×10^−9^), CD56 ^dim^ CD16^+^ NK cells (P = 4.4×10^−9^), TH1^+^ CD4^+^ T cells (P = 6.0×10^−6^), and memory TREG^+^ CD4^+^ T cells (P = 1.1×10^−5^), and exhibited notable cell-specific promoter-interacting eQTL of *ORMDL3* among naïve B cell (P = 4.2×10^−13^) and CD56 ^dim^ CD16^+^ NK cells (P =5.0×10^−12^) (Figure 3e and Supplemental Methods S8). Among these immune cell types, the CC genotype of rs9303277 shows prominent association with higher expression of *ORMDL3* compared with other genotypes (Supplemental Figure S14a-k).

Growing attentions have concentrated on the role of *ORMDL3* in the development of inflammatory diseases, including PBC^7,9,47^. In view of the main goal of current study was to characterize genetics-modulated liver cell subpopulations for PBC, the majority of our subsequent analyses focused on revealing the effects of *ORMDL3*-mediated cholangiocytes.

### Characterization of biological functions of ORMDL3-mediated cholangiocytes

We compared the expression profiles of *ORMDL3*^*+*^ cholangiocytes with *ORMDL3*^-^ cholangiocytes, and found that there were 77 significantly differential expressed genes (DEGs) with six up-regulated DEGs and 71 down-regulated DEGs among *ORMDL3*^*+*^ cholangiocytes (FDR < 0.05, Figure 4a and Supplemental Tables S13-S14). With regard to six up-regulated DEGs, two genes of *GALNT1* and *HERPUD2* have been reported to attenuate inflammatory responses ^48,49^. There were seven significant KEGG pathways overrepresented, and the most significant one is Herpes simplex virus 1 infection (FDR < 0.05, Figure 4c and Supplemental Table S15). Among these 71 down-regulated DEGs, *HLA-DRA* has been shown to be associated with PBC in previous GWASs ^7^, and *CXCL8, CCL3, CXCL1, TIMP1, SPP1*, and *IRF1* were inflammatory and cytokine genes, which have been reported to be linked with the chemotaxis of immune cells that efflux to the site of cytokine storms in response to ongoing tissue damage ^50,51^. *IFI16* is reported to be an innate immune sensor for intracellular DNA ^52^.

**Figure 4.**
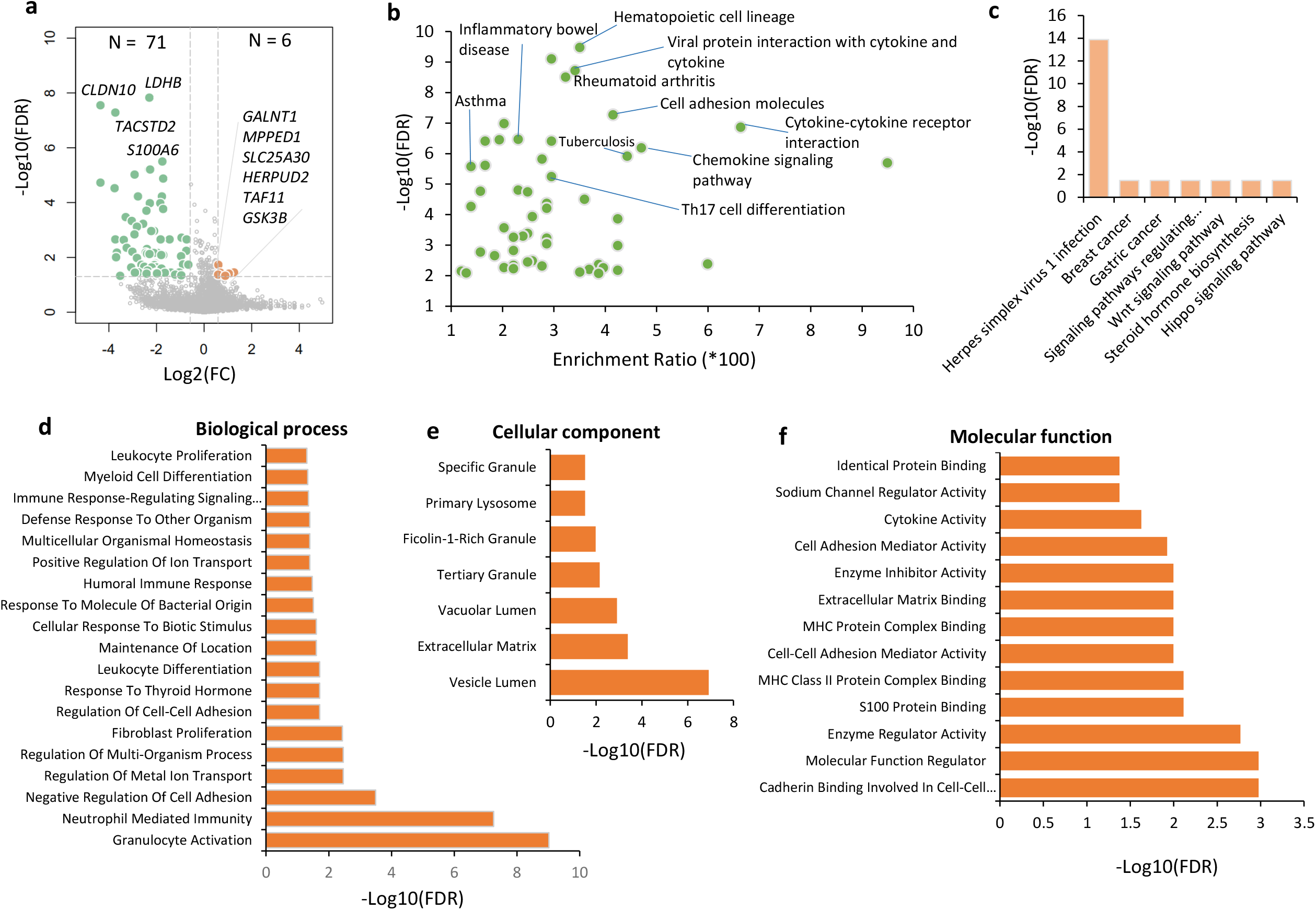
Differential gene expression analysis between *ORMDL3*^+^ and *ORMDL3*^-^ cholangiocytes. a) Volcano plot showing the differentially expressed genes (DEGs) between *ORMDL3*^+^ cholangiocytes and *ORMDL3*^-^ cholangiocytes. Green color represents 71 significantly down-regulated DEGs, and orange color represents 6 significantly up-regulated DEGs. b)-c) Pathway enrichment analyses of (b) 71 down-regulated DEGs and (c) 6 up-regulated DEGs based on the KEGG pathway resource. d)-f) GO enrichment analyses of 71 down-regulated DEGs according to three categories: (d) biological process, (e) cellular component, and (f) molecular function.

Pathway enrichment analysis revealed that there were 55 significant KEGG pathways enriched by these 71 down-regulated DEGs (FDR < 0.05, Figure 4b and Supplemental Table S14), such as, hematopoietic cell lineage, inflammatory bowel disease, rheumatoid arthritis, Th17 cell differentiation, cytokine-cytokine receptor interaction, and chemokine signaling pathway, of which several have been demonstrated to implicate in inflammatory-related diseases ^11,53,54^. To further explore the biological functions of these 71 down-regulated DEGs, we conducted GO-term enrichment analysis according to three categories of biological process (BP), cellular component (CC), and molecular function (MF). There were 19 BP-terms, 7 CC-terms, and 13 MF-terms showing notable enrichments, respectively (FDR < 0.05, Figure 4d-f and Supplemental Tables S16-S18). These functional terms were largely linked with immune-related functions, such as granulocyte activation, neutrophil mediated immunity, S100 protein binding, and MHC class II protein complex binding. Moreover, these genes were prominently enriched in six disease-related terms, including liver cirrhosis, chemical and drug induced liver injury, acute coronary syndrome, acute kidney injury, neoplasm invasiveness, and chronic obstructive airway disease (FDR < 0.05, Supplemental Figure S15 and Table S19). These results suggest that *ORMDL3*^-^ cholangiocytes have immune-modulatory effects on PBC risk.

## Discussion

Using an integrative genomics approach based on multiple layers of evidence, we identified that the genetics-influenced expression of 29 risk genes were remarkably associated with PBC. Among them, 10 genes, including *IRF5*^5,6,8,9^, *SOCS1*^5^, *SYNGR1*^5,7^, *ORMDL3*^7,9,46^, *MANBA*^5^, *IDUA* ^5^, *DGKQ* ^5^, *FCRL3* ^55^, *NAAA* ^56^, and *SH2B3* ^5^, have been documented to be associated with PBC. There were 19 newly identified PBC-risk genes, such as *GSNK2B, LY6G5B, DDAH2, C6orf48*, and *HLA-DMA*. The novel risk gene of *DDAH2*, which encodes a dimethylarginine dimethylaminohydrolase, has been reported to be associated with the autoimmune diseases of rheumatoid arthritis ^57^ and type I diabetes ^58^. The *HLA-DMA* gene, which belongs to the major histocompatibility complex (MHC) class II alpha chain paralogues, has been widely reported to be associated with autoimmune diseases, including type I diabetes mellitus ^59^, rheumatoid arthritis ^60^, and systemic lupus erythematosus ^61^. The *ORMDL3* gene, which has been associated with PBC susceptibility ^7,9,46^, is related to biological functions of innate immune system and metabolism. Moreover, *ORMDL3* has also been extensively reported to be linked with other inflammatory diseases, including childhood asthma ^62^, inflammatory bowel diseases ^53^, rheumatoid arthritis ^54^, and Crohn’s disease ^63^. Functional enrichment analyses uncovered that these risk genes were notably enriched in several biological processes or disease-terms that are relevant to autoimmune phenotypes ^64,65^. Together, these results suggest these identified genes are more likely to be genuine genes implicated in PBC risk.

To examine the liver and its immune microenvironment for PBC, we performed the gene-property analysis and regression-based polygenic analysis based on human and mouse liver tissues with immune cells. Our gene-property analysis identified that four immune cell subpopulations, Cst3^+^ dendritic cell, Chil3^+^ macrophage, Trbc2^+^ T cell, and Gzma^+^ T cell, are significantly associated with PBC risk. Dendritic cells have been shown to be relevant to the pathogenesis of PBC ^42,66^. Earlier studies have demonstrated that an intense biliary inflammatory CD8+ and CD4+ T cell response has been used for characterizing PBC ^67^. Moreover, dendritic cells are critical for inducing antigen-specific T-cell tolerance and for activating the self-specific T cells, which have central roles in many aspects of the pathogenesis of autoimmune liver diseases ^68^. Macrophages are key components of the innate immune system, and have critical immuno-modulatory and tissue-repairing roles in reducing immune responses and enhancing tissue regeneration ^69^. Multiple lines of evidence ^70^ have demonstrated that the infiltration of macrophages in diseased tissues is considered to be a hallmark of several autoimmune diseases, including PBC ^43^. Recently, Dorris et al. ^71^ reported that the PBC susceptibility gene *C5orf30* modulates macrophage-mediated immune regulations. Overall, these results suggest that three main immune cells, dendritic cells, macrophages, and T cells, are vulnerable to be influenced by PBC-relevant genetic association signals.

Moreover, our regression-based polygenic analysis suggested that cholangiocytes were significantly influenced by PBC-related genetic association signals. The non-parenchymal cholangiocytes have been reported to be injury in numerous human diseases termed as cholangiopathies ^72^, including PBC ^72,73^. Recently, Banales and coworkers ^72^ have demonstrated that cholangiocytes play pivotal roles in innate and adaptive immune responses relevant to immune-mediated cholangiopathies. Erice et al. ^74^ reported that microRNA-506 induces PBC-like features in human cholangiocytes and promotes the activating processes of immune responses. We further found that *ORMDL3*^*+*^ cholangiocytes predisposed to attenuate the release of inflammatory factors, whereas *ORMDL3*^-^ cholangiocytes, containing numerous highly-expressed inflammatory cytokine genes, tend to have genetics-mediated immune regulation for PBC risk. Many biological pathways or functional terms associated with autoimmune-related diseases have been significantly enriched, such as Th17 cell differentiation, cytokine-cytokine receptor interaction, and chemokine signaling pathway, inflammatory bowel diseases, and rheumatoid arthritis ^11,53,54^. Overall, these results suggest that down-expression of *ORMDL3* has a vital role in the immune-mediated injury of cholangiocytes conferring high risk to PBC.

There exist some limitations should be cautious. Although we leveraged integrated bioinformatics methods to highlight PBC-associated risk genes based on multiple omics data, there were many potential risk genes with suggestive evidence for PBC as shown in the supplemental tables needed to be further studied. Furthermore, due to the heterogeneity across different datasets used in the present investigation, we leveraged different statistical methods for multiple testing correction for each dataset, such as FDR < 0.05 for MAGMA-based gene association analysis and S-MultiXcan analysis, permuted P < 0.05 for genome-wide pathway enrichment analysis, Bonferroni corrected P < 0.05 for gene-property analysis, and empirical P value < 0.05 for *in silico* permutation analysis. In view of current integrative genomic analysis is only based on samples derived from European ancestries, more studies based on other ancestries should be performed to validate the effects of these risk genes on PBC.

In conclusion, current study provides multiple lines of evidence to support 29 genes including 19 novel genes are remarkably associated with PBC susceptibility. To the best of our knowledge, this is the first study to parse genetics-influenced human liver cell subpopulations that contribute risk to PBC, and found that *ORMDL3*^-^ cholangiocytes potentially play important immune-regulatory roles in the pathogenesis of PBC. Current study gives several highlighted genes and genetics-risk cell type for functional experimentations to reveal the genetic mechanisms of PBC.

## Data Availability

All the GWAS summary data applied in the present analysis can be obtained from the IEU open GWAS project (https://gwas.mrcieu.ac.uk/). The GTEx eQTL data (version 8) can be gained from the Zenodo repository (https://zenodo.org/record/3518299#.Xv6Z6igzbgl). All scripts used in the Methods are deposited in the public available GitHub repository: https://github.com/mayunlong89/PBC_project.

## Acknowledgements

We appreciate Cordell and his colleagues who have deposited and shared GWAS summary data on primary biliary cirrhosis in the IEU open GWAS project.

