## Supplemental Figures for "Combination of single cell sequencing data and GWAS summary statistics reveals genetically-influenced liver cell types for primary biliary cholangitis"

**Supplemental Figure S1. Circus plot showing the results of gene-level genetic association analysis of GWAS summary statistics on PBC.** Note: This analysis was performed using the MAGMA software. A circular symbol in the outer ring stands for a given gene, and color of a circular symbol represents the statistical significance of a specific gene, i.e., red color marks genes significantly associated with PBC with FDR ≤ 1×10^-8^, orange color marks genes significantly associated with PBC with FDR ranging from 1×10^-8^ to 0.001, light blue indicates genes significantly associated with PBC with FDR ranging from 0.001 to 0.05, and dark blue indicates genes showing non-significant associations with PBC (FDR > 0.05). There were 563 genes showing significant associations with PBC (FDR ≤ 0.05).

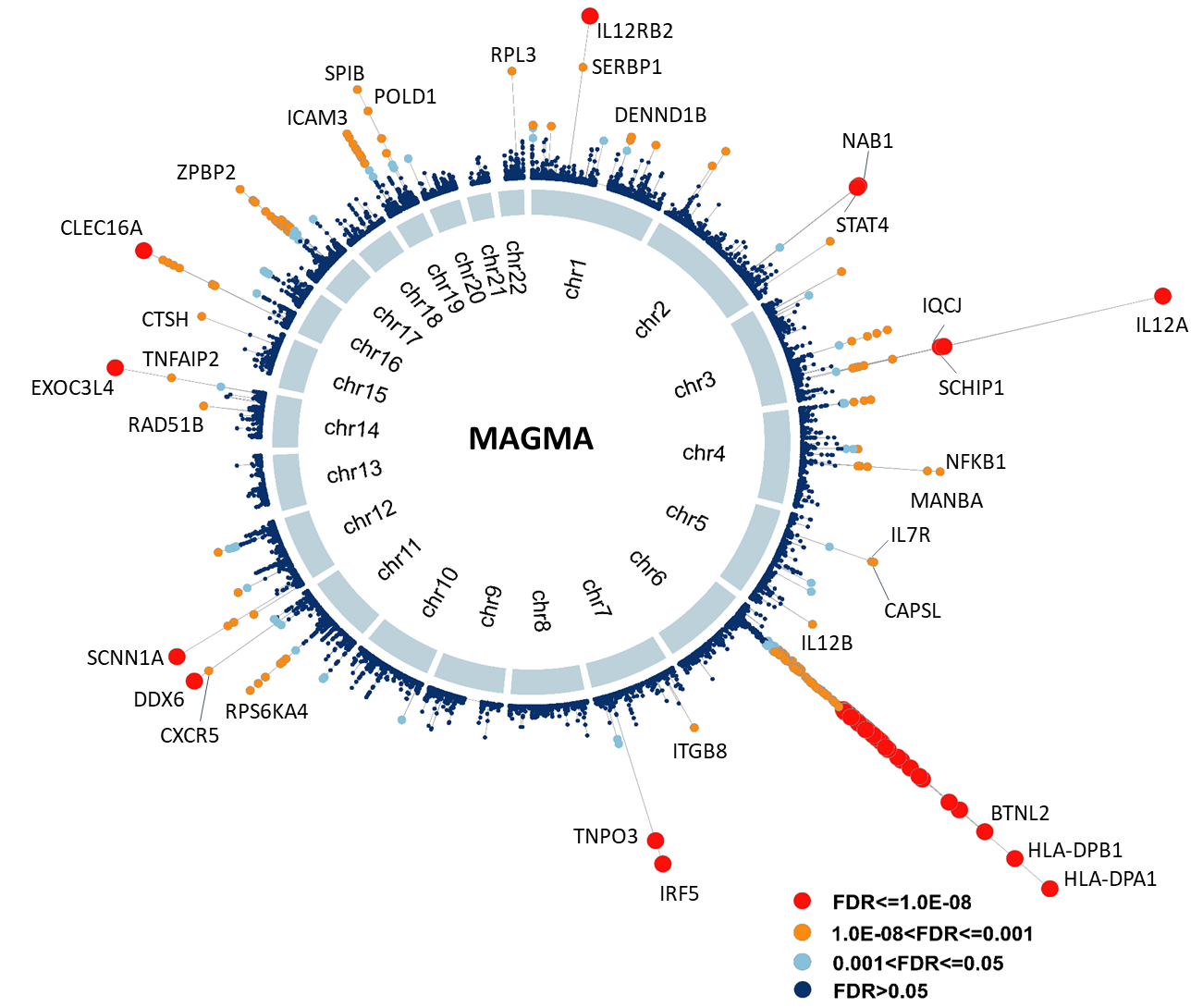

**Supplemental Figure S2. MAGMA-based functional annotation analysis for GWAS summary data.** a) MAGMA-based pathway enrichment analysis based on GWAS summary data on PBC. b) Multidimensional scaling analysis of 41 significant pathways identified MAGMA-based pathway analysis. Note: Circular ring size represents the number of each enriched pathway. Color stands for the exponent function transformed *beta* value of each enriched pathway. Number of circular ring in the plot represents the pathway ID as shown in the Supplemental Table S3.

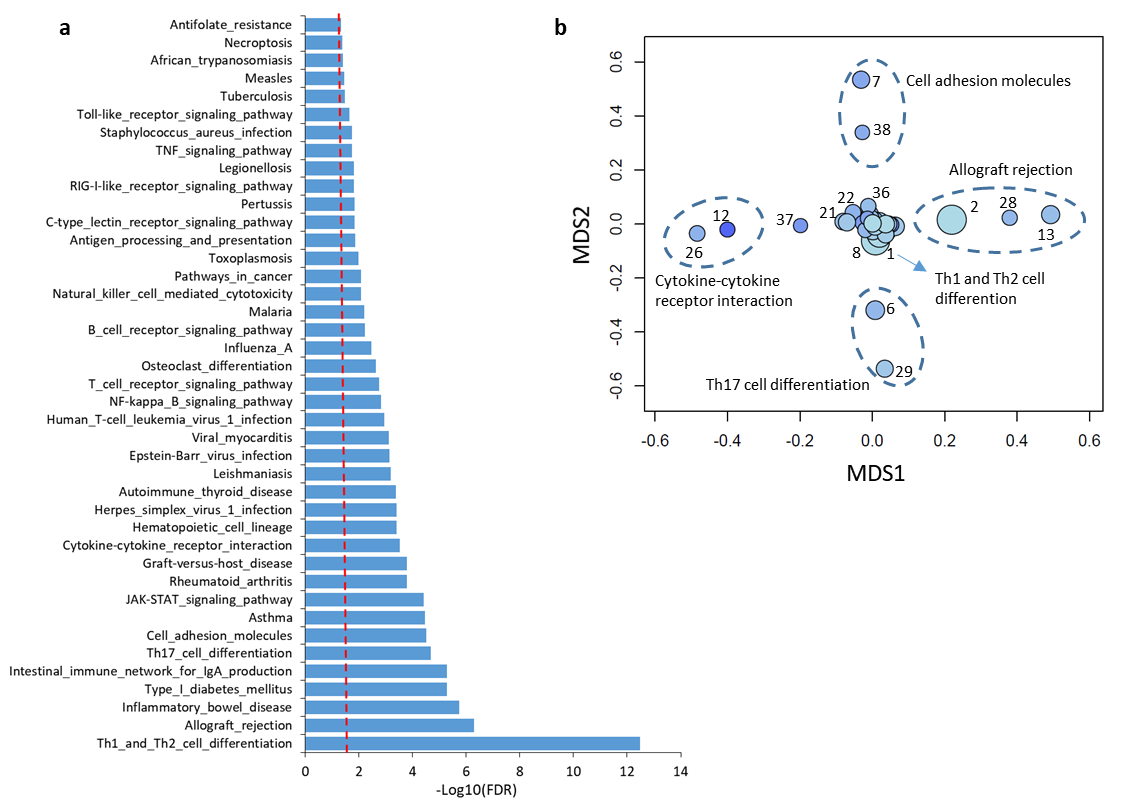

**Supplemental Figure S3. Venn plot shows the overlap of results between MAGMA and S-MultiXcan analysis.** Note: There is a high overlap rate of 86.6% (232/268) between significant genes from S-MultiXcan analysis and that from MAGMA analysis.

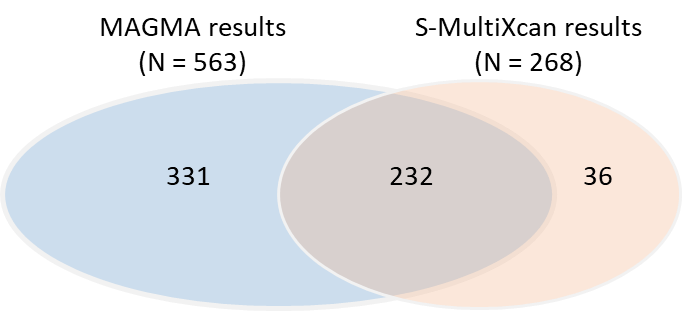

**Supplemental Figure 4. Results from integrative genomics analyses**. a)-b) Circus plot demonstrates the results of S-PrediXcan integrative genomics analysis on (a) liver tissue and (b) blood tissue. c) Venn plot showing the overlapped genes between S-MultiXcan, MAGMA, and S-PrediXcan on both liver and blood. d)-f) Pearson correlation of top-ranked genes identified from S-MultiXcan analysis with that from (d) MAGMA analysis, (e) S-PrediXcan analysis on liver, and (f) S-PrediXcan analysis on blood. g-i) *In silico* permutation analysis of 100,000 random selections for the overlapped genes between S-MultiXcan and (g) MAGMA, (h) S-PrediXcan on liver, and (i) S-PrediXcan on blood.

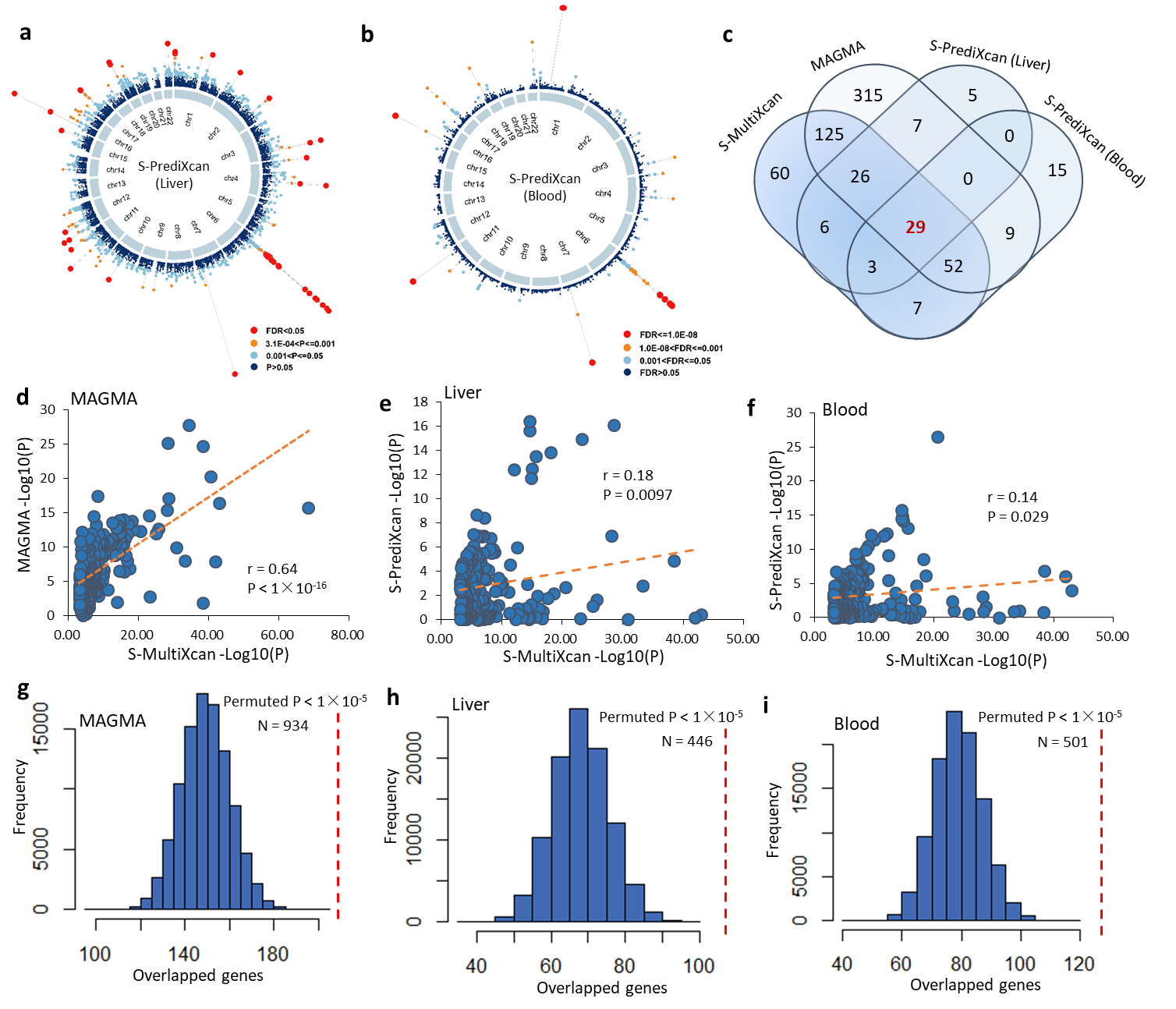

**Supplemental Figure S5. Permutation analysis of 100,000 times for results from MAGMA and S-PrediXcan (liver and blood) compared with results from S-MultiXcan across multiple tissues.** a) for the comparison between MAGMA and S-MultiXcan (the observed number of overlapped genes is 232, permuted P < 1×10^-5^); b) for the comparison between S-PrediXcan on liver and S-MultiXcan (the observed number of overlapped genes is 64, permuted P < 1×10^-5^), and c) for the comparison between S-PrediXcan on blood and S-MultiXcan (the observed number of overlapped genes is 91, permuted P < 1×10^-5^). Note: the observed gene from each method was based on the significant threshold at FDR ≤ 0.05.

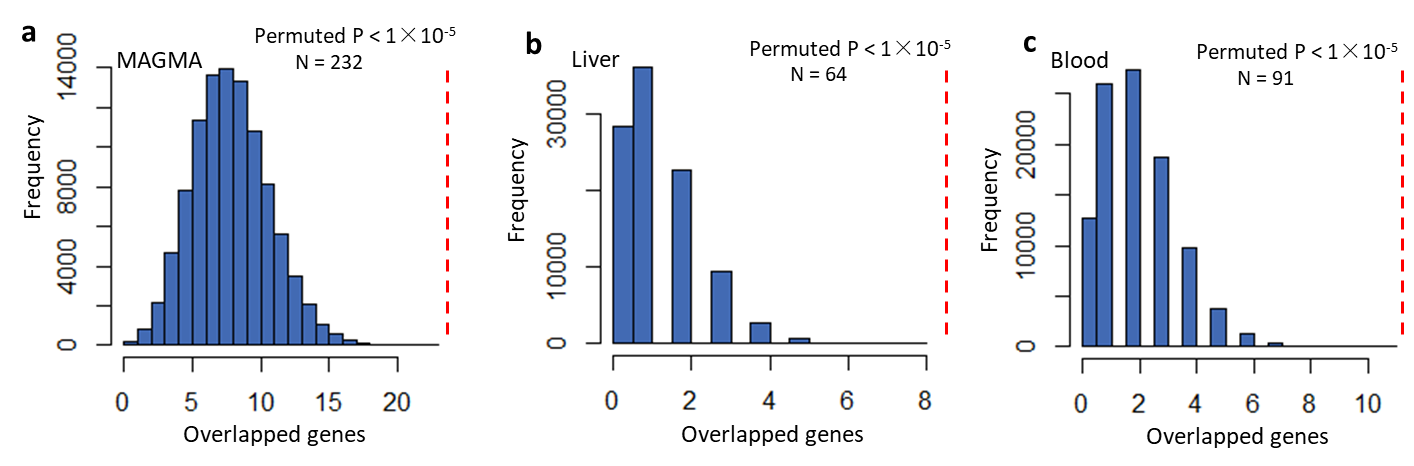

**Supplemental Figure S6. Functional characterization of 29 risk associated with PBC**. a) Schematic diagram showing the literature mining and GWAS catalog searching for novel PBC-risk genes. b) Phenotype-based enrichment analysis based on the GLAD4U database for 29 risk genes. c) PPI network analysis of 29 identified risk genes based on both STRING and GeneMANIA database. d) Functional enrichment analysis of GO-term of biological process for 29 PBC-risk genes. e) Drug-gene interaction analysis showing 10 druggable gene categories based on the DGIdb database.

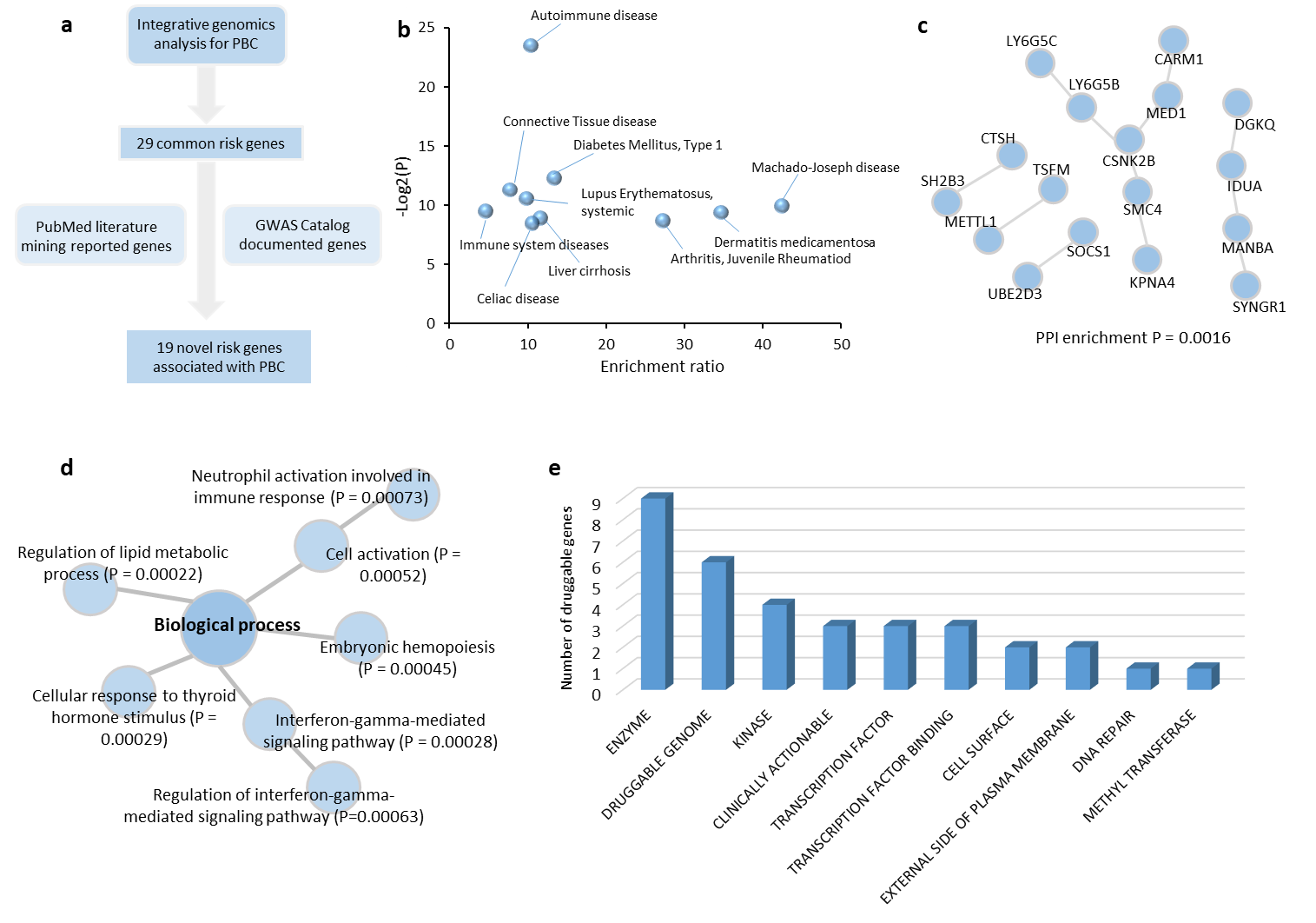

**Supplemental Figure S7. Functional enrichment analysis of GO-term of biological process for 29 PBC-risk genes.**

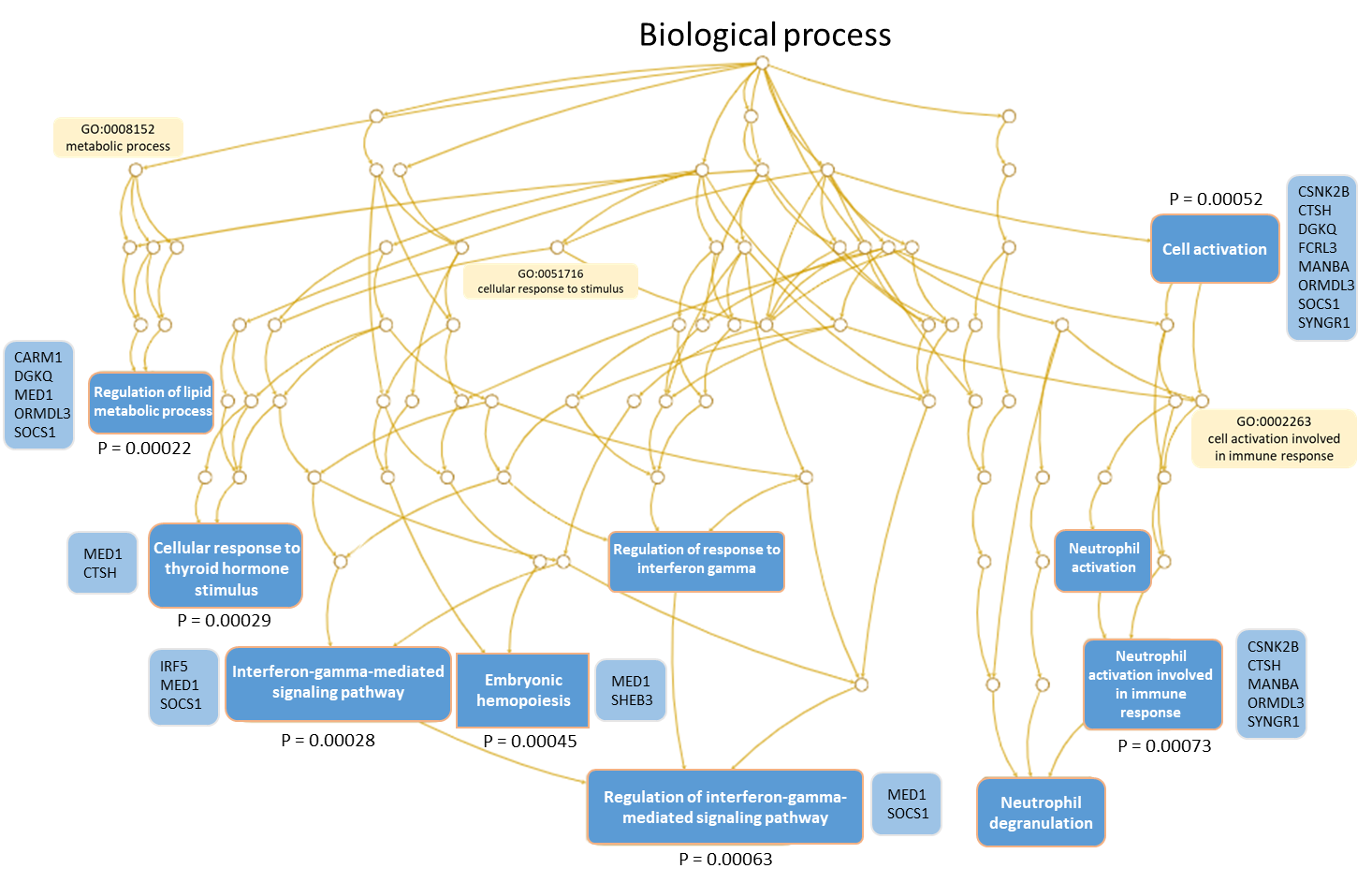

**Supplemental Figure S8. Functional enrichment analysis of GO-term of cellular components for 29 PBC-risk genes.**

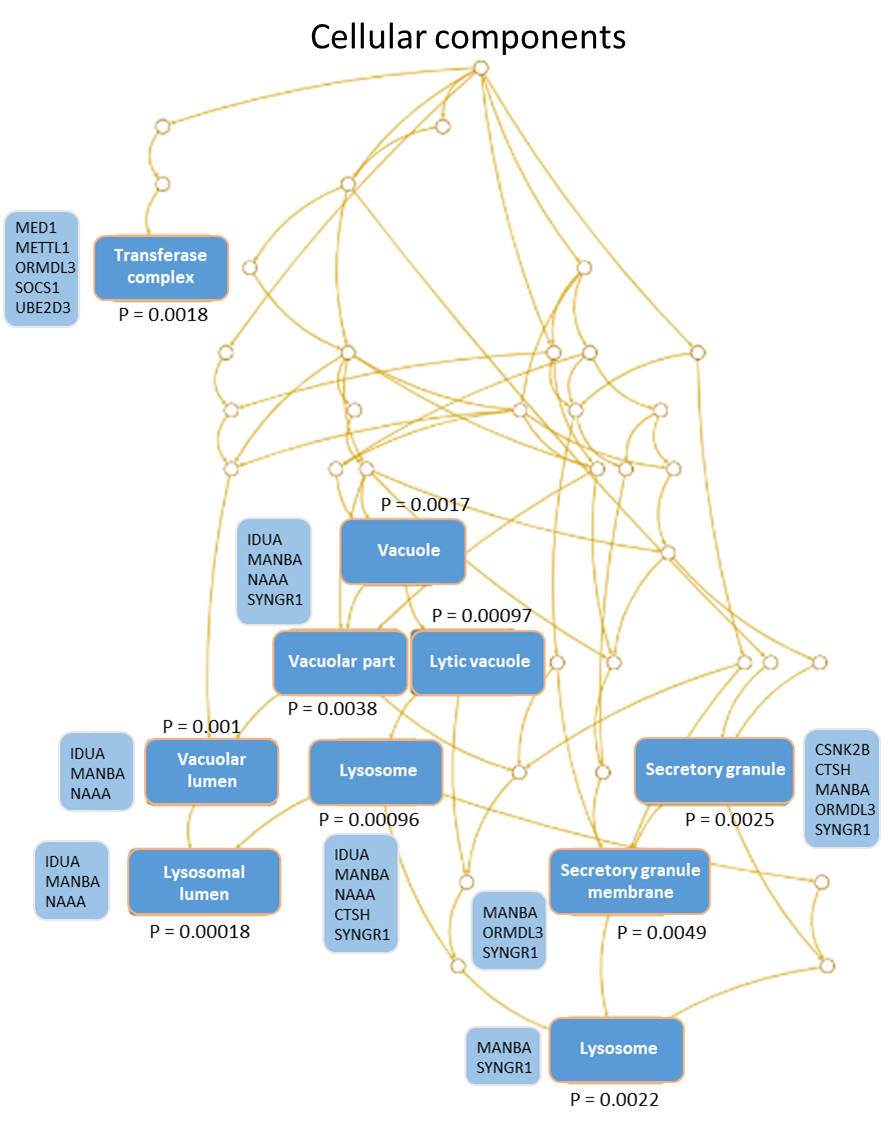

**Supplemental Figure S9.** **Functional enrichment analysis of GO-term of molecular function for 29 PBC-risk genes.**

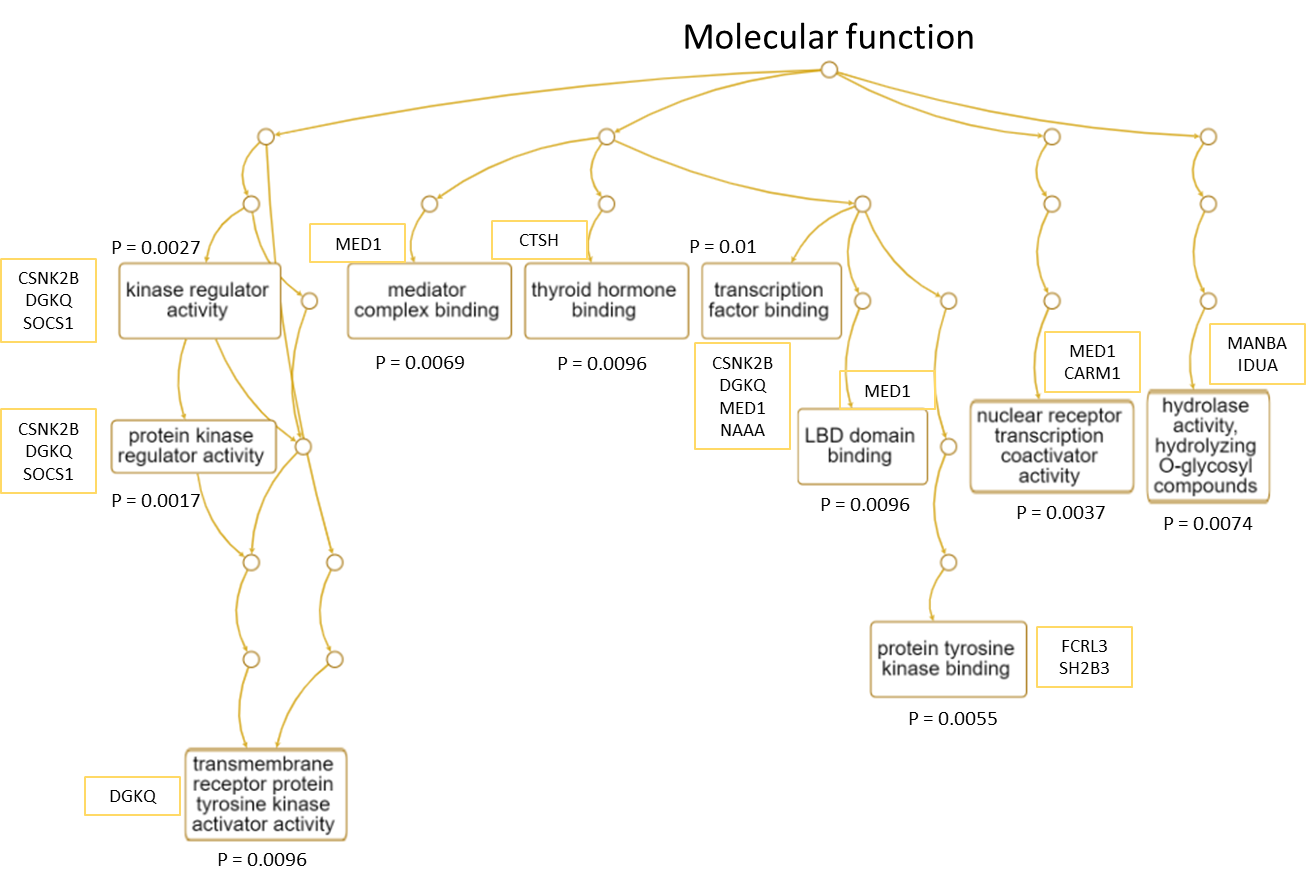

**Supplemental Figure S10. Differential gene expression analysis of 29 risk genes based on bulk RNA profiles of liver tissue (Dataset of GSE159676).** a)-e) Violin plot showing the significantly differential expressions of genes in the dataset of GSE159676 between healthy controls and PBC patients.

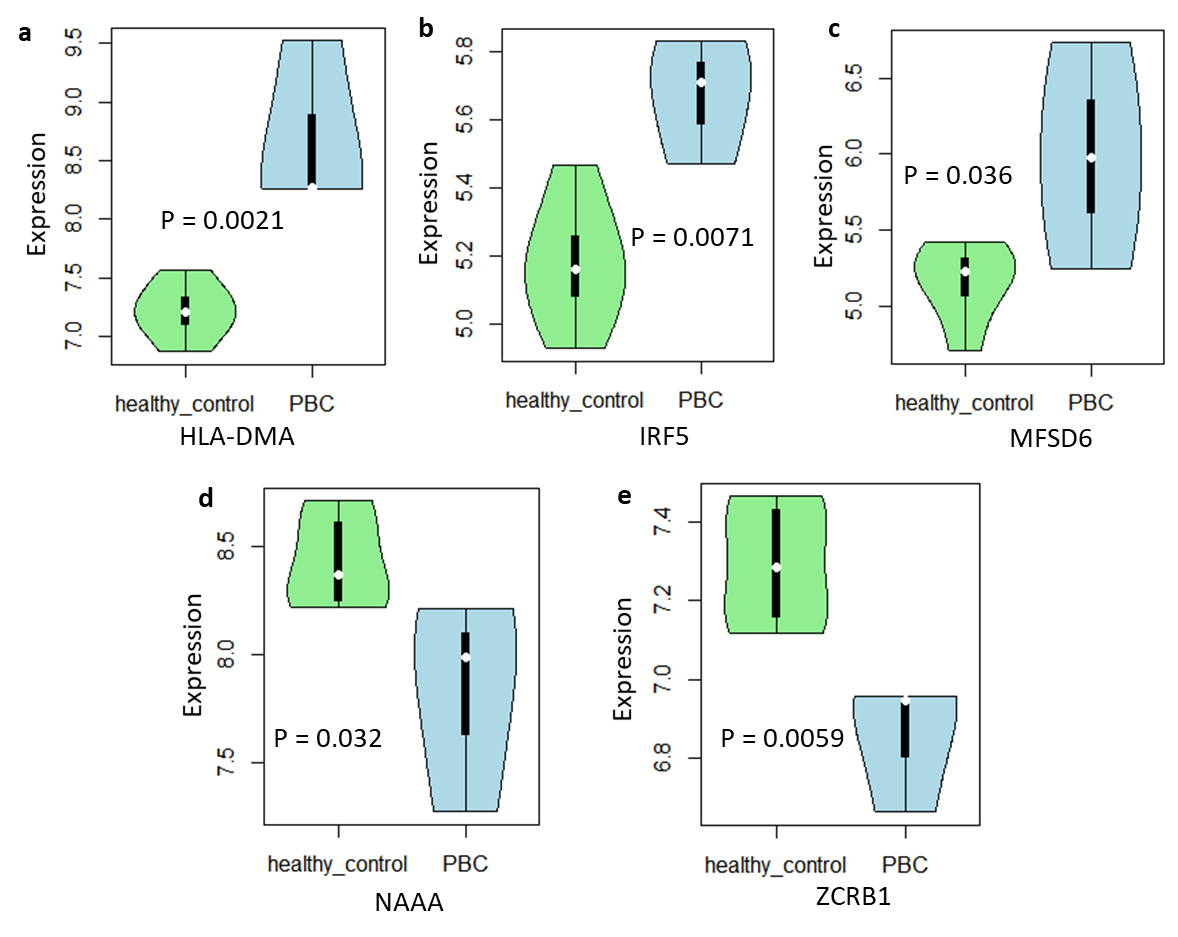

**Supplemental Figure S11. Differential gene expression analysis of 29 risk genes based on bulk RNA profiles of blood tissue (Dataset of GSE119600).** a)-m) Violin plot showing the significantly differential expressions of genes in the dataset of GSE119600 between healthy controls and PBC patients.

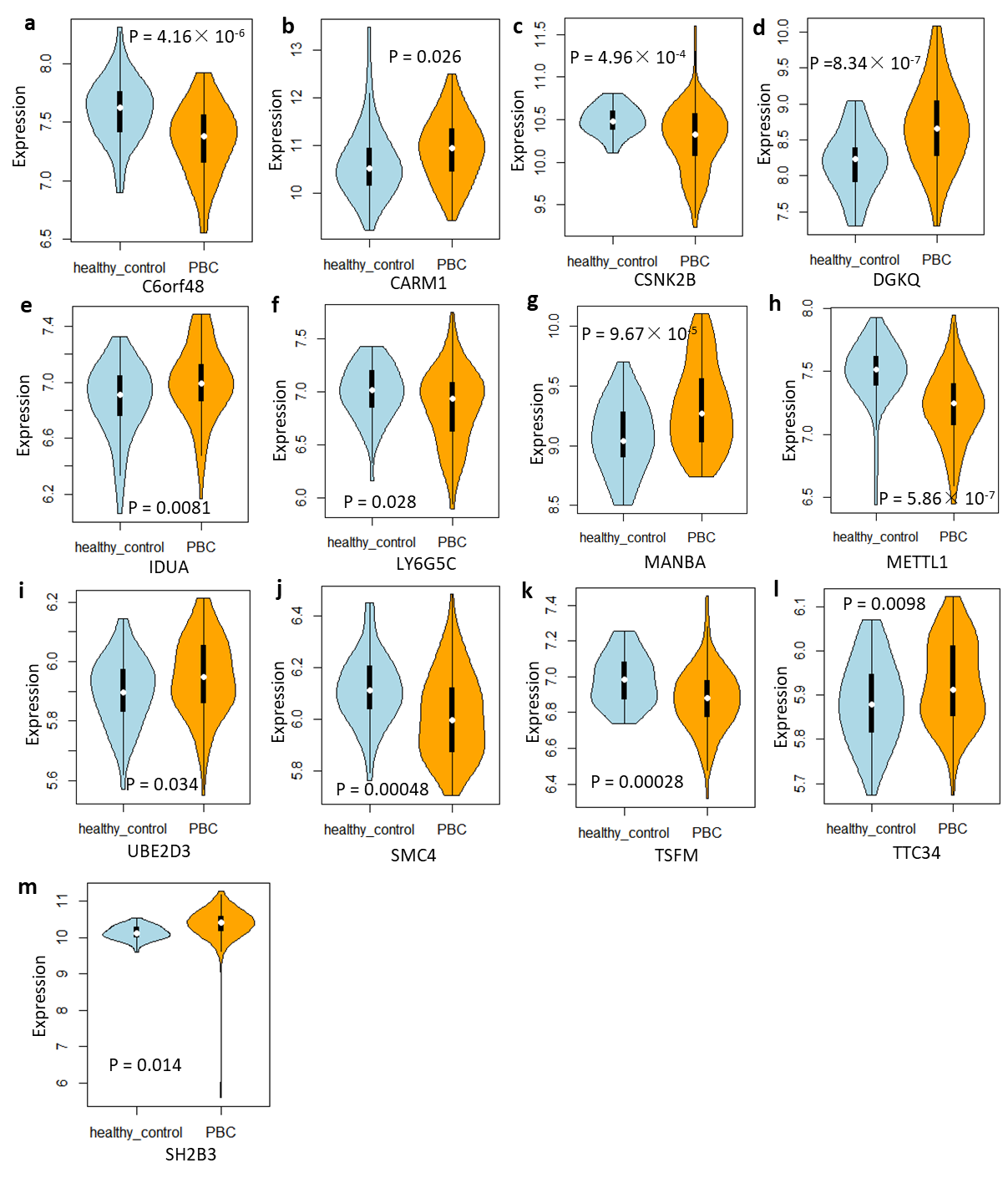

**Supplemental Figure S12. Differential gene expression analysis of 29 risk genes based on bulk RNA profiles in CD4+T cells (Dataset of GSE93170).** a) Results of co-expression patterns among 29 genes in CD4+T cells between control group and PBC group. b)-i) Boxplot showing the significantly differential expression of genes in CD4+T cells between healthy control and PBC patients. b) CSNK2B, c) DDAH2, d) KPNA4, e) SOCS1, f) TSFM, g)SYNGR1, h) UBE2D3, i) ZCRB1.

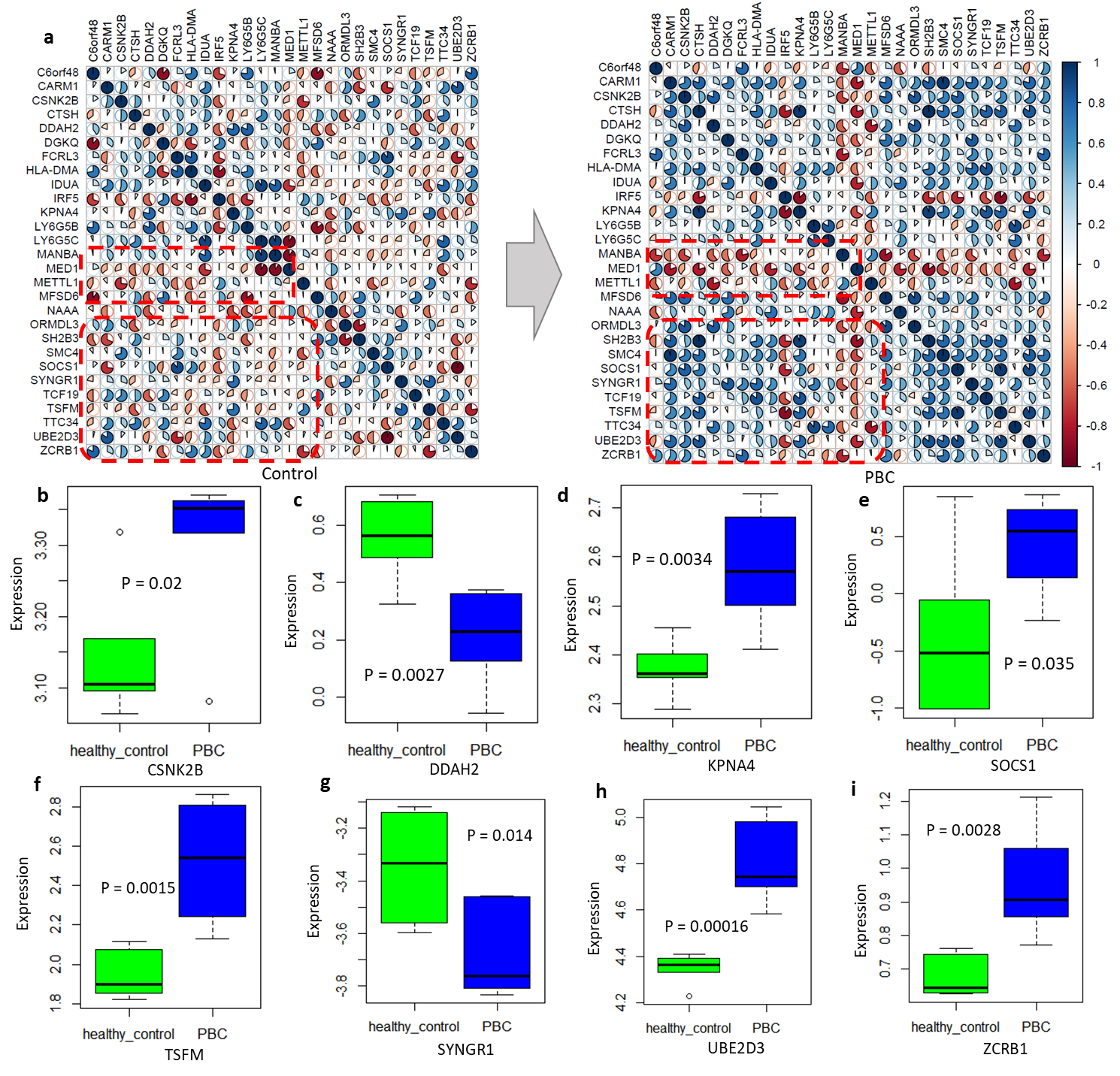

**Supplemental Figure S13. Gene-drug interaction analysis for these identified PBC-associated risk genes.** Note: Orange ring represents a specific gene, and blue triangle represents a given drug. The drug-gene interactions were plotted on the basis of the DGIdb database (<https://www.dgidb.org/>).

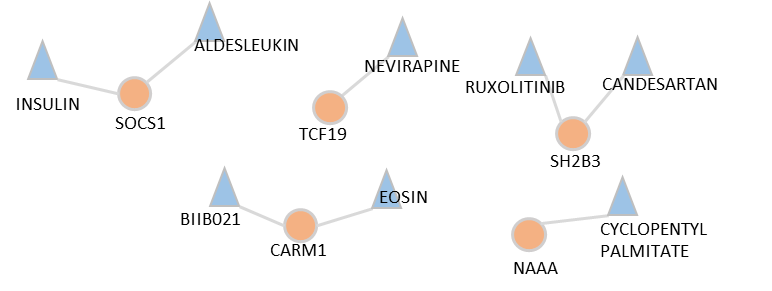

**Supplemental Figure S14.** **Box plots showing the effects of different genotypes (CC, CT, and TT) of rs9303277 on the expression of *ORMDL3* among different immune cell types.** This dataset was based on the Database of Immune Cell Expression quantitative trait loci and Epigenomics (DICE, <https://dice-database.org/landing>)

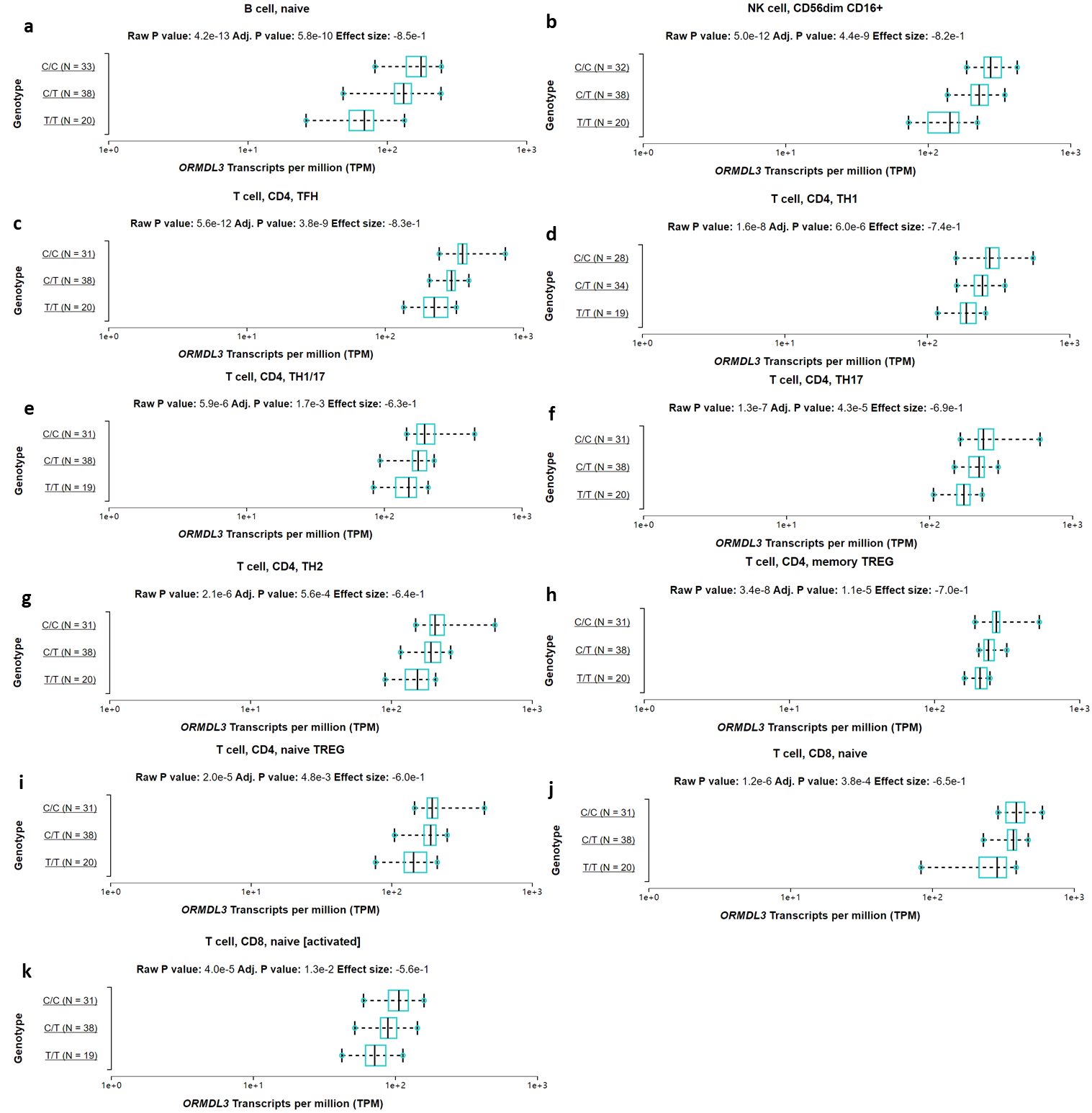

**Supplemental Figure S15. Scatter plot showing the disease-term enrichment analysis for 71 significantly down-regulated genes among *ORMLD3^+^* cholangiocytes based on the Disgenet database.** The interesting input list contains 71 genes in which 70 genes are unambiguously mapped to 70 unique entrez gene IDs and one gene can not be mapped to any entrez gene ID. Note: There were six significant disease-terms overrepresented (FDR < 0.05, see Supplemental Table S19).

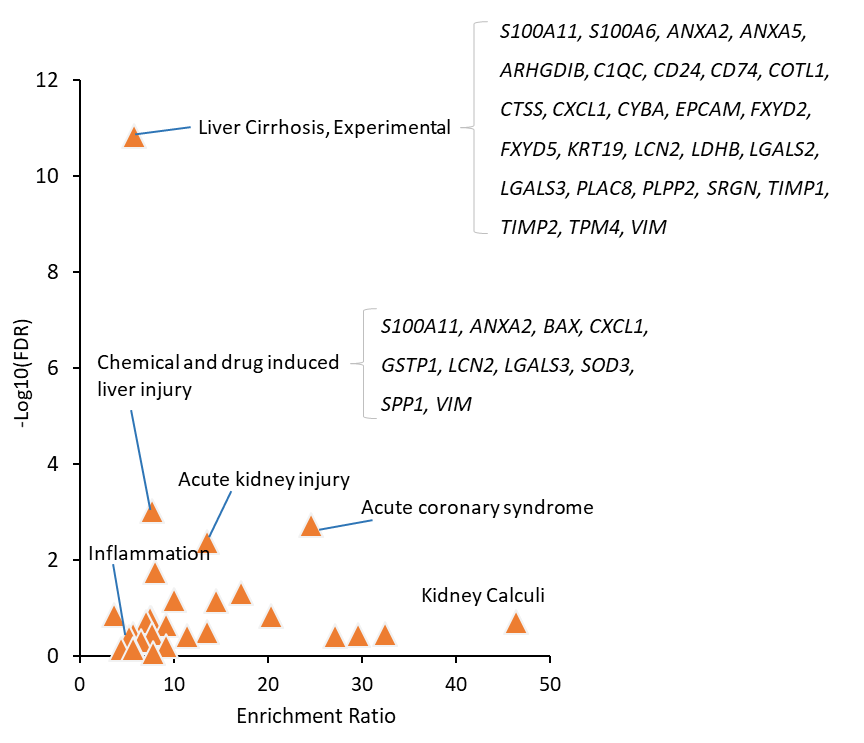
