## Supplemental Tables for "Combination of single cell sequencing data and GWAS summary statistics reveals genetically-influenced liver cell types for primary biliary cholangitis"

**Supplemental Table S1. Summary of collected datasets in current integrative genomics analysis**

| **Characteristics** | **Sample size** | **Description** | **Database resource (URL)** |
| --- | --- | --- | --- |
| GWAS summary data | 13,239 | This dataset was used for extracting genetic information and statistical values. All samples was derived from European ancestry. | This dataset was downloaded from the IEU open GWAS project (<https://gwas.mrcieu.ac.uk/>) |
| eQTL data | 838 | The expression quantitative trait loci datasets were based on 49 tissues from GTEx database (version 8; dbGaP Accession phs000424.v8.p2). The genotype data used for eQTL analysis were on the basis of genome-wide sequencing from 838 donors, which all had RNA-seq data. | The GTEx eQTL datasets Version 8 were downloaded from Zenodo repository (<https://zenodo.org/record/3518299#.Xv6Z6igzbgl>). |
| Bulk-based expression profiles dataset #1 | 9 | For the liver dataset of GSE159676, there were six healthy controls and three PBC cases based on fresh frozen tissue obtained from explanted livers or diagnostic liver biopsies. The Affymetrix Human Gene 1.0 st array was leveraged to obtain bulk expression profiles of 17,046 probes. | <https://www.ncbi.nlm.nih.gov/geo/query/acc.cgi?acc=GSE159676> |
| Bulk-based expression profiles dataset #2 | 137 | With respect to the blood dataset of GSE119600, bulk-based expression profiles were performed using RNA isolated from whole blood samples from 47 healthy controls and 90 PBC patients. The Illumina HumanHT-12 V4.0 expression beadchip was leveraged to obtain blood transcriptome of 47,230 probes. | <https://www.ncbi.nlm.nih.gov/geo/query/acc.cgi?acc=GSE119600> |
| Single cell-based expression profiles dataset #1 | 12 | With regard to the dataset of GSE93170, there were clinically and pathologically diagnosed six healthy controls and six PBC patients enrolled with written informed consent. Peripheral CD4+T cells were used to extract total RNA. The Agilent microarray of SurePrint G3 human GE 8×60K microarray kit was leveraged to produce gene expression profiles according to manufacturer’s protocols. | <https://www.ncbi.nlm.nih.gov/geo/query/acc.cgi?acc=GSE93170> |
| Single cell-based expression profiles dataset #2 | 5 | As for the GSE115469 dataset, five samples from primary liver patients were used for single cell RNA sequencing based on the 10× Genomics Chromium Single Cell Kits. A total of 8,444 parenchymal and non-parenchymal cells have obtained the transcriptional profiles based on the CellRanger analysis pipeline. | <https://www.ncbi.nlm.nih.gov/geo/query/acc.cgi?acc=GSE115469> |

**Supplemental Table S2. Significant genes identified from MAGMA-based gene association analysis (FDR < 0.05)**

| **Gene** | **CHR** | **Number of SNPs** | **Z score** | **P value** | **FDR** |
| --- | --- | --- | --- | --- | --- |
| *HLA-DPA1* | 6 | 104 | 11.02 | 1.49E-28 | 2.69E-24 |
| *IL12A* | 3 | 36 | 10.45 | 7.34E-26 | 6.63E-22 |
| *HLA-DPB1* | 6 | 104 | 10.36 | 1.85E-25 | 1.11E-21 |
| *LOC101929163* | 6 | 87 | 9.82 | 4.68E-23 | 2.11E-19 |
| *BTNL2* | 6 | 101 | 9.35 | 4.56E-21 | 1.65E-17 |
| *RXRB* | 6 | 32 | 9.15 | 2.95E-20 | 8.88E-17 |
| *SLC39A7* | 6 | 25 | 8.62 | 3.42E-18 | 8.83E-15 |
| *HSD17B8* | 6 | 25 | 8.54 | 6.81E-18 | 1.54E-14 |
| *HLA-DQA1* | 6 | 23 | 8.34 | 3.75E-17 | 7.53E-14 |
| *HLA-DRB1* | 6 | 15 | 8.14 | 1.94E-16 | 3.51E-13 |
| *HLA-DQB1* | 6 | 18 | 8.06 | 3.89E-16 | 6.39E-13 |
| *COL11A2* | 6 | 73 | 7.85 | 2.16E-15 | 3.25E-12 |
| *RING1* | 6 | 27 | 7.78 | 3.57E-15 | 4.96E-12 |
| *LY6G5B* | 6 | 52 | 7.66 | 9.23E-15 | 1.19E-11 |
| *LY6G5C* | 6 | 51 | 7.63 | 1.17E-14 | 1.41E-11 |
| *CSNK2B* | 6 | 57 | 7.60 | 1.48E-14 | 1.67E-11 |
| *ABHD16A* | 6 | 56 | 7.53 | 2.53E-14 | 2.69E-11 |
| *GPANK1* | 6 | 62 | 7.45 | 4.65E-14 | 4.67E-11 |
| *C6orf48* | 6 | 20 | 7.44 | 5.08E-14 | 4.83E-11 |
| *APOM* | 6 | 70 | 7.40 | 6.82E-14 | 6.16E-11 |
| *C6orf47* | 6 | 61 | 7.39 | 7.22E-14 | 6.21E-11 |
| *IRF5* | 7 | 17 | 7.37 | 8.74E-14 | 7.18E-11 |
| *LY6G6F* | 6 | 40 | 7.28 | 1.72E-13 | 1.35E-10 |
| *TAP2* | 6 | 151 | 7.25 | 2.11E-13 | 1.59E-10 |
| *AIF1* | 6 | 46 | 7.21 | 2.83E-13 | 2.05E-10 |
| *BAG6* | 6 | 78 | 7.16 | 4.07E-13 | 2.83E-10 |
| *IL12RB2* | 1 | 47 | 7.14 | 4.53E-13 | 3.03E-10 |
| *VARS* | 6 | 37 | 7.09 | 6.63E-13 | 4.28E-10 |
| *C6orf10* | 6 | 189 | 7.04 | 9.47E-13 | 5.58E-10 |
| *HSPA1B* | 6 | 25 | 7.04 | 9.51E-13 | 5.58E-10 |
| *HLA-DRA* | 6 | 102 | 7.04 | 9.57E-13 | 5.58E-10 |
| *CLEC16A* | 16 | 150 | 7.01 | 1.20E-12 | 6.78E-10 |
| *PRRC2A* | 6 | 68 | 7.00 | 1.27E-12 | 6.95E-10 |
| *NEU1* | 6 | 35 | 6.99 | 1.38E-12 | 7.33E-10 |
| *LSM2* | 6 | 22 | 6.88 | 3.05E-12 | 1.57E-09 |
| *DDAH2* | 6 | 39 | 6.86 | 3.36E-12 | 1.69E-09 |
| *TNPO3* | 7 | 42 | 6.85 | 3.66E-12 | 1.79E-09 |
| *HLA-DMB* | 6 | 59 | 6.83 | 4.17E-12 | 1.98E-09 |
| *SCHIP1* | 3 | 188 | 6.79 | 5.65E-12 | 2.62E-09 |
| *EXOC3L4* | 14 | 10 | 6.76 | 6.71E-12 | 2.96E-09 |
| *VWA7* | 6 | 34 | 6.76 | 6.72E-12 | 2.96E-09 |
| *LY6G6C* | 6 | 35 | 6.73 | 8.73E-12 | 3.76E-09 |
| *IQCJ-SCHIP1* | 3 | 267 | 6.71 | 9.85E-12 | 4.14E-09 |
| *CLIC1* | 6 | 40 | 6.69 | 1.08E-11 | 4.42E-09 |
| *NAB1* | 2 | 26 | 6.69 | 1.10E-11 | 4.42E-09 |
| *HLA-DOB* | 6 | 122 | 6.67 | 1.26E-11 | 4.95E-09 |
| *STAT4* | 2 | 59 | 6.64 | 1.54E-11 | 5.92E-09 |
| *SLC44A4* | 6 | 44 | 6.63 | 1.64E-11 | 6.06E-09 |
| *C6orf25* | 6 | 38 | 6.63 | 1.67E-11 | 6.06E-09 |
| *PSMB8* | 6 | 108 | 6.63 | 1.68E-11 | 6.06E-09 |
| *LY6G6D* | 6 | 32 | 6.63 | 1.71E-11 | 6.06E-09 |
| *SCNN1A* | 12 | 23 | 6.62 | 1.83E-11 | 6.36E-09 |
| *MSH5* | 6 | 50 | 6.60 | 2.00E-11 | 6.73E-09 |
| *DDX6* | 11 | 24 | 6.60 | 2.01E-11 | 6.73E-09 |
| *SAPCD1* | 6 | 35 | 6.58 | 2.43E-11 | 7.98E-09 |
| *SOCS1* | 16 | 21 | 6.53 | 3.28E-11 | 1.06E-08 |
| *HSPA1L* | 6 | 24 | 6.50 | 4.09E-11 | 1.30E-08 |
| *PRRT1* | 6 | 31 | 6.49 | 4.25E-11 | 1.32E-08 |
| *HLA-DMA* | 6 | 63 | 6.46 | 5.35E-11 | 1.62E-08 |
| *NFKB1* | 4 | 64 | 6.46 | 5.37E-11 | 1.62E-08 |
| *NCR3* | 6 | 35 | 6.42 | 6.83E-11 | 2.02E-08 |
| *TNP2* | 16 | 26 | 6.40 | 7.85E-11 | 2.29E-08 |
| *EGFL8* | 6 | 35 | 6.34 | 1.17E-10 | 3.36E-08 |
| *ZBTB12* | 6 | 25 | 6.29 | 1.57E-10 | 4.43E-08 |
| *PRM3* | 16 | 29 | 6.27 | 1.80E-10 | 4.96E-08 |
| *SPIB* | 19 | 18 | 6.27 | 1.81E-10 | 4.96E-08 |
| *LTA* | 6 | 37 | 6.25 | 2.00E-10 | 5.39E-08 |
| *CXCR5* | 11 | 17 | 6.22 | 2.56E-10 | 6.80E-08 |
| *MANBA* | 4 | 52 | 6.18 | 3.17E-10 | 8.30E-08 |
| *PRM2* | 16 | 28 | 6.13 | 4.41E-10 | 1.14E-07 |
| *TNF* | 6 | 36 | 6.11 | 4.89E-10 | 1.24E-07 |
| *PPT2* | 6 | 39 | 6.10 | 5.28E-10 | 1.32E-07 |
| *LST1* | 6 | 35 | 6.06 | 6.68E-10 | 1.65E-07 |
| *LTB* | 6 | 37 | 6.02 | 8.76E-10 | 2.12E-07 |
| *ZPBP2* | 17 | 9 | 6.02 | 8.78E-10 | 2.12E-07 |
| *AGPAT1* | 6 | 42 | 6.00 | 9.90E-10 | 2.35E-07 |
| *EHMT2* | 6 | 43 | 5.97 | 1.16E-09 | 2.72E-07 |
| *SERBP1* | 1 | 25 | 5.97 | 1.20E-09 | 2.78E-07 |
| *C2* | 6 | 73 | 5.95 | 1.35E-09 | 3.09E-07 |
| *HLA-B* | 6 | 62 | 5.93 | 1.55E-09 | 3.50E-07 |
| *MICB* | 6 | 86 | 5.92 | 1.57E-09 | 3.50E-07 |
| *ATF6B* | 6 | 39 | 5.88 | 2.06E-09 | 4.54E-07 |
| *TAP1* | 6 | 110 | 5.84 | 2.68E-09 | 5.83E-07 |
| *NOTCH4* | 6 | 109 | 5.83 | 2.85E-09 | 6.13E-07 |
| *RPL3* | 22 | 22 | 5.81 | 3.18E-09 | 6.76E-07 |
| *RPS6KA4* | 11 | 21 | 5.77 | 3.94E-09 | 8.27E-07 |
| *BRD2* | 6 | 71 | 5.77 | 3.98E-09 | 8.27E-07 |
| *POLD1* | 19 | 21 | 5.71 | 5.63E-09 | 1.16E-06 |
| *TNXB* | 6 | 52 | 5.68 | 6.83E-09 | 1.39E-06 |
| *ARHGAP31* | 3 | 101 | 5.65 | 8.05E-09 | 1.62E-06 |
| *HLA-DOA* | 6 | 100 | 5.63 | 9.01E-09 | 1.79E-06 |
| *NFKBIL1* | 6 | 71 | 5.62 | 9.71E-09 | 1.91E-06 |
| *GSDMB* | 17 | 23 | 5.61 | 9.95E-09 | 1.93E-06 |
| *GRB7* | 17 | 9 | 5.59 | 1.11E-08 | 2.13E-06 |
| *HLA-DRB5* | 6 | 1 | 5.59 | 1.15E-08 | 2.19E-06 |
| *ORMDL3* | 17 | 23 | 5.57 | 1.24E-08 | 2.33E-06 |
| *GPSM3* | 6 | 42 | 5.56 | 1.32E-08 | 2.43E-06 |
| *ARL14* | 3 | 16 | 5.56 | 1.32E-08 | 2.43E-06 |
| *LOC101928947* | 17 | 22 | 5.54 | 1.53E-08 | 2.79E-06 |
| *CCDC88B* | 11 | 20 | 5.50 | 1.86E-08 | 3.36E-06 |
| *FKBPL* | 6 | 29 | 5.50 | 1.93E-08 | 3.45E-06 |
| *HSPA1A* | 6 | 20 | 5.46 | 2.41E-08 | 4.27E-06 |
| *ICAM3* | 19 | 21 | 5.44 | 2.63E-08 | 4.61E-06 |
| *TNFAIP2* | 14 | 12 | 5.44 | 2.73E-08 | 4.74E-06 |
| *HLA-C* | 6 | 87 | 5.42 | 2.97E-08 | 5.11E-06 |
| *DDX39B* | 6 | 80 | 5.41 | 3.21E-08 | 5.47E-06 |
| *CAPSL* | 5 | 45 | 5.39 | 3.60E-08 | 6.08E-06 |
| *TMEM39A* | 3 | 32 | 5.36 | 4.08E-08 | 6.83E-06 |
| *CCL20* | 2 | 12 | 5.36 | 4.15E-08 | 6.88E-06 |
| *IL7R* | 5 | 35 | 5.33 | 5.01E-08 | 8.23E-06 |
| *FDX1L* | 19 | 20 | 5.32 | 5.25E-08 | 8.55E-06 |
| *PRDX5* | 11 | 12 | 5.26 | 7.20E-08 | 1.16E-05 |
| *PRM1* | 16 | 28 | 5.23 | 8.38E-08 | 1.34E-05 |
| *MCCD1* | 6 | 70 | 5.22 | 8.99E-08 | 1.42E-05 |
| *LTBR* | 12 | 14 | 5.22 | 9.15E-08 | 1.44E-05 |
| *HIST1H4J* | 6 | 18 | 5.20 | 9.79E-08 | 1.52E-05 |
| *PLCL2* | 3 | 77 | 5.19 | 1.08E-07 | 1.67E-05 |
| *LBH* | 2 | 34 | 5.18 | 1.10E-07 | 1.68E-05 |
| *ATP6V1G2* | 6 | 61 | 5.18 | 1.12E-07 | 1.70E-05 |
| *IKZF3* | 17 | 28 | 5.17 | 1.14E-07 | 1.72E-05 |
| *HIST1H2AJ* | 6 | 14 | 5.16 | 1.21E-07 | 1.79E-05 |
| *HIST1H2BM* | 6 | 14 | 5.16 | 1.21E-07 | 1.79E-05 |
| *DEXI* | 16 | 17 | 5.15 | 1.30E-07 | 1.91E-05 |
| *HIST1H4K* | 6 | 20 | 5.15 | 1.34E-07 | 1.95E-05 |
| *AGER* | 6 | 41 | 5.14 | 1.37E-07 | 1.98E-05 |
| *RMI2* | 16 | 24 | 5.14 | 1.39E-07 | 1.99E-05 |
| *RAVER1* | 19 | 25 | 5.12 | 1.50E-07 | 2.13E-05 |
| *CTSH* | 15 | 25 | 5.12 | 1.55E-07 | 2.19E-05 |
| *HIST1H3H* | 6 | 15 | 5.11 | 1.60E-07 | 2.24E-05 |
| *POGLUT1* | 3 | 26 | 5.09 | 1.75E-07 | 2.43E-05 |
| *HIST1H2AK* | 6 | 18 | 5.08 | 1.86E-07 | 2.57E-05 |
| *PBX2* | 6 | 43 | 5.07 | 1.98E-07 | 2.71E-05 |
| *HIST1H2BL* | 6 | 14 | 5.05 | 2.16E-07 | 2.91E-05 |
| *HIST1H2AI* | 6 | 14 | 5.05 | 2.16E-07 | 2.91E-05 |
| *PSORS1C1* | 6 | 172 | 5.04 | 2.27E-07 | 3.04E-05 |
| *HIST1H2BN* | 6 | 25 | 5.03 | 2.43E-07 | 3.23E-05 |
| *PSMB9* | 6 | 85 | 5.01 | 2.67E-07 | 3.52E-05 |
| *PLEKHG6* | 12 | 18 | 5.01 | 2.69E-07 | 3.52E-05 |
| *TNFRSF1A* | 12 | 17 | 5.01 | 2.77E-07 | 3.60E-05 |
| *ICAM5* | 19 | 20 | 4.99 | 3.10E-07 | 4.00E-05 |
| *RNF5* | 6 | 35 | 4.97 | 3.32E-07 | 4.25E-05 |
| *MED1* | 17 | 18 | 4.96 | 3.47E-07 | 4.42E-05 |
| *TYK2* | 19 | 28 | 4.90 | 4.86E-07 | 6.14E-05 |
| *ZGLP1* | 19 | 22 | 4.90 | 4.91E-07 | 6.16E-05 |
| *MYBPC2* | 19 | 34 | 4.88 | 5.27E-07 | 6.57E-05 |
| *OR5V1* | 6 | 36 | 4.86 | 5.96E-07 | 7.38E-05 |
| *HIST1H1E* | 6 | 17 | 4.85 | 6.15E-07 | 7.56E-05 |
| *HIST1H2BD* | 6 | 26 | 4.84 | 6.49E-07 | 7.92E-05 |
| *STARD3* | 17 | 40 | 4.81 | 7.55E-07 | 9.16E-05 |
| *SLC26A1* | 4 | 26 | 4.79 | 8.25E-07 | 9.94E-05 |
| *OR2H1* | 6 | 28 | 4.79 | 8.33E-07 | 9.97E-05 |
| *LRRC3C* | 17 | 14 | 4.78 | 8.84E-07 | 1.05E-04 |
| *IFT80* | 3 | 38 | 4.77 | 9.04E-07 | 1.07E-04 |
| *C19orf52* | 19 | 10 | 4.75 | 1.02E-06 | 1.20E-04 |
| *FBXL20* | 17 | 54 | 4.75 | 1.03E-06 | 1.20E-04 |
| *CDSN* | 6 | 118 | 4.75 | 1.04E-06 | 1.20E-04 |
| *DXO* | 6 | 41 | 4.71 | 1.21E-06 | 1.39E-04 |
| *HLA-A* | 6 | 22 | 4.71 | 1.24E-06 | 1.41E-04 |
| *YIPF2* | 19 | 12 | 4.71 | 1.25E-06 | 1.41E-04 |
| *PPP1R1B* | 17 | 18 | 4.71 | 1.25E-06 | 1.41E-04 |
| *CRHR1* | 17 | 123 | 4.68 | 1.46E-06 | 1.62E-04 |
| *ESRRA* | 11 | 7 | 4.68 | 1.46E-06 | 1.62E-04 |
| *TRMT112* | 11 | 7 | 4.68 | 1.46E-06 | 1.62E-04 |
| *C6orf15* | 6 | 99 | 4.67 | 1.49E-06 | 1.64E-04 |
| *TRIM31* | 6 | 70 | 4.67 | 1.52E-06 | 1.66E-04 |
| *STK19* | 6 | 39 | 4.67 | 1.54E-06 | 1.68E-04 |
| *IL12B* | 5 | 26 | 4.65 | 1.69E-06 | 1.83E-04 |
| *POU5F1* | 6 | 101 | 4.64 | 1.72E-06 | 1.85E-04 |
| *CARM1* | 19 | 21 | 4.64 | 1.74E-06 | 1.86E-04 |
| *CCHCR1* | 6 | 153 | 4.64 | 1.75E-06 | 1.86E-04 |
| *SLC9B1* | 4 | 44 | 4.63 | 1.84E-06 | 1.94E-04 |
| *SPPL2C* | 17 | 30 | 4.62 | 1.90E-06 | 1.98E-04 |
| *TCF19* | 6 | 117 | 4.62 | 1.90E-06 | 1.98E-04 |
| *TIMMDC1* | 3 | 39 | 4.62 | 1.94E-06 | 2.01E-04 |
| *MICA* | 6 | 65 | 4.62 | 1.96E-06 | 2.02E-04 |
| *SMC4* | 3 | 24 | 4.61 | 2.02E-06 | 2.07E-04 |
| *BAD* | 11 | 17 | 4.60 | 2.14E-06 | 2.18E-04 |
| *DNMT3A* | 2 | 46 | 4.60 | 2.16E-06 | 2.19E-04 |
| *LOC101929578* | 17 | 15 | 4.59 | 2.23E-06 | 2.25E-04 |
| *CDK12* | 17 | 33 | 4.59 | 2.27E-06 | 2.28E-04 |
| *GPR137* | 11 | 12 | 4.58 | 2.35E-06 | 2.35E-04 |
| *DGKQ* | 4 | 27 | 4.58 | 2.37E-06 | 2.35E-04 |
| *HIST1H2AG* | 6 | 12 | 4.57 | 2.38E-06 | 2.35E-04 |
| *ITGB8* | 7 | 85 | 4.56 | 2.55E-06 | 2.50E-04 |
| *TRIM59* | 3 | 16 | 4.51 | 3.22E-06 | 3.14E-04 |
| *NEUROD2* | 17 | 11 | 4.50 | 3.32E-06 | 3.23E-04 |
| *RPL6* | 12 | 3 | 4.50 | 3.40E-06 | 3.29E-04 |
| *OR11A1* | 6 | 46 | 4.47 | 3.89E-06 | 3.74E-04 |
| *BTN3A2* | 6 | 56 | 4.46 | 4.05E-06 | 3.87E-04 |
| *ZSCAN23* | 6 | 32 | 4.45 | 4.22E-06 | 4.01E-04 |
| *MAPT* | 17 | 93 | 4.45 | 4.25E-06 | 4.02E-04 |
| *PPHLN1* | 12 | 51 | 4.45 | 4.28E-06 | 4.03E-04 |
| *ICAM4* | 19 | 16 | 4.44 | 4.48E-06 | 4.19E-04 |
| *PLCB3* | 11 | 28 | 4.44 | 4.52E-06 | 4.21E-04 |
| *CFB* | 6 | 51 | 4.44 | 4.59E-06 | 4.25E-04 |
| *ICAM1* | 19 | 22 | 4.43 | 4.75E-06 | 4.38E-04 |
| *KPNA4* | 3 | 27 | 4.41 | 5.09E-06 | 4.67E-04 |
| *RAD51B* | 14 | 399 | 4.40 | 5.35E-06 | 4.88E-04 |
| *TMEM160* | 19 | 6 | 4.40 | 5.46E-06 | 4.96E-04 |
| *CISD2* | 4 | 21 | 4.40 | 5.53E-06 | 5.00E-04 |
| *HIST1H1B* | 6 | 17 | 4.39 | 5.63E-06 | 5.06E-04 |
| *SKIV2L* | 6 | 54 | 4.38 | 5.85E-06 | 5.23E-04 |
| *STH* | 17 | 24 | 4.37 | 6.17E-06 | 5.44E-04 |
| *DENND1B* | 1 | 89 | 4.37 | 6.19E-06 | 5.44E-04 |
| *HIST1H2AL* | 6 | 19 | 4.37 | 6.20E-06 | 5.44E-04 |
| *NINJ2* | 12 | 85 | 4.37 | 6.20E-06 | 5.44E-04 |
| *UBE2D3* | 4 | 29 | 4.35 | 6.67E-06 | 5.81E-04 |
| *FCRL3* | 1 | 24 | 4.35 | 6.69E-06 | 5.81E-04 |
| *LRRC37A* | 17 | 3 | 4.33 | 7.47E-06 | 6.43E-04 |
| *ARL17B* | 17 | 3 | 4.33 | 7.47E-06 | 6.43E-04 |
| *SLC9B2* | 4 | 38 | 4.32 | 7.72E-06 | 6.61E-04 |
| *TCAP* | 17 | 29 | 4.30 | 8.70E-06 | 7.41E-04 |
| *NAAA* | 4 | 40 | 4.30 | 8.73E-06 | 7.41E-04 |
| *BTN2A1* | 6 | 39 | 4.29 | 9.06E-06 | 7.64E-04 |
| *KANSL1* | 17 | 59 | 4.29 | 9.09E-06 | 7.64E-04 |
| *PGAP3* | 17 | 33 | 4.27 | 9.94E-06 | 8.31E-04 |
| *MUC22* | 6 | 115 | 4.26 | 1.04E-05 | 8.66E-04 |
| *IDUA* | 4 | 26 | 4.25 | 1.08E-05 | 8.95E-04 |
| *MMEL1* | 1 | 31 | 4.24 | 1.10E-05 | 9.08E-04 |
| *FCRL4* | 1 | 45 | 4.24 | 1.11E-05 | 9.12E-04 |
| *LAPTM5* | 1 | 30 | 4.23 | 1.18E-05 | 9.65E-04 |
| *PNMT* | 17 | 31 | 4.22 | 1.20E-05 | 9.72E-04 |
| *CUTA* | 6 | 14 | 4.22 | 1.20E-05 | 9.72E-04 |
| *CD80* | 3 | 53 | 4.19 | 1.40E-05 | 1.13E-03 |
| *C19orf38* | 19 | 14 | 4.16 | 1.60E-05 | 1.28E-03 |
| *TRIM15* | 6 | 80 | 4.16 | 1.60E-05 | 1.28E-03 |
| *NELFE* | 6 | 54 | 4.16 | 1.61E-05 | 1.28E-03 |
| *FAM213B* | 1 | 26 | 4.15 | 1.66E-05 | 1.32E-03 |
| *PHF1* | 6 | 17 | 4.14 | 1.71E-05 | 1.35E-03 |
| *HIST1H2BJ* | 6 | 15 | 4.14 | 1.72E-05 | 1.35E-03 |
| *RPP21* | 6 | 43 | 4.14 | 1.75E-05 | 1.37E-03 |
| *SDAD1* | 4 | 46 | 4.13 | 1.78E-05 | 1.39E-03 |
| *ZC3HAV1* | 7 | 58 | 4.12 | 1.86E-05 | 1.44E-03 |
| *TRAFD1* | 12 | 8 | 4.12 | 1.87E-05 | 1.44E-03 |
| *TRIM39-RPP21* | 6 | 56 | 4.12 | 1.90E-05 | 1.46E-03 |
| *ZCRB1* | 12 | 13 | 4.11 | 1.98E-05 | 1.52E-03 |
| *HIST1H4I* | 6 | 12 | 4.11 | 1.99E-05 | 1.52E-03 |
| *WNT3* | 17 | 35 | 4.11 | 2.00E-05 | 1.52E-03 |
| *NSF* | 17 | 36 | 4.11 | 2.01E-05 | 1.52E-03 |
| *BTN2A2* | 6 | 64 | 4.10 | 2.09E-05 | 1.57E-03 |
| *IFITM3* | 11 | 10 | 4.07 | 2.35E-05 | 1.76E-03 |
| *DUS2* | 16 | 16 | 4.07 | 2.38E-05 | 1.78E-03 |
| *OR10C1* | 6 | 30 | 4.05 | 2.56E-05 | 1.90E-03 |
| *ARL14EPL* | 5 | 35 | 4.04 | 2.63E-05 | 1.95E-03 |
| *ERBB2* | 17 | 26 | 4.03 | 2.79E-05 | 2.05E-03 |
| *DDX28* | 16 | 7 | 4.03 | 2.79E-05 | 2.05E-03 |
| *MFSD6* | 2 | 49 | 4.03 | 2.82E-05 | 2.06E-03 |
| *TRIM10* | 6 | 93 | 4.02 | 2.92E-05 | 2.13E-03 |
| *HLA-G* | 6 | 87 | 4.00 | 3.12E-05 | 2.25E-03 |
| *KCNK4* | 11 | 12 | 4.00 | 3.12E-05 | 2.25E-03 |
| *IFITM1* | 11 | 10 | 3.99 | 3.24E-05 | 2.33E-03 |
| *ZSCAN31* | 6 | 45 | 3.99 | 3.31E-05 | 2.37E-03 |
| *SH2B3* | 12 | 12 | 3.98 | 3.42E-05 | 2.44E-03 |
| *CD58* | 1 | 20 | 3.98 | 3.44E-05 | 2.45E-03 |
| *UGT3A1* | 5 | 73 | 3.96 | 3.74E-05 | 2.65E-03 |
| *ATXN2* | 12 | 29 | 3.96 | 3.77E-05 | 2.66E-03 |
| *ZC3H4* | 19 | 18 | 3.95 | 3.84E-05 | 2.69E-03 |
| *PLEKHM1* | 17 | 7 | 3.95 | 3.84E-05 | 2.69E-03 |
| *ZC3HAV1L* | 7 | 26 | 3.94 | 3.99E-05 | 2.78E-03 |
| *TRIM39* | 6 | 53 | 3.94 | 4.14E-05 | 2.88E-03 |
| *BAHCC1* | 17 | 13 | 3.93 | 4.16E-05 | 2.88E-03 |
| *DPEP2* | 16 | 10 | 3.93 | 4.25E-05 | 2.93E-03 |
| *UVSSA* | 4 | 31 | 3.93 | 4.28E-05 | 2.94E-03 |
| *C4A* | 6 | 23 | 3.92 | 4.35E-05 | 2.98E-03 |
| *LOC554223* | 6 | 82 | 3.92 | 4.40E-05 | 3.00E-03 |
| *OR14J1* | 6 | 27 | 3.92 | 4.46E-05 | 3.02E-03 |
| *RARB* | 3 | 304 | 3.92 | 4.46E-05 | 3.02E-03 |
| *SIRPG* | 20 | 47 | 3.92 | 4.48E-05 | 3.02E-03 |
| *TRIM40* | 6 | 90 | 3.92 | 4.51E-05 | 3.02E-03 |
| *TRAF3* | 14 | 46 | 3.92 | 4.52E-05 | 3.02E-03 |
| *KIFC1* | 6 | 21 | 3.91 | 4.69E-05 | 3.13E-03 |
| *CRIPAK* | 4 | 10 | 3.90 | 4.84E-05 | 3.22E-03 |
| *RGL2* | 6 | 35 | 3.89 | 4.98E-05 | 3.30E-03 |
| *ZKSCAN3* | 6 | 42 | 3.89 | 5.07E-05 | 3.34E-03 |
| *LOC101928548* | 4 | 7 | 3.89 | 5.09E-05 | 3.34E-03 |
| *C5orf30* | 5 | 14 | 3.88 | 5.15E-05 | 3.37E-03 |
| *CXCL9* | 4 | 31 | 3.88 | 5.20E-05 | 3.39E-03 |
| *PATE2* | 11 | 24 | 3.88 | 5.27E-05 | 3.42E-03 |
| *TMED1* | 19 | 8 | 3.88 | 5.28E-05 | 3.42E-03 |
| *TMEM175* | 4 | 26 | 3.87 | 5.39E-05 | 3.48E-03 |
| *ALDH2* | 12 | 18 | 3.85 | 5.80E-05 | 3.73E-03 |
| *PHLDB1* | 11 | 23 | 3.85 | 5.84E-05 | 3.74E-03 |
| *BRAP* | 12 | 19 | 3.85 | 5.89E-05 | 3.74E-03 |
| *NFATC3* | 16 | 38 | 3.85 | 5.89E-05 | 3.74E-03 |
| *TAPBP* | 6 | 30 | 3.85 | 5.92E-05 | 3.74E-03 |
| *ZNF771* | 16 | 11 | 3.85 | 5.92E-05 | 3.74E-03 |
| *FCRL5* | 1 | 53 | 3.84 | 6.25E-05 | 3.91E-03 |
| *MAEA* | 4 | 38 | 3.84 | 6.25E-05 | 3.91E-03 |
| *FGFRL1* | 4 | 22 | 3.84 | 6.26E-05 | 3.91E-03 |
| *ZKSCAN4* | 6 | 37 | 3.82 | 6.63E-05 | 4.13E-03 |
| *C3orf80* | 3 | 10 | 3.82 | 6.68E-05 | 4.15E-03 |
| *HIST1H2BK* | 6 | 13 | 3.82 | 6.77E-05 | 4.19E-03 |
| *PFDN6* | 6 | 29 | 3.82 | 6.79E-05 | 4.19E-03 |
| *PGBD1* | 6 | 35 | 3.81 | 7.09E-05 | 4.35E-03 |
| *NPAS1* | 19 | 13 | 3.80 | 7.11E-05 | 4.35E-03 |
| *PFKFB3* | 10 | 93 | 3.80 | 7.14E-05 | 4.36E-03 |
| *POU2AF1* | 11 | 37 | 3.79 | 7.47E-05 | 4.54E-03 |
| *ABT1* | 6 | 24 | 3.78 | 7.97E-05 | 4.83E-03 |
| *MIEN1* | 17 | 4 | 3.77 | 8.11E-05 | 4.90E-03 |
| *TTC34* | 1 | 7 | 3.77 | 8.27E-05 | 4.98E-03 |
| *DHX16* | 6 | 29 | 3.75 | 8.87E-05 | 5.32E-03 |
| *UBD* | 6 | 51 | 3.74 | 9.12E-05 | 5.45E-03 |
| *LOC101928621* | 4 | 21 | 3.74 | 9.14E-05 | 5.45E-03 |
| *PATE1* | 11 | 29 | 3.74 | 9.25E-05 | 5.50E-03 |
| *EFR3B* | 2 | 55 | 3.74 | 9.33E-05 | 5.53E-03 |
| *ACTL7A* | 9 | 53 | 3.74 | 9.38E-05 | 5.54E-03 |
| *ZSCAN12* | 6 | 46 | 3.73 | 9.66E-05 | 5.68E-03 |
| *IL15RA* | 10 | 58 | 3.73 | 9.68E-05 | 5.68E-03 |
| *ID3* | 1 | 14 | 3.72 | 9.97E-05 | 5.83E-03 |
| *PDE4A* | 19 | 20 | 3.72 | 1.00E-04 | 5.83E-03 |
| *CXCL10* | 4 | 38 | 3.71 | 1.05E-04 | 6.09E-03 |
| *ACTG1* | 17 | 20 | 3.70 | 1.10E-04 | 6.35E-03 |
| *TM4SF4* | 3 | 55 | 3.69 | 1.12E-04 | 6.46E-03 |
| *ACTL7B* | 9 | 53 | 3.69 | 1.13E-04 | 6.49E-03 |
| *CRB1* | 1 | 77 | 3.69 | 1.13E-04 | 6.49E-03 |
| *GSDMA* | 17 | 16 | 3.68 | 1.17E-04 | 6.68E-03 |
| *FAM109A* | 12 | 19 | 3.67 | 1.19E-04 | 6.81E-03 |
| *NFKB2* | 10 | 12 | 3.67 | 1.21E-04 | 6.86E-03 |
| *NKAPL* | 6 | 30 | 3.67 | 1.22E-04 | 6.89E-03 |
| *MYOZ1* | 10 | 13 | 3.67 | 1.22E-04 | 6.91E-03 |
| *DPEP3* | 16 | 8 | 3.67 | 1.23E-04 | 6.91E-03 |
| *HMCN1* | 1 | 108 | 3.66 | 1.25E-04 | 7.01E-03 |
| *PTPN11* | 12 | 18 | 3.65 | 1.31E-04 | 7.34E-03 |
| *NAA25* | 12 | 21 | 3.64 | 1.34E-04 | 7.46E-03 |
| *GPR65* | 14 | 20 | 3.64 | 1.36E-04 | 7.53E-03 |
| *WDR46* | 6 | 31 | 3.64 | 1.36E-04 | 7.53E-03 |
| *ODF3B* | 22 | 18 | 3.64 | 1.37E-04 | 7.58E-03 |
| *TNFSF15* | 9 | 28 | 3.63 | 1.41E-04 | 7.74E-03 |
| *GPX5* | 6 | 29 | 3.63 | 1.42E-04 | 7.80E-03 |
| *DAXX* | 6 | 18 | 3.63 | 1.43E-04 | 7.81E-03 |
| *PPP1R18* | 6 | 27 | 3.63 | 1.44E-04 | 7.85E-03 |
| *MAS1L* | 6 | 24 | 3.62 | 1.45E-04 | 7.87E-03 |
| *BAK1* | 6 | 31 | 3.62 | 1.45E-04 | 7.88E-03 |
| *PSORS1C2* | 6 | 118 | 3.62 | 1.47E-04 | 7.95E-03 |
| *IFITM2* | 11 | 9 | 3.62 | 1.48E-04 | 7.97E-03 |
| *DNM2* | 19 | 49 | 3.61 | 1.51E-04 | 8.12E-03 |
| *ORAI2* | 7 | 27 | 3.60 | 1.58E-04 | 8.45E-03 |
| *FAM43B* | 1 | 35 | 3.60 | 1.58E-04 | 8.46E-03 |
| *TXNIP* | 1 | 2 | 3.60 | 1.62E-04 | 8.58E-03 |
| *ZBTB22* | 6 | 17 | 3.60 | 1.62E-04 | 8.58E-03 |
| *GNAI3* | 1 | 38 | 3.60 | 1.62E-04 | 8.58E-03 |
| *ART3* | 4 | 89 | 3.59 | 1.67E-04 | 8.84E-03 |
| *GALC* | 14 | 105 | 3.59 | 1.68E-04 | 8.84E-03 |
| *GNAT2* | 1 | 26 | 3.58 | 1.72E-04 | 9.01E-03 |
| *ASPRV1* | 2 | 17 | 3.58 | 1.72E-04 | 9.01E-03 |
| *TYMP* | 22 | 17 | 3.58 | 1.73E-04 | 9.04E-03 |
| *TEX40* | 11 | 8 | 3.58 | 1.74E-04 | 9.07E-03 |
| *CXCL11* | 4 | 27 | 3.55 | 1.90E-04 | 9.85E-03 |
| *FUT11* | 10 | 19 | 3.55 | 1.94E-04 | 1.01E-02 |
| *AGL* | 1 | 61 | 3.55 | 1.96E-04 | 1.01E-02 |
| *AVIL* | 12 | 20 | 3.54 | 1.99E-04 | 1.03E-02 |
| *RGS9BP* | 19 | 15 | 3.54 | 2.00E-04 | 1.03E-02 |
| *ACAD10* | 12 | 25 | 3.54 | 2.00E-04 | 1.03E-02 |
| *DCTPP1* | 16 | 9 | 3.54 | 2.01E-04 | 1.03E-02 |
| *CDKN2B* | 9 | 26 | 3.54 | 2.02E-04 | 1.03E-02 |
| *ZNF48* | 16 | 19 | 3.53 | 2.05E-04 | 1.04E-02 |
| *ANKRD27* | 19 | 64 | 3.53 | 2.08E-04 | 1.05E-02 |
| *FSCN2* | 17 | 31 | 3.53 | 2.11E-04 | 1.07E-02 |
| *CTSB* | 8 | 46 | 3.52 | 2.14E-04 | 1.08E-02 |
| *NDST2* | 10 | 14 | 3.52 | 2.16E-04 | 1.09E-02 |
| *IL6* | 7 | 41 | 3.52 | 2.18E-04 | 1.09E-02 |
| *HECTD4* | 12 | 31 | 3.51 | 2.22E-04 | 1.11E-02 |
| *PLA2G15* | 16 | 16 | 3.51 | 2.23E-04 | 1.11E-02 |
| *GNRHR* | 4 | 21 | 3.51 | 2.27E-04 | 1.13E-02 |
| *UTS2B* | 3 | 52 | 3.50 | 2.29E-04 | 1.13E-02 |
| *SLC30A7* | 1 | 69 | 3.50 | 2.34E-04 | 1.15E-02 |
| *CDC37* | 19 | 15 | 3.49 | 2.41E-04 | 1.18E-02 |
| *42979* | 16 | 15 | 3.49 | 2.42E-04 | 1.19E-02 |
| *LBR* | 1 | 36 | 3.48 | 2.46E-04 | 1.20E-02 |
| *RIN3* | 14 | 120 | 3.48 | 2.47E-04 | 1.20E-02 |
| *SP2* | 17 | 30 | 3.48 | 2.47E-04 | 1.20E-02 |
| *TET2* | 4 | 58 | 3.47 | 2.59E-04 | 1.26E-02 |
| *TNFRSF11A* | 18 | 65 | 3.47 | 2.65E-04 | 1.28E-02 |
| *ZSCAN26* | 6 | 33 | 3.46 | 2.74E-04 | 1.32E-02 |
| *ARHGAP27* | 17 | 23 | 3.45 | 2.76E-04 | 1.33E-02 |
| *PNPO* | 17 | 23 | 3.45 | 2.78E-04 | 1.34E-02 |
| *42803* | 12 | 17 | 3.45 | 2.83E-04 | 1.35E-02 |
| *CDK4* | 12 | 14 | 3.44 | 2.87E-04 | 1.37E-02 |
| *PPP1R14B* | 11 | 22 | 3.44 | 2.89E-04 | 1.38E-02 |
| *MYLPF* | 16 | 14 | 3.44 | 2.90E-04 | 1.38E-02 |
| *ZNF311* | 6 | 39 | 3.44 | 2.91E-04 | 1.38E-02 |
| *C4B* | 6 | 5 | 3.43 | 2.97E-04 | 1.40E-02 |
| *KATNAL2* | 18 | 61 | 3.43 | 3.02E-04 | 1.42E-02 |
| *ZSWIM8* | 10 | 21 | 3.43 | 3.02E-04 | 1.42E-02 |
| *NUDT19* | 19 | 24 | 3.42 | 3.11E-04 | 1.46E-02 |
| *TMEM88B* | 1 | 1 | 3.42 | 3.12E-04 | 1.46E-02 |
| *VWA1* | 1 | 1 | 3.42 | 3.12E-04 | 1.46E-02 |
| *ACAP3* | 1 | 13 | 3.42 | 3.15E-04 | 1.47E-02 |
| *AGAP2* | 12 | 19 | 3.41 | 3.19E-04 | 1.48E-02 |
| *SIK2* | 11 | 36 | 3.41 | 3.28E-04 | 1.52E-02 |
| *SEC24C* | 10 | 22 | 3.40 | 3.34E-04 | 1.54E-02 |
| *CHCHD1* | 10 | 19 | 3.40 | 3.34E-04 | 1.54E-02 |
| *SCO2* | 22 | 16 | 3.40 | 3.36E-04 | 1.54E-02 |
| *TSFM* | 12 | 25 | 3.40 | 3.37E-04 | 1.54E-02 |
| *MLLT6* | 17 | 15 | 3.40 | 3.38E-04 | 1.54E-02 |
| *SYNPO2L* | 10 | 15 | 3.40 | 3.41E-04 | 1.55E-02 |
| *GTF2H1* | 11 | 28 | 3.39 | 3.44E-04 | 1.57E-02 |
| *ESRP2* | 16 | 14 | 3.39 | 3.45E-04 | 1.57E-02 |
| *TNFAIP3* | 6 | 20 | 3.39 | 3.46E-04 | 1.57E-02 |
| *REST* | 4 | 22 | 3.39 | 3.50E-04 | 1.58E-02 |
| *METTL21B* | 12 | 19 | 3.39 | 3.54E-04 | 1.60E-02 |
| *NCAPH2* | 22 | 21 | 3.38 | 3.56E-04 | 1.60E-02 |
| *TMEM116* | 12 | 26 | 3.38 | 3.62E-04 | 1.62E-02 |
| *IL12RB1* | 19 | 19 | 3.37 | 3.70E-04 | 1.65E-02 |
| *CYP27B1* | 12 | 17 | 3.37 | 3.78E-04 | 1.68E-02 |
| *METTL1* | 12 | 17 | 3.37 | 3.78E-04 | 1.68E-02 |
| *SMARCA4* | 19 | 46 | 3.37 | 3.79E-04 | 1.68E-02 |
| *B3GALT4* | 6 | 25 | 3.37 | 3.83E-04 | 1.69E-02 |
| *PRSS16* | 6 | 19 | 3.36 | 3.97E-04 | 1.75E-02 |
| *RASGRF1* | 15 | 85 | 3.35 | 4.00E-04 | 1.76E-02 |
| *GALNT14* | 2 | 144 | 3.35 | 4.05E-04 | 1.78E-02 |
| *FAM214A* | 15 | 47 | 3.35 | 4.11E-04 | 1.80E-02 |
| *ENAH* | 1 | 67 | 3.34 | 4.16E-04 | 1.82E-02 |
| *TBC1D10B* | 16 | 14 | 3.34 | 4.19E-04 | 1.83E-02 |
| *RHPN2* | 19 | 38 | 3.34 | 4.20E-04 | 1.83E-02 |
| *KLHL8* | 4 | 45 | 3.34 | 4.24E-04 | 1.84E-02 |
| *KLHDC7B* | 22 | 18 | 3.34 | 4.25E-04 | 1.84E-02 |
| *POLR3GL* | 1 | 2 | 3.34 | 4.26E-04 | 1.84E-02 |
| *PSD* | 10 | 20 | 3.34 | 4.26E-04 | 1.84E-02 |
| *HLA-DQB2* | 6 | 99 | 3.33 | 4.31E-04 | 1.85E-02 |
| *RPS18* | 6 | 26 | 3.33 | 4.35E-04 | 1.87E-02 |
| *HIST1H2BE* | 6 | 26 | 3.33 | 4.37E-04 | 1.87E-02 |
| *SYNGAP1* | 6 | 24 | 3.32 | 4.55E-04 | 1.94E-02 |
| *TGFBR2* | 3 | 85 | 3.31 | 4.64E-04 | 1.98E-02 |
| *PRKAG3* | 2 | 9 | 3.31 | 4.71E-04 | 2.00E-02 |
| *ERICH6* | 3 | 31 | 3.30 | 4.77E-04 | 2.02E-02 |
| *ARL6IP4* | 12 | 8 | 3.30 | 4.81E-04 | 2.03E-02 |
| *LNPEP* | 5 | 40 | 3.30 | 4.81E-04 | 2.03E-02 |
| *CTDSP2* | 12 | 23 | 3.30 | 4.82E-04 | 2.03E-02 |
| *YAF2* | 12 | 33 | 3.29 | 4.93E-04 | 2.07E-02 |
| *ALKBH4* | 7 | 19 | 3.29 | 4.95E-04 | 2.08E-02 |
| *HIST1H3I* | 6 | 15 | 3.29 | 5.06E-04 | 2.11E-02 |
| *HIST1H4L* | 6 | 15 | 3.29 | 5.06E-04 | 2.11E-02 |
| *NAPSA* | 19 | 9 | 3.28 | 5.19E-04 | 2.16E-02 |
| *ICOS* | 2 | 36 | 3.28 | 5.20E-04 | 2.16E-02 |
| *AFF1* | 4 | 104 | 3.28 | 5.21E-04 | 2.16E-02 |
| *GPATCH1* | 19 | 33 | 3.28 | 5.25E-04 | 2.16E-02 |
| *MRPL20* | 1 | 3 | 3.28 | 5.26E-04 | 2.16E-02 |
| *ANKRD65* | 1 | 3 | 3.28 | 5.26E-04 | 2.16E-02 |
| *VPS52* | 6 | 32 | 3.27 | 5.30E-04 | 2.18E-02 |
| *POM121L2* | 6 | 21 | 3.27 | 5.35E-04 | 2.19E-02 |
| *FLOT1* | 6 | 38 | 3.27 | 5.42E-04 | 2.21E-02 |
| *DPH5* | 1 | 28 | 3.26 | 5.54E-04 | 2.26E-02 |
| *SKOR2* | 18 | 31 | 3.25 | 5.72E-04 | 2.32E-02 |
| *PATE3* | 11 | 23 | 3.25 | 5.72E-04 | 2.32E-02 |
| *CD2BP2* | 16 | 11 | 3.24 | 5.88E-04 | 2.38E-02 |
| *DRD4* | 11 | 19 | 3.24 | 5.89E-04 | 2.38E-02 |
| *MAPKAPK5* | 12 | 27 | 3.24 | 5.99E-04 | 2.41E-02 |
| *CISD3* | 17 | 16 | 3.24 | 5.99E-04 | 2.41E-02 |
| *IER3* | 6 | 35 | 3.24 | 6.06E-04 | 2.43E-02 |
| *HNRNPK* | 9 | 13 | 3.23 | 6.12E-04 | 2.45E-02 |
| *UBLCP1* | 5 | 24 | 3.22 | 6.35E-04 | 2.54E-02 |
| *WDR88* | 19 | 34 | 3.22 | 6.41E-04 | 2.56E-02 |
| *SF3A1* | 22 | 41 | 3.21 | 6.54E-04 | 2.60E-02 |
| *CDA* | 1 | 63 | 3.21 | 6.56E-04 | 2.61E-02 |
| *TBC1D10A* | 22 | 44 | 3.21 | 6.62E-04 | 2.62E-02 |
| *SLMO1* | 18 | 30 | 3.21 | 6.69E-04 | 2.65E-02 |
| *ZDHHC13* | 11 | 44 | 3.21 | 6.74E-04 | 2.66E-02 |
| *IL24* | 1 | 17 | 3.20 | 6.86E-04 | 2.70E-02 |
| *DAPP1* | 4 | 45 | 3.20 | 6.93E-04 | 2.72E-02 |
| *GCKR* | 2 | 23 | 3.19 | 7.02E-04 | 2.75E-02 |
| *OSTN* | 3 | 54 | 3.19 | 7.12E-04 | 2.78E-02 |
| *RMI1* | 9 | 16 | 3.19 | 7.12E-04 | 2.78E-02 |
| *SCT* | 11 | 23 | 3.19 | 7.23E-04 | 2.81E-02 |
| *CCDC157* | 22 | 35 | 3.18 | 7.34E-04 | 2.85E-02 |
| *OR2F1* | 7 | 14 | 3.17 | 7.54E-04 | 2.92E-02 |
| *TNIP1* | 5 | 93 | 3.17 | 7.55E-04 | 2.92E-02 |
| *CLEC2D* | 12 | 56 | 3.17 | 7.64E-04 | 2.95E-02 |
| *OR2J3* | 6 | 19 | 3.17 | 7.69E-04 | 2.96E-02 |
| *C6orf136* | 6 | 19 | 3.17 | 7.72E-04 | 2.96E-02 |
| *PATE4* | 11 | 6 | 3.17 | 7.73E-04 | 2.96E-02 |
| *OS9* | 12 | 27 | 3.17 | 7.73E-04 | 2.96E-02 |
| *CD83* | 6 | 39 | 3.16 | 7.83E-04 | 2.99E-02 |
| *HIST1H4H* | 6 | 27 | 3.16 | 7.92E-04 | 3.02E-02 |
| *VPS41* | 7 | 117 | 3.16 | 7.96E-04 | 3.03E-02 |
| *TCEB3B* | 18 | 14 | 3.15 | 8.04E-04 | 3.05E-02 |
| *SPON2* | 4 | 23 | 3.15 | 8.04E-04 | 3.05E-02 |
| *PITPNM2* | 12 | 57 | 3.15 | 8.24E-04 | 3.11E-02 |
| *KEAP1* | 19 | 20 | 3.15 | 8.25E-04 | 3.11E-02 |
| *ERAP2* | 5 | 41 | 3.15 | 8.26E-04 | 3.11E-02 |
| *CAMK2G* | 10 | 37 | 3.14 | 8.33E-04 | 3.13E-02 |
| *IRF8* | 16 | 81 | 3.14 | 8.54E-04 | 3.20E-02 |
| *ERAP1* | 5 | 98 | 3.13 | 8.79E-04 | 3.29E-02 |
| *TRIM26* | 6 | 84 | 3.13 | 8.84E-04 | 3.30E-02 |
| *FOXP1* | 3 | 277 | 3.12 | 8.92E-04 | 3.32E-02 |
| *ABO* | 9 | 38 | 3.12 | 9.00E-04 | 3.35E-02 |
| *SPPL3* | 12 | 78 | 3.12 | 9.05E-04 | 3.36E-02 |
| *OAS2* | 12 | 43 | 3.12 | 9.10E-04 | 3.37E-02 |
| *GBF1* | 10 | 56 | 3.11 | 9.21E-04 | 3.40E-02 |
| *CHAF1B* | 21 | 27 | 3.11 | 9.47E-04 | 3.49E-02 |
| *TBC1D23* | 3 | 42 | 3.11 | 9.47E-04 | 3.49E-02 |
| *OR12D3* | 6 | 59 | 3.10 | 9.54E-04 | 3.50E-02 |
| *VEGFB* | 11 | 21 | 3.10 | 9.63E-04 | 3.53E-02 |
| *DHX9* | 1 | 17 | 3.10 | 9.65E-04 | 3.53E-02 |
| *SPINK2* | 4 | 21 | 3.10 | 9.74E-04 | 3.56E-02 |
| *SLC7A6* | 16 | 26 | 3.09 | 9.89E-04 | 3.60E-02 |
| *CDHR5* | 11 | 25 | 3.09 | 9.98E-04 | 3.63E-02 |
| *HYLS1* | 11 | 32 | 3.09 | 1.00E-03 | 3.64E-02 |
| *HSD17B11* | 4 | 24 | 3.09 | 1.01E-03 | 3.65E-02 |
| *AGAP5* | 10 | 6 | 3.09 | 1.01E-03 | 3.65E-02 |
| *PCGF2* | 17 | 21 | 3.09 | 1.01E-03 | 3.65E-02 |
| *HIST1H2BI* | 6 | 25 | 3.09 | 1.02E-03 | 3.65E-02 |
| *LMF2* | 22 | 18 | 3.09 | 1.02E-03 | 3.65E-02 |
| *PDE6H* | 12 | 35 | 3.09 | 1.02E-03 | 3.65E-02 |
| *GFRA1* | 10 | 163 | 3.08 | 1.02E-03 | 3.66E-02 |
| *C17orf70* | 17 | 25 | 3.08 | 1.03E-03 | 3.68E-02 |
| *FKBP2* | 11 | 22 | 3.08 | 1.03E-03 | 3.68E-02 |
| *TSPAN31* | 12 | 12 | 3.08 | 1.04E-03 | 3.69E-02 |
| *CTBP1* | 4 | 30 | 3.07 | 1.06E-03 | 3.76E-02 |
| *DOCK2* | 5 | 344 | 3.07 | 1.08E-03 | 3.81E-02 |
| *FCMR* | 1 | 29 | 3.06 | 1.09E-03 | 3.86E-02 |
| *BFSP1* | 20 | 70 | 3.06 | 1.10E-03 | 3.88E-02 |
| *EXOC2* | 6 | 160 | 3.05 | 1.15E-03 | 4.06E-02 |
| *HIST1H3G* | 6 | 26 | 3.05 | 1.16E-03 | 4.07E-02 |
| *AURKAIP1* | 1 | 5 | 3.05 | 1.16E-03 | 4.07E-02 |
| *CPTP* | 1 | 5 | 3.04 | 1.17E-03 | 4.09E-02 |
| *SLC15A1* | 13 | 92 | 3.04 | 1.17E-03 | 4.09E-02 |
| *EXTL2* | 1 | 23 | 3.04 | 1.18E-03 | 4.11E-02 |
| *TAS1R3* | 1 | 7 | 3.04 | 1.18E-03 | 4.12E-02 |
| *SYNGR1* | 22 | 30 | 3.04 | 1.19E-03 | 4.13E-02 |
| *IFNGR2* | 21 | 39 | 3.04 | 1.19E-03 | 4.13E-02 |
| *OR2B3* | 6 | 15 | 3.03 | 1.21E-03 | 4.20E-02 |
| *CX3CR1* | 3 | 40 | 3.03 | 1.22E-03 | 4.23E-02 |
| *CD226* | 18 | 49 | 3.03 | 1.23E-03 | 4.25E-02 |
| *SLC36A1* | 5 | 60 | 3.02 | 1.24E-03 | 4.28E-02 |
| *ELOVL3* | 10 | 17 | 3.02 | 1.25E-03 | 4.28E-02 |
| *ERP29* | 12 | 15 | 3.02 | 1.25E-03 | 4.28E-02 |
| *GTDC1* | 2 | 94 | 3.02 | 1.25E-03 | 4.28E-02 |
| *HIRA* | 22 | 72 | 3.02 | 1.25E-03 | 4.28E-02 |
| *CPSF3L* | 1 | 8 | 3.02 | 1.25E-03 | 4.28E-02 |
| *FAM131A* | 3 | 18 | 3.02 | 1.26E-03 | 4.28E-02 |
| *GABBR1* | 6 | 59 | 3.02 | 1.26E-03 | 4.29E-02 |
| *HAAO* | 2 | 31 | 3.02 | 1.27E-03 | 4.29E-02 |
| *DVL1* | 1 | 10 | 3.02 | 1.27E-03 | 4.29E-02 |
| *LRWD1* | 7 | 16 | 3.02 | 1.27E-03 | 4.29E-02 |
| *POMC* | 2 | 20 | 3.02 | 1.28E-03 | 4.32E-02 |
| *KRT39* | 17 | 31 | 3.01 | 1.29E-03 | 4.35E-02 |
| *RADIL* | 7 | 37 | 3.01 | 1.30E-03 | 4.37E-02 |
| *TRIM27* | 6 | 33 | 3.01 | 1.31E-03 | 4.38E-02 |
| *KIF21B* | 1 | 42 | 3.01 | 1.31E-03 | 4.38E-02 |
| *CYP27A1* | 2 | 22 | 3.01 | 1.32E-03 | 4.40E-02 |
| *IL20* | 1 | 8 | 3.00 | 1.34E-03 | 4.47E-02 |
| *NFKBIA* | 14 | 24 | 3.00 | 1.35E-03 | 4.49E-02 |
| *GPR61* | 1 | 21 | 3.00 | 1.35E-03 | 4.49E-02 |
| *CD5L* | 1 | 21 | 3.00 | 1.36E-03 | 4.49E-02 |
| *CYP21A2* | 6 | 7 | 2.99 | 1.37E-03 | 4.55E-02 |
| *FDFT1* | 8 | 68 | 2.99 | 1.38E-03 | 4.55E-02 |
| *UBA6* | 4 | 37 | 2.99 | 1.38E-03 | 4.56E-02 |
| *PUSL1* | 1 | 7 | 2.99 | 1.39E-03 | 4.56E-02 |
| *MXRA8* | 1 | 8 | 2.98 | 1.42E-03 | 4.68E-02 |
| *TMEM50B* | 21 | 36 | 2.98 | 1.44E-03 | 4.71E-02 |
| *GNAO1* | 16 | 96 | 2.98 | 1.45E-03 | 4.76E-02 |
| *SPIRE1* | 18 | 74 | 2.98 | 1.46E-03 | 4.77E-02 |
| *MPHOSPH9* | 12 | 39 | 2.98 | 1.46E-03 | 4.77E-02 |
| *FLRT1* | 11 | 49 | 2.97 | 1.47E-03 | 4.78E-02 |
| *GPX6* | 6 | 29 | 2.97 | 1.51E-03 | 4.90E-02 |
| *FSTL3* | 19 | 21 | 2.96 | 1.52E-03 | 4.92E-02 |
| *OAS3* | 12 | 54 | 2.96 | 1.53E-03 | 4.94E-02 |
| *PPP2R1B* | 11 | 24 | 2.96 | 1.55E-03 | 4.99E-02 |
| *TTC39C* | 18 | 81 | 2.96 | 1.55E-03 | 4.99E-02 |
| *TREH* | 11 | 20 | 2.96 | 1.55E-03 | 4.99E-02 |
| *FNDC4* | 2 | 13 | 2.96 | 1.55E-03 | 5.00E-02 |
| *ST8SIA4* | 5 | 47 | 2.96 | 1.56E-03 | 5.00E-02 |

Note: CHR = Chromosome.

**Supplemental Table S3. MAGMA-identified significant enriched pathways based on GWAS summary data**

| **ID** | **Pathway ID** | **Pathway names** | **Beta** | **SE** | **P value** | **FDR** |
| --- | --- | --- | --- | --- | --- | --- |
| 1 | hsa04658 | Th1_and_Th2_cell_differentiation | 0.79 | 0.10 | 9.94E-16 | 3.27E-13 |
| 2 | hsa05330 | Allograft_rejection | 0.92 | 0.16 | 3.15E-09 | 5.18E-07 |
| 3 | hsa05321 | Inflammatory_bowel_disease | 0.65 | 0.12 | 1.66E-08 | 1.82E-06 |
| 4 | hsa04940 | Type_I_diabetes_mellitus | 0.78 | 0.15 | 7.82E-08 | 5.27E-06 |
| 5 | hsa04672 | Intestinal_immune_network_for_IgA_production | 0.75 | 0.14 | 8.01E-08 | 5.27E-06 |
| 6 | hsa04659 | Th17_cell_differentiation | 0.48 | 0.10 | 3.91E-07 | 2.14E-05 |
| 7 | hsa04514 | Cell_adhesion_molecules | 0.40 | 0.08 | 6.51E-07 | 3.06E-05 |
| 8 | hsa05310 | Asthma | 0.89 | 0.19 | 8.53E-07 | 3.51E-05 |
| 9 | hsa04630 | JAK-STAT_signaling_pathway | 0.37 | 0.08 | 1.08E-06 | 3.95E-05 |
| 10 | hsa05323 | Rheumatoid_arthritis | 0.49 | 0.11 | 5.44E-06 | 1.66E-04 |
| 11 | hsa05332 | Graft-versus-host_disease | 0.73 | 0.17 | 5.54E-06 | 1.66E-04 |
| 12 | hsa04060 | Cytokine-cytokine_receptor_interaction | 0.27 | 0.06 | 1.08E-05 | 2.96E-04 |
| 13 | hsa04640 | Hematopoietic_cell_lineage | 0.45 | 0.11 | 1.58E-05 | 3.92E-04 |
| 14 | hsa05168 | Herpes_simplex_virus_1_infection | 0.30 | 0.07 | 1.67E-05 | 3.92E-04 |
| 15 | hsa05320 | Autoimmune_thyroid_disease | 0.59 | 0.14 | 1.90E-05 | 4.17E-04 |
| 16 | hsa05140 | Leishmaniasis | 0.47 | 0.12 | 3.16E-05 | 6.50E-04 |
| 17 | hsa05169 | Epstein-Barr_virus_infection | 0.28 | 0.07 | 3.81E-05 | 7.37E-04 |
| 18 | hsa05416 | Viral_myocarditis | 0.51 | 0.13 | 4.15E-05 | 7.59E-04 |
| 19 | hsa05166 | Human_T-cell_leukemia_virus_1_infection | 0.23 | 0.06 | 6.56E-05 | 1.14E-03 |
| 20 | hsa04064 | NF-kappa_B_signaling_pathway | 0.37 | 0.10 | 9.21E-05 | 1.52E-03 |
| 21 | hsa04660 | T_cell_receptor_signaling_pathway | 0.36 | 0.10 | 1.12E-04 | 1.76E-03 |
| 22 | hsa04380 | Osteoclast_differentiation | 0.31 | 0.09 | 1.53E-04 | 2.29E-03 |
| 23 | hsa05164 | Influenza_A | 0.26 | 0.07 | 2.35E-04 | 3.37E-03 |
| 24 | hsa04662 | B_cell_receptor_signaling_pathway | 0.40 | 0.12 | 4.45E-04 | 6.11E-03 |
| 25 | hsa05144 | Malaria | 0.42 | 0.13 | 4.94E-04 | 6.50E-03 |
| 26 | hsa04650 | Natural_killer_cell_mediated_cytotoxicity | 0.30 | 0.09 | 6.91E-04 | 8.53E-03 |
| 27 | hsa05200 | Pathways_in_cancer | 0.13 | 0.04 | 7.00E-04 | 8.53E-03 |
| 28 | hsa05145 | Toxoplasmosis | 0.28 | 0.09 | 8.66E-04 | 1.02E-02 |
| 29 | hsa04612 | Antigen_processing_and_presentation | 0.39 | 0.13 | 1.25E-03 | 1.41E-02 |
| 30 | hsa04625 | C-type_lectin_receptor_signaling_pathway | 0.28 | 0.09 | 1.36E-03 | 1.48E-02 |
| 31 | hsa05133 | Pertussis | 0.36 | 0.12 | 1.40E-03 | 1.48E-02 |
| 32 | hsa04622 | RIG-I-like_receptor_signaling_pathway | 0.35 | 0.12 | 1.49E-03 | 1.53E-02 |
| 33 | hsa05134 | Legionellosis | 0.36 | 0.12 | 1.58E-03 | 1.58E-02 |
| 34 | hsa04668 | TNF_signaling_pathway | 0.27 | 0.09 | 1.89E-03 | 1.83E-02 |
| 35 | hsa05150 | Staphylococcus_aureus_infection | 0.40 | 0.14 | 1.98E-03 | 1.86E-02 |
| 36 | hsa04620 | Toll-like_receptor_signaling_pathway | 0.28 | 0.10 | 2.51E-03 | 2.30E-02 |
| 37 | hsa05152 | Tuberculosis | 0.20 | 0.07 | 3.73E-03 | 3.31E-02 |
| 38 | hsa05162 | Measles | 0.22 | 0.08 | 4.19E-03 | 3.63E-02 |
| 39 | hsa05143 | African_trypanosomiasis | 0.42 | 0.16 | 4.81E-03 | 4.06E-02 |
| 40 | hsa04217 | Necroptosis | 0.21 | 0.08 | 5.04E-03 | 4.14E-02 |
| 41 | hsa01523 | Antifolate_resistance | 0.40 | 0.16 | 5.96E-03 | 4.78E-02 |

**Supplemental Table S4. Significant PBC-associated genes identified by the S-MultiXcan tool**

| **Gene Name** | **CHR** | **Z mean** | **P** | **FDR** | **GWAS catalog** |
| --- | --- | --- | --- | --- | --- |
| *HLA-DRB1* | 6 | -2.97 | 4.95E-69 | 1.10E-64 | Yes |
| *HLA-DQA1* | 6 | -2.36 | 1.03E-43 | 1.15E-39 | No |
| *HLA-DRB5* | 6 | 0.60 | 1.17E-42 | 8.69E-39 | No |
| *BTNL2* | 6 | -2.66 | 3.26E-41 | 1.82E-37 | Yes |
| *HLA-DQA2* | 6 | 0.34 | 4.17E-39 | 1.86E-35 | No |
| *HLA-DPB1* | 6 | -2.01 | 5.91E-39 | 2.19E-35 | Yes |
| *HLA-DPA1* | 6 | -0.27 | 5.73E-35 | 1.82E-31 | No |
| *HLA-DOA* | 6 | 0.23 | 4.69E-34 | 1.31E-30 | No |
| *EGFL8* | 6 | -1.18 | 1.37E-31 | 3.39E-28 | No |
| *HSD17B8* | 6 | -5.49 | 2.66E-29 | 5.93E-26 | No |
| *IL12A* | 3 | -3.03 | 5.42E-29 | 1.10E-25 | Yes |
| *HLA-DQB1* | 6 | -5.09 | 7.04E-29 | 1.31E-25 | Yes |
| *TAP2* | 6 | -1.70 | 1.89E-26 | 3.24E-23 | No |
| *HLA-DRA* | 6 | 4.14 | 8.94E-26 | 1.42E-22 | Yes |
| *CYP21A2* | 6 | 3.22 | 5.24E-24 | 7.78E-21 | No |
| *COL11A2* | 6 | -5.50 | 1.05E-23 | 1.46E-20 | No |
| *IL12RB2* | 1 | 1.06 | 2.94E-21 | 3.85E-18 | Yes |
| *AIF1* | 6 | -1.82 | 2.47E-19 | 3.06E-16 | No |
| *CSNK2B* | 6 | 1.40 | 7.17E-19 | 8.41E-16 | No |
| *AGER* | 6 | 0.09 | 2.71E-18 | 3.02E-15 | No |
| *GPANK1* | 6 | 0.39 | 9.63E-18 | 1.02E-14 | No |
| *NFKBIL1* | 6 | -0.66 | 1.69E-17 | 1.71E-14 | No |
| *PRRC2A* | 6 | 0.85 | 1.80E-17 | 1.74E-14 | No |
| *C2* | 6 | 3.36 | 5.44E-17 | 5.05E-14 | No |
| *EXOC3L4* | 14 | -2.54 | 6.53E-17 | 5.82E-14 | Yes |
| *RNF5* | 6 | 1.96 | 1.11E-16 | 9.51E-14 | No |
| *HSPA1L* | 6 | 0.55 | 2.18E-16 | 1.80E-13 | No |
| *LY6G5B* | 6 | -7.92 | 2.43E-16 | 1.93E-13 | No |
| *PRRT1* | 6 | 4.11 | 9.01E-16 | 6.92E-13 | No |
| *BAG6* | 6 | -0.38 | 1.24E-15 | 9.21E-13 | No |
| *DDAH2* | 6 | -5.57 | 1.39E-15 | 9.99E-13 | No |
| *HSPA1B* | 6 | -3.54 | 2.04E-15 | 1.42E-12 | No |
| *LY6G5C* | 6 | -7.82 | 2.40E-15 | 1.62E-12 | No |
| *IRF5* | 7 | 7.11 | 2.62E-15 | 1.72E-12 | Yes |
| *PSMB9* | 6 | -3.25 | 3.31E-15 | 2.11E-12 | No |
| *AGPAT1* | 6 | 0.34 | 4.39E-15 | 2.72E-12 | No |
| *C6orf47* | 6 | -1.98 | 5.79E-15 | 3.49E-12 | No |
| *SFTA2* | 6 | -0.51 | 1.06E-14 | 6.21E-12 | No |
| *LY6G6F* | 6 | 0.27 | 2.35E-14 | 1.34E-11 | No |
| *MSH5* | 6 | -0.51 | 3.03E-14 | 1.69E-11 | No |
| *HLA-B* | 6 | 1.45 | 5.25E-14 | 2.85E-11 | No |
| *CLIC1* | 6 | 0.02 | 1.78E-13 | 9.22E-11 | No |
| *ABHD16A* | 6 | -0.09 | 2.08E-13 | 1.05E-10 | No |
| *IQCJ-SCHIP1* | 3 | 7.28 | 2.23E-13 | 1.10E-10 | No |
| *VWA7* | 6 | 0.51 | 4.51E-13 | 2.14E-10 | No |
| *SLC44A4* | 6 | 2.05 | 4.73E-13 | 2.20E-10 | No |
| *SCHIP1* | 3 | -2.51 | 5.22E-13 | 2.37E-10 | Yes |
| *SOCS1* | 16 | -3.26 | 9.33E-13 | 4.16E-10 | Yes |
| *LY6G6D* | 6 | 0.84 | 2.87E-12 | 1.25E-09 | No |
| *NAB1* | 2 | 1.30 | 4.47E-12 | 1.91E-09 | Yes |
| *PSORS1C2* | 6 | 0.06 | 6.12E-12 | 2.52E-09 | No |
| *SERBP1* | 1 | 3.49 | 3.75E-11 | 1.52E-08 | Yes |
| *PBX2* | 6 | -1.83 | 7.53E-11 | 3.00E-08 | No |
| *CLEC16A* | 16 | -1.04 | 1.04E-10 | 4.06E-08 | Yes |
| *DDX6* | 11 | 3.52 | 1.10E-10 | 4.22E-08 | Yes |
| *C6orf15* | 6 | 0.92 | 1.46E-10 | 5.51E-08 | No |
| *DENND1B* | 1 | 2.68 | 1.61E-10 | 5.98E-08 | Yes |
| *TAP1* | 6 | -3.25 | 1.97E-10 | 7.19E-08 | No |
| *EHMT2* | 6 | -0.51 | 3.17E-10 | 1.12E-07 | No |
| *PSMB8* | 6 | 1.83 | 3.22E-10 | 1.12E-07 | No |
| *STK19* | 6 | -3.11 | 8.05E-10 | 2.76E-07 | No |
| *TNPO3* | 7 | 0.79 | 9.92E-10 | 3.35E-07 | Yes |
| *IKZF3* | 17 | 3.93 | 1.15E-09 | 3.82E-07 | Yes |
| *SAPCD1* | 6 | 1.12 | 1.47E-09 | 4.75E-07 | No |
| *ZPBP2* | 17 | 3.02 | 1.47E-09 | 4.75E-07 | Yes |
| *NELFE* | 6 | 0.18 | 1.63E-09 | 5.19E-07 | No |
| *SYNGR1* | 22 | -1.30 | 2.83E-09 | 8.88E-07 | Yes |
| *NEU1* | 6 | 0.12 | 3.23E-09 | 9.99E-07 | No |
| *SLC39A7* | 6 | -3.80 | 3.71E-09 | 1.13E-06 | No |
| *TMEM39A* | 3 | -2.16 | 4.65E-09 | 1.40E-06 | Yes |
| *HLA-C* | 6 | -3.30 | 5.32E-09 | 1.58E-06 | No |
| *RGL2* | 6 | 0.62 | 6.01E-09 | 1.74E-06 | No |
| *MIEN1* | 17 | -1.55 | 8.40E-09 | 2.40E-06 | No |
| *C4B* | 6 | -0.10 | 9.14E-09 | 2.58E-06 | No |
| *C6orf48* | 6 | -4.46 | 9.57E-09 | 2.66E-06 | No |
| *NFKB1* | 4 | -0.03 | 1.01E-08 | 2.78E-06 | Yes |
| *HIST1H3H* | 6 | -1.22 | 1.40E-08 | 3.79E-06 | No |
| *PGBD1* | 6 | -3.11 | 1.43E-08 | 3.79E-06 | No |
| *NCR3* | 6 | 0.62 | 1.43E-08 | 3.79E-06 | No |
| *ZKSCAN3* | 6 | 0.76 | 1.67E-08 | 4.33E-06 | No |
| *ZSCAN12* | 6 | -1.00 | 1.86E-08 | 4.76E-06 | No |
| *IL7R* | 5 | 3.19 | 2.37E-08 | 6.00E-06 | Yes |
| *ZSCAN23* | 6 | -2.03 | 3.25E-08 | 8.13E-06 | No |
| *RPL3* | 22 | -0.87 | 3.64E-08 | 9.01E-06 | Yes |
| *LY6G6C* | 6 | 2.91 | 3.70E-08 | 9.06E-06 | No |
| *GSDMB* | 17 | -2.67 | 3.81E-08 | 9.12E-06 | Yes |
| *HIST1H2BK* | 6 | -0.04 | 3.85E-08 | 9.12E-06 | No |
| *HLA-DQB2* | 6 | 2.42 | 3.89E-08 | 9.12E-06 | No |
| *LTB* | 6 | -4.03 | 3.98E-08 | 9.24E-06 | No |
| *RING1* | 6 | 1.36 | 4.89E-08 | 1.12E-05 | No |
| *TNFAIP2* | 14 | 1.39 | 6.12E-08 | 1.39E-05 | Yes |
| *HSPA1A* | 6 | -4.03 | 6.57E-08 | 1.48E-05 | No |
| *MYBPC2* | 19 | 0.23 | 6.70E-08 | 1.49E-05 | Yes |
| *IFT80* | 3 | -4.98 | 6.78E-08 | 1.50E-05 | No |
| *TRIM59* | 3 | -3.11 | 8.05E-08 | 1.76E-05 | No |
| *ZSCAN31* | 6 | -0.90 | 8.44E-08 | 1.83E-05 | No |
| *HLA-DMA* | 6 | -3.99 | 8.87E-08 | 1.90E-05 | No |
| *TYK2* | 19 | -1.82 | 9.93E-08 | 2.10E-05 | Yes |
| *SMC4* | 3 | -1.34 | 1.00E-07 | 2.10E-05 | No |
| *CCHCR1* | 6 | 0.28 | 1.02E-07 | 2.12E-05 | No |
| *PNMT* | 17 | -1.25 | 1.33E-07 | 2.74E-05 | No |
| *CRB1* | 1 | 0.22 | 1.39E-07 | 2.84E-05 | No |
| *LRRC37A2* | 17 | -0.55 | 1.44E-07 | 2.92E-05 | No |
| *TCF19* | 6 | 4.55 | 1.49E-07 | 2.99E-05 | No |
| *HLA-G* | 6 | 3.10 | 1.54E-07 | 3.06E-05 | No |
| *C6orf10* | 6 | 5.19 | 2.10E-07 | 4.14E-05 | Yes |
| *CXCL11* | 4 | -0.47 | 2.14E-07 | 4.18E-05 | No |
| *HIST1H4I* | 6 | -0.79 | 2.34E-07 | 4.53E-05 | No |
| *TRIM27* | 6 | 0.86 | 2.42E-07 | 4.65E-05 | No |
| *HIST1H2AJ* | 6 | 5.16 | 2.53E-07 | 4.82E-05 | No |
| *BRD2* | 6 | -1.18 | 3.59E-07 | 6.72E-05 | No |
| *PPP1R14B* | 11 | 4.68 | 3.65E-07 | 6.78E-05 | No |
| *ZGLP1* | 19 | 2.95 | 3.74E-07 | 6.89E-05 | No |
| *LBH* | 2 | -0.15 | 4.35E-07 | 7.94E-05 | Yes |
| *TCAP* | 17 | -3.06 | 5.06E-07 | 9.16E-05 | No |
| *RMI2* | 16 | -3.53 | 5.74E-07 | 1.03E-04 | No |
| *WDR46* | 6 | 0.95 | 6.09E-07 | 1.09E-04 | No |
| *NKAPL* | 6 | 1.41 | 6.27E-07 | 1.11E-04 | No |
| *FKBP2* | 11 | 0.49 | 6.68E-07 | 1.16E-04 | No |
| *ORMDL3* | 17 | -2.92 | 7.07E-07 | 1.22E-04 | Yes |
| *CCDC88B* | 11 | 4.13 | 1.04E-06 | 1.78E-04 | Yes |
| *KPNA4* | 3 | -1.22 | 1.07E-06 | 1.82E-04 | No |
| *PRM3* | 16 | -4.87 | 1.10E-06 | 1.86E-04 | No |
| *BTN1A1* | 6 | -0.95 | 1.12E-06 | 1.88E-04 | No |
| *SKIV2L* | 6 | 1.00 | 1.14E-06 | 1.90E-04 | No |
| *PPP1R18* | 6 | 1.33 | 1.25E-06 | 2.05E-04 | No |
| *PRDX5* | 11 | -0.61 | 1.25E-06 | 2.05E-04 | No |
| *HLA-DMB* | 6 | -1.83 | 1.27E-06 | 2.07E-04 | No |
| *PDGFB* | 22 | -0.62 | 1.30E-06 | 2.10E-04 | Yes |
| *DEXI* | 16 | 4.03 | 1.36E-06 | 2.18E-04 | Yes |
| *MANBA* | 4 | -1.45 | 1.48E-06 | 2.35E-04 | Yes |
| *TNFRSF1A* | 12 | -2.35 | 1.55E-06 | 2.45E-04 | Yes |
| *RPS6KA4* | 11 | 1.19 | 1.56E-06 | 2.45E-04 | Yes |
| *MFSD6* | 2 | -0.32 | 1.61E-06 | 2.51E-04 | No |
| *CDK12* | 17 | 3.67 | 1.64E-06 | 2.54E-04 | No |
| *MED1* | 17 | -2.13 | 1.76E-06 | 2.70E-04 | No |
| *STARD3* | 17 | 0.69 | 1.81E-06 | 2.76E-04 | No |
| *HIST1H1B* | 6 | 2.53 | 1.94E-06 | 2.92E-04 | No |
| *TREH* | 11 | 1.72 | 1.94E-06 | 2.92E-04 | No |
| *CFB* | 6 | -1.41 | 2.12E-06 | 3.17E-04 | No |
| *IDUA* | 4 | 3.95 | 2.37E-06 | 3.47E-04 | Yes |
| *HIST1H2BN* | 6 | 2.38 | 2.78E-06 | 4.02E-04 | No |
| *DNAJC4* | 11 | 2.97 | 3.39E-06 | 4.87E-04 | No |
| *STAT1* | 2 | 0.63 | 3.49E-06 | 4.98E-04 | Yes |
| *LST1* | 6 | 0.05 | 3.53E-06 | 5.01E-04 | No |
| *NRM* | 6 | 0.73 | 3.67E-06 | 5.17E-04 | No |
| *HLA-A* | 6 | 0.11 | 3.73E-06 | 5.23E-04 | No |
| *DGKQ* | 4 | 3.24 | 3.86E-06 | 5.37E-04 | Yes |
| *ZKSCAN4* | 6 | -3.70 | 3.90E-06 | 5.40E-04 | No |
| *TIMMDC1* | 3 | -0.44 | 3.95E-06 | 5.43E-04 | Yes |
| *CDC37* | 19 | -4.22 | 4.12E-06 | 5.63E-04 | No |
| *RPP21* | 6 | 1.71 | 4.40E-06 | 5.98E-04 | No |
| *NINJ2* | 12 | -0.72 | 7.38E-06 | 9.84E-04 | No |
| *TNFSF11* | 13 | -3.04 | 7.74E-06 | 1.03E-03 | No |
| *YIPF2* | 19 | 4.23 | 8.05E-06 | 1.06E-03 | No |
| *FCRL3* | 1 | -4.40 | 8.14E-06 | 1.07E-03 | No |
| *ABT1* | 6 | -1.33 | 8.43E-06 | 1.10E-03 | No |
| *IER3* | 6 | 1.94 | 8.51E-06 | 1.10E-03 | No |
| *RNF39* | 6 | 0.64 | 8.58E-06 | 1.10E-03 | No |
| *ZCRB1* | 12 | 3.22 | 9.34E-06 | 1.20E-03 | No |
| *SLC26A1* | 4 | -4.24 | 1.06E-05 | 1.34E-03 | No |
| *HLA-E* | 6 | -0.02 | 1.07E-05 | 1.35E-03 | No |
| *BTN2A2* | 6 | -1.85 | 1.12E-05 | 1.40E-03 | No |
| *HIST1H2BD* | 6 | -3.17 | 1.17E-05 | 1.45E-03 | No |
| *AGAP5* | 10 | -4.28 | 1.29E-05 | 1.59E-03 | No |
| *PRM1* | 16 | 1.04 | 1.43E-05 | 1.75E-03 | No |
| *PRSS16* | 6 | -2.66 | 1.47E-05 | 1.78E-03 | No |
| *ARHGAP31* | 3 | 0.02 | 1.59E-05 | 1.90E-03 | No |
| *ARL5C* | 17 | -2.86 | 1.71E-05 | 2.04E-03 | No |
| *CARM1* | 19 | -2.90 | 2.11E-05 | 2.49E-03 | No |
| *ZSCAN16* | 6 | -0.67 | 2.24E-05 | 2.63E-03 | No |
| *TNF* | 6 | -1.51 | 2.37E-05 | 2.75E-03 | No |
| *GPR137* | 11 | 0.94 | 2.54E-05 | 2.93E-03 | No |
| *ATXN2* | 12 | -0.76 | 2.79E-05 | 3.20E-03 | Yes |
| *UBE2D3* | 4 | -4.18 | 2.83E-05 | 3.23E-03 | No |
| *FBXL20* | 17 | 3.60 | 3.22E-05 | 3.66E-03 | No |
| *SDAD1* | 4 | -0.73 | 3.25E-05 | 3.67E-03 | No |
| *CISD2* | 4 | -4.06 | 3.26E-05 | 3.67E-03 | Yes |
| *KCNC3* | 19 | 2.42 | 3.64E-05 | 4.05E-03 | No |
| *LTA* | 6 | 0.86 | 3.82E-05 | 4.23E-03 | No |
| *HIST1H2BJ* | 6 | 3.78 | 4.15E-05 | 4.58E-03 | No |
| *HIST1H2BF* | 6 | -0.40 | 4.50E-05 | 4.91E-03 | No |
| *TRIM31* | 6 | 1.97 | 4.56E-05 | 4.96E-03 | No |
| *GAK* | 4 | -0.92 | 4.62E-05 | 5.00E-03 | Yes |
| *LTBR* | 12 | 4.25 | 5.31E-05 | 5.66E-03 | No |
| *PHLDB1* | 11 | -1.88 | 5.41E-05 | 5.74E-03 | No |
| *HIST1H2BE* | 6 | -4.05 | 5.56E-05 | 5.87E-03 | No |
| *ZNF184* | 6 | 1.66 | 5.59E-05 | 5.87E-03 | No |
| *ICAM5* | 19 | 2.66 | 5.63E-05 | 5.89E-03 | No |
| *TAB1* | 22 | 0.78 | 5.71E-05 | 5.94E-03 | No |
| *GABBR1* | 6 | 0.95 | 5.84E-05 | 6.05E-03 | No |
| *PGAP3* | 17 | -3.71 | 6.04E-05 | 6.23E-03 | No |
| *ZSCAN9* | 6 | -1.66 | 6.26E-05 | 6.43E-03 | No |
| *NAAA* | 4 | 4.08 | 6.73E-05 | 6.88E-03 | No |
| *CAPSL* | 5 | -3.26 | 7.15E-05 | 7.27E-03 | Yes |
| *MICA* | 6 | -1.63 | 7.48E-05 | 7.57E-03 | No |
| *ICAM3* | 19 | -0.14 | 8.62E-05 | 8.69E-03 | No |
| *TRIM10* | 6 | 2.20 | 9.26E-05 | 9.29E-03 | No |
| *PPEF2* | 4 | 2.67 | 9.84E-05 | 9.79E-03 | No |
| *PSORS1C1* | 6 | 0.25 | 1.02E-04 | 1.01E-02 | No |
| *DCTPP1* | 16 | -1.07 | 1.04E-04 | 1.02E-02 | No |
| *GSDMA* | 17 | 3.16 | 1.18E-04 | 1.16E-02 | No |
| *SH2B3* | 12 | -1.69 | 1.20E-04 | 1.17E-02 | Yes |
| *TRIM26* | 6 | -0.34 | 1.24E-04 | 1.20E-02 | No |
| *VARS* | 6 | -0.88 | 1.27E-04 | 1.22E-02 | No |
| *ZSWIM8* | 10 | -0.73 | 1.29E-04 | 1.24E-02 | No |
| *SEC24C* | 10 | -2.50 | 1.30E-04 | 1.25E-02 | No |
| *C3orf80* | 3 | 2.63 | 1.34E-04 | 1.27E-02 | No |
| *SLC9B1* | 4 | 1.39 | 1.37E-04 | 1.30E-02 | No |
| *FUT11* | 10 | 3.87 | 1.38E-04 | 1.30E-02 | No |
| *TRIM15* | 6 | 0.42 | 1.47E-04 | 1.37E-02 | No |
| *SPIB* | 19 | 0.10 | 1.52E-04 | 1.41E-02 | Yes |
| *NDST2* | 10 | 2.66 | 1.64E-04 | 1.50E-02 | No |
| *MICB* | 6 | 2.13 | 1.73E-04 | 1.58E-02 | No |
| *RAVER1* | 19 | 0.45 | 1.75E-04 | 1.60E-02 | No |
| *C5orf30* | 5 | 2.96 | 1.85E-04 | 1.67E-02 | Yes |
| *CTSH* | 15 | 1.75 | 2.05E-04 | 1.84E-02 | No |
| *ZKSCAN8* | 6 | -0.12 | 2.16E-04 | 1.93E-02 | No |
| *BRAP* | 12 | -3.94 | 2.20E-04 | 1.95E-02 | No |
| *ABCF1* | 6 | 0.83 | 2.26E-04 | 1.99E-02 | No |
| *FAM213B* | 1 | -0.56 | 2.28E-04 | 2.00E-02 | No |
| *HKR1* | 19 | 0.76 | 2.42E-04 | 2.12E-02 | No |
| *ANKRD27* | 19 | 1.90 | 2.45E-04 | 2.14E-02 | No |
| *LDLR* | 19 | -3.58 | 2.48E-04 | 2.16E-02 | No |
| *DDX28* | 16 | 1.07 | 2.56E-04 | 2.22E-02 | No |
| *OR11A1* | 6 | -3.65 | 2.57E-04 | 2.22E-02 | No |
| *POMC* | 2 | 0.03 | 2.60E-04 | 2.24E-02 | No |
| *HLA-DOB* | 6 | -0.77 | 2.62E-04 | 2.25E-02 | No |
| *ITGB8* | 7 | -2.21 | 2.67E-04 | 2.28E-02 | No |
| *TTC34* | 1 | 3.56 | 2.82E-04 | 2.39E-02 | No |
| *PLCL2* | 3 | 0.68 | 3.10E-04 | 2.59E-02 | Yes |
| *METTL1* | 12 | 2.19 | 3.11E-04 | 2.59E-02 | No |
| *TSFM* | 12 | 3.45 | 3.14E-04 | 2.61E-02 | No |
| *CYP27B1* | 12 | 3.15 | 3.20E-04 | 2.65E-02 | No |
| *SIRPG* | 20 | -0.06 | 3.22E-04 | 2.66E-02 | No |
| *CXCL10* | 4 | -2.57 | 3.29E-04 | 2.71E-02 | No |
| *ITGAL* | 16 | -2.13 | 3.37E-04 | 2.76E-02 | No |
| *RBKS* | 2 | 3.54 | 3.62E-04 | 2.94E-02 | No |
| *ATAT1* | 6 | 1.03 | 3.79E-04 | 3.06E-02 | No |
| *IL12B* | 5 | -3.55 | 3.80E-04 | 3.06E-02 | Yes |
| *ALDH2* | 12 | 3.23 | 3.84E-04 | 3.08E-02 | Yes |
| *PLEKHM1* | 17 | -0.53 | 3.87E-04 | 3.09E-02 | No |
| *KCNJ15* | 21 | 0.35 | 3.98E-04 | 3.16E-02 | No |
| *METTL21B* | 12 | -3.52 | 3.99E-04 | 3.16E-02 | No |
| *HIST1H4E* | 6 | -1.17 | 4.00E-04 | 3.16E-02 | No |
| *ESRP2* | 16 | 2.98 | 4.21E-04 | 3.32E-02 | No |
| *MYLPF* | 16 | 3.52 | 4.35E-04 | 3.41E-02 | No |
| *GALC* | 14 | 0.99 | 4.45E-04 | 3.46E-02 | No |
| *NFATC3* | 16 | -0.12 | 4.48E-04 | 3.46E-02 | No |
| *BTN3A2* | 6 | 3.43 | 4.64E-04 | 3.56E-02 | No |
| *KIFC1* | 6 | 1.11 | 4.65E-04 | 3.56E-02 | No |
| *SLC7A6* | 16 | -0.90 | 4.66E-04 | 3.56E-02 | No |
| *PPP1R1B* | 17 | 0.22 | 4.80E-04 | 3.66E-02 | No |
| *HIBCH* | 2 | -1.44 | 4.98E-04 | 3.78E-02 | No |
| *BRE* | 2 | 0.33 | 5.13E-04 | 3.89E-02 | No |
| *FLOT1* | 6 | -2.29 | 5.33E-04 | 4.03E-02 | No |
| *CD58* | 1 | 0.52 | 5.46E-04 | 4.11E-02 | Yes |
| *RPS18* | 6 | 2.35 | 5.54E-04 | 4.15E-02 | No |
| *PRMT7* | 16 | 2.58 | 5.93E-04 | 4.42E-02 | No |
| *LRRC3C* | 17 | 3.43 | 6.03E-04 | 4.48E-02 | No |
| *CDKN2A* | 9 | -0.72 | 6.05E-04 | 4.48E-02 | No |
| *DPEP3* | 16 | -0.34 | 6.09E-04 | 4.49E-02 | No |
| *ERAP2* | 5 | 3.42 | 6.10E-04 | 4.49E-02 | No |
| *PDCD2L* | 19 | 2.25 | 6.19E-04 | 4.53E-02 | No |
| *ZBTB22* | 6 | 2.85 | 6.50E-04 | 4.75E-02 | No |
| *ACADM* | 1 | -0.91 | 6.83E-04 | 4.97E-02 | No |
| *BTN3A1* | 6 | -0.78 | 6.87E-04 | 4.98E-02 | No |
| *BAK1* | 6 | -2.46 | 6.89E-04 | 4.98E-02 | No |

**Supplemental Table S5. Significant genes identified from S-PrediXcan analysis by integrating GWAS summary data with GTEx liver eQTL data**

| **Gene name** | **Z score** | **P value** | **FDR** | **GWAS Catalog documented genes** |
| --- | --- | --- | --- | --- |
| *IRF5* | 8.43 | 3.57E-17 | 4.29E-13 | Yes |
| *HSD17B8* | -8.33 | 8.24E-17 | 4.95E-13 | No |
| *LY6G5C* | -8.21 | 2.16E-16 | 8.64E-13 | No |
| *CYP21A2* | 8.01 | 1.13E-15 | 3.39E-12 | No |
| *CSNK2B* | 7.70 | 1.34E-14 | 3.22E-11 | No |
| *LY6G5B* | -7.61 | 2.76E-14 | 5.52E-11 | No |
| *PRRT1* | 7.29 | 3.13E-13 | 5.25E-10 | No |
| *SOCS1* | -7.27 | 3.50E-13 | 5.25E-10 | Yes |
| *DDAH2* | -7.04 | 1.99E-12 | 2.65E-09 | No |
| *MANBA* | -5.99 | 2.05E-09 | 2.46E-06 | Yes |
| *IFT80* | -5.91 | 3.52E-09 | 3.84E-06 | No |
| *RP11-783K16.13* | 5.36 | 8.54E-08 | 8.54E-05 | No |
| *SMC4* | -5.31 | 1.10E-07 | 9.38E-05 | No |
| *HLA-DQB1* | -5.31 | 1.13E-07 | 9.38E-05 | Yes |
| *RMI2* | -5.29 | 1.20E-07 | 9.38E-05 | No |
| *DEXI* | 5.29 | 1.25E-07 | 9.38E-05 | Yes |
| *TCF19* | 5.27 | 1.34E-07 | 9.46E-05 | No |
| *HCG27* | 5.23 | 1.72E-07 | 1.15E-04 | No |
| *DGKQ* | 4.94 | 7.75E-07 | 4.90E-04 | Yes |
| *PSMB8-AS1* | -4.88 | 1.06E-06 | 6.36E-04 | No |
| *C6orf48* | -4.83 | 1.37E-06 | 7.83E-04 | No |
| *SYNGR1* | -4.82 | 1.46E-06 | 7.97E-04 | Yes |
| *YIPF2* | 4.73 | 2.21E-06 | 1.14E-03 | No |
| *CARM1* | -4.72 | 2.37E-06 | 1.14E-03 | No |
| *TNFRSF1A* | -4.72 | 2.37E-06 | 1.14E-03 | Yes |
| *NELFE* | -4.70 | 2.64E-06 | 1.22E-03 | No |
| *HLA-DMA* | -4.68 | 2.86E-06 | 1.27E-03 | No |
| *HLA-C* | -4.67 | 3.03E-06 | 1.30E-03 | No |
| *TREH* | 4.59 | 4.38E-06 | 1.81E-03 | No |
| *ICAM5* | 4.57 | 4.77E-06 | 1.91E-03 | No |
| *ORMDL3* | 4.52 | 6.07E-06 | 2.35E-03 | Yes |
| *HIST1H2BJ* | 4.50 | 6.70E-06 | 2.51E-03 | No |
| *UBE2D3* | -4.48 | 7.62E-06 | 2.76E-03 | No |
| *GRB7* | 4.47 | 7.83E-06 | 2.76E-03 | No |
| *FCRL3* | -4.45 | 8.39E-06 | 2.84E-03 | No |
| *NKAPL* | 4.45 | 8.51E-06 | 2.84E-03 | No |
| *RP1-86C11.7* | 4.36 | 1.28E-05 | 4.15E-03 | No |
| *HLA-DQA2* | 4.35 | 1.33E-05 | 4.20E-03 | No |
| *LTBR* | 4.33 | 1.46E-05 | 4.49E-03 | No |
| *AGAP5* | -4.33 | 1.50E-05 | 4.50E-03 | No |
| *ZCRB1* | -4.31 | 1.65E-05 | 4.83E-03 | No |
| *LTA* | 4.26 | 2.00E-05 | 5.72E-03 | No |
| *TTC34* | 4.25 | 2.10E-05 | 5.86E-03 | No |
| *NAB1* | -4.18 | 2.94E-05 | 8.02E-03 | Yes |
| *HCG20* | 4.16 | 3.17E-05 | 8.46E-03 | No |
| *RBKS* | 4.16 | 3.24E-05 | 8.46E-03 | No |
| *CTSH* | 4.15 | 3.34E-05 | 8.53E-03 | No |
| *MMEL1* | -4.12 | 3.80E-05 | 9.50E-03 | No |
| *FBXL20* | 4.10 | 4.11E-05 | 1.01E-02 | No |
| *IFITM3* | -4.07 | 4.66E-05 | 1.12E-02 | No |
| *MYBPC2* | -4.07 | 4.74E-05 | 1.12E-02 | Yes |
| *CD58* | 4.04 | 5.30E-05 | 1.22E-02 | Yes |
| *NAAA* | 4.03 | 5.54E-05 | 1.25E-02 | No |
| *LINC00240* | -3.97 | 7.28E-05 | 1.62E-02 | No |
| *MED1* | -3.96 | 7.55E-05 | 1.65E-02 | No |
| *CD2BP2* | 3.95 | 7.88E-05 | 1.69E-02 | No |
| *DDX28* | 3.92 | 8.69E-05 | 1.83E-02 | No |
| *KPNA4* | -3.89 | 1.01E-04 | 2.10E-02 | No |
| *CISD2* | -3.88 | 1.07E-04 | 2.17E-02 | Yes |
| *STAT6* | -3.87 | 1.09E-04 | 2.19E-02 | No |
| *SH2B3* | 3.85 | 1.20E-04 | 2.35E-02 | Yes |
| *TESK1* | -3.84 | 1.23E-04 | 2.38E-02 | No |
| *C9orf64* | -3.83 | 1.27E-04 | 2.41E-02 | No |
| *MFSD6* | -3.82 | 1.36E-04 | 2.55E-02 | No |
| *TNFRSF14* | 3.77 | 1.64E-04 | 3.04E-02 | No |
| *SEPT1* | -3.72 | 1.99E-04 | 3.62E-02 | No |
| *IDUA* | 3.69 | 2.25E-04 | 3.94E-02 | Yes |
| *UVSSA* | 3.69 | 2.25E-04 | 3.94E-02 | No |
| *STARD3* | 3.69 | 2.26E-04 | 3.94E-02 | No |
| *ESRP2* | 3.68 | 2.30E-04 | 3.94E-02 | No |
| *ZC3HAV1L* | -3.65 | 2.62E-04 | 4.42E-02 | No |
| *ZSCAN26* | 3.65 | 2.65E-04 | 4.42E-02 | No |
| *GSDMA* | 3.62 | 3.00E-04 | 4.87E-02 | No |
| *HCP5B* | -3.61 | 3.01E-04 | 4.87E-02 | No |
| *TSFM* | 3.61 | 3.04E-04 | 4.87E-02 | No |
| *METTL1* | 3.60 | 3.13E-04 | 4.95E-02 | No |

**Supplemental Table S6. Significant genes identified from S-PrediXcan analysis by integrating GWAS summary data with GTEx blood eQTL data**

| **Gene name** | **Z score** | **P value** | **FDR** | **GWAS Catalog documented genes** |
| --- | --- | --- | --- | --- |
| *IL12RB2* | 10.81 | 3.06E-27 | 3.68E-23 | Yes |
| *IRF5* | 8.23 | 1.87E-16 | 1.12E-12 | Yes |
| *HSPA1B* | -7.99 | 1.36E-15 | 5.45E-12 | No |
| *LY6G5C* | -7.89 | 3.01E-15 | 9.05E-12 | No |
| *DDAH2* | -7.79 | 6.47E-15 | 1.56E-11 | No |
| *LY6G5B* | -7.48 | 7.66E-14 | 1.54E-10 | No |
| *SOCS1* | -7.25 | 4.27E-13 | 7.34E-10 | Yes |
| *EHMT2* | -7.14 | 9.01E-13 | 1.35E-09 | No |
| *LY6G6D* | -7.11 | 1.17E-12 | 1.56E-09 | No |
| *DDX6* | 6.95 | 3.67E-12 | 4.41E-09 | Yes |
| *IKZF3* | 6.65 | 2.92E-11 | 3.19E-08 | Yes |
| *SYNGR1* | 6.43 | 1.30E-10 | 1.30E-07 | Yes |
| *HLA-G* | 6.22 | 4.85E-10 | 4.49E-07 | No |
| *SLC44A4* | -6.16 | 7.41E-10 | 6.37E-07 | No |
| *TYK2* | -6.01 | 1.91E-09 | 1.53E-06 | Yes |
| *CSNK2B* | 5.93 | 3.04E-09 | 2.24E-06 | No |
| *TCF19* | 5.92 | 3.16E-09 | 2.24E-06 | No |
| *GSDMB* | -5.90 | 3.53E-09 | 2.25E-06 | Yes |
| *SMC4* | -5.90 | 3.55E-09 | 2.25E-06 | No |
| *NFKB1* | 5.61 | 2.08E-08 | 1.25E-05 | Yes |
| *TNPO3* | -5.41 | 6.30E-08 | 3.61E-05 | Yes |
| *ORMDL3* | -5.27 | 1.35E-07 | 7.38E-05 | Yes |
| *HLA-DQA2* | -5.25 | 1.49E-07 | 7.79E-05 | No |
| *RP11-396B14.2* | -5.21 | 1.93E-07 | 9.67E-05 | No |
| *LY6G6C* | -5.13 | 2.82E-07 | 1.36E-04 | No |
| *ZSCAN31* | 5.09 | 3.65E-07 | 1.66E-04 | No |
| *BTN2A2* | -5.08 | 3.73E-07 | 1.66E-04 | No |
| *ZKSCAN3* | -4.97 | 6.67E-07 | 2.86E-04 | No |
| *AIF1* | -4.96 | 7.04E-07 | 2.86E-04 | No |
| *ZKSCAN4* | -4.96 | 7.14E-07 | 2.86E-04 | No |
| *HLA-DRB5* | 4.93 | 8.38E-07 | 3.25E-04 | No |
| *DGKQ* | 4.91 | 8.94E-07 | 3.36E-04 | Yes |
| *HCP5B* | -4.78 | 1.73E-06 | 6.19E-04 | No |
| *C6orf48* | -4.78 | 1.75E-06 | 6.19E-04 | No |
| *ZSCAN12* | 4.73 | 2.29E-06 | 7.75E-04 | No |
| *CCDC88B* | 4.72 | 2.32E-06 | 7.75E-04 | Yes |
| *PGBD1* | -4.71 | 2.50E-06 | 8.13E-04 | No |
| *CARM1* | -4.68 | 2.82E-06 | 8.92E-04 | No |
| *HLA-DMA* | -4.67 | 2.97E-06 | 9.16E-04 | No |
| *VWA7* | -4.62 | 3.88E-06 | 1.17E-03 | No |
| *TAB1* | 4.54 | 5.63E-06 | 1.65E-03 | No |
| *MIEN1* | 4.48 | 7.47E-06 | 2.14E-03 | No |
| *HIST1H2BE* | -4.41 | 1.03E-05 | 2.84E-03 | No |
| *FCRL3* | -4.41 | 1.04E-05 | 2.84E-03 | No |
| *RP1-86C11.7* | 4.36 | 1.28E-05 | 3.40E-03 | No |
| *AGAP5* | -4.36 | 1.30E-05 | 3.40E-03 | No |
| *SAPCD1* | -4.35 | 1.34E-05 | 3.41E-03 | No |
| *LTB* | -4.35 | 1.36E-05 | 3.41E-03 | No |
| *KPNA4* | -4.33 | 1.46E-05 | 3.58E-03 | No |
| *PGAP3* | -4.32 | 1.58E-05 | 3.80E-03 | No |
| *PLCG2* | -4.30 | 1.68E-05 | 3.96E-03 | No |
| *HIST1H2BK* | 4.28 | 1.90E-05 | 4.39E-03 | No |
| *CTA-14H9.5* | 4.27 | 1.98E-05 | 4.49E-03 | No |
| *GPANK1* | -4.26 | 2.06E-05 | 4.59E-03 | No |
| *PPP1R1B* | -4.23 | 2.33E-05 | 5.02E-03 | No |
| *TTC34* | 4.23 | 2.38E-05 | 5.02E-03 | No |
| *ZCRB1* | 4.22 | 2.41E-05 | 5.02E-03 | No |
| *HKR1* | 4.22 | 2.42E-05 | 5.02E-03 | No |
| *UBE2D3* | -4.22 | 2.49E-05 | 5.08E-03 | No |
| *SLC26A1* | -4.20 | 2.66E-05 | 5.33E-03 | No |
| *RP11-574K11.29* | 4.14 | 3.44E-05 | 6.78E-03 | No |
| *NAAA* | 4.14 | 3.53E-05 | 6.85E-03 | No |
| *NOTCH4* | 4.12 | 3.83E-05 | 7.31E-03 | No |
| *PPHLN1* | 4.10 | 4.07E-05 | 7.65E-03 | No |
| *RPL3* | -4.07 | 4.70E-05 | 8.70E-03 | Yes |
| *PLCL2* | 4.00 | 6.43E-05 | 1.17E-02 | Yes |
| *SLC7A6* | -3.98 | 6.95E-05 | 1.25E-02 | No |
| *ZKSCAN8* | -3.94 | 8.22E-05 | 1.45E-02 | No |
| *MED1* | -3.92 | 8.91E-05 | 1.53E-02 | No |
| *FUT11* | 3.92 | 8.91E-05 | 1.53E-02 | No |
| *GTF2H1* | -3.91 | 9.30E-05 | 1.58E-02 | No |
| *HLA-DQA1* | -3.88 | 1.03E-04 | 1.70E-02 | No |
| *ANKRD27* | 3.88 | 1.05E-04 | 1.70E-02 | No |
| *KPNB1* | -3.88 | 1.06E-04 | 1.70E-02 | No |
| *FAM166B* | 3.88 | 1.06E-04 | 1.70E-02 | No |
| *PNMT* | -3.87 | 1.11E-04 | 1.75E-02 | No |
| *SH2B3* | 3.85 | 1.20E-04 | 1.87E-02 | Yes |
| *LCAT* | 3.84 | 1.25E-04 | 1.92E-02 | No |
| *MANBA* | -3.78 | 1.57E-04 | 2.39E-02 | Yes |
| *ALDH2* | -3.77 | 1.61E-04 | 2.41E-02 | Yes |
| *OR2H2* | -3.77 | 1.63E-04 | 2.41E-02 | No |
| *CLEC7A* | -3.76 | 1.68E-04 | 2.47E-02 | No |
| *HLA-DQB2* | 3.75 | 1.76E-04 | 2.54E-02 | No |
| *WNT3* | -3.75 | 1.77E-04 | 2.54E-02 | No |
| *TRIM59* | -3.75 | 1.79E-04 | 2.54E-02 | No |
| *RP11-81H14.2* | 3.74 | 1.86E-04 | 2.55E-02 | No |
| *HLA-DMB* | 3.74 | 1.87E-04 | 2.55E-02 | No |
| *ZBTB12* | -3.74 | 1.87E-04 | 2.55E-02 | No |
| *HIST1H4I* | 3.73 | 1.91E-04 | 2.57E-02 | No |
| *DAXX* | -3.72 | 2.01E-04 | 2.69E-02 | No |
| *PLA2G15* | -3.71 | 2.11E-04 | 2.78E-02 | No |
| *GPX3* | 3.69 | 2.26E-04 | 2.96E-02 | No |
| *FAM213B* | -3.68 | 2.34E-04 | 3.02E-02 | No |
| *SLC6A19* | -3.67 | 2.44E-04 | 3.12E-02 | No |
| *MSH5* | -3.67 | 2.47E-04 | 3.13E-02 | No |
| *CTSH* | 3.64 | 2.76E-04 | 3.46E-02 | No |
| *DPEP2* | 3.63 | 2.79E-04 | 3.46E-02 | No |
| *METTL1* | -3.60 | 3.13E-04 | 3.81E-02 | No |
| *METTL21B* | -3.60 | 3.13E-04 | 3.81E-02 | No |
| *IDUA* | 3.60 | 3.22E-04 | 3.87E-02 | Yes |
| *OGFOD2* | 3.59 | 3.26E-04 | 3.87E-02 | No |
| *TSFM* | 3.59 | 3.28E-04 | 3.87E-02 | No |
| *MICB* | 3.58 | 3.39E-04 | 3.95E-02 | No |
| *PRPF40A* | -3.58 | 3.48E-04 | 4.02E-02 | No |
| *ERAP2* | 3.57 | 3.55E-04 | 4.07E-02 | No |
| *CD226* | -3.56 | 3.74E-04 | 4.24E-02 | No |
| *TMEM163* | 3.55 | 3.86E-04 | 4.33E-02 | No |
| *GOPC* | 3.54 | 4.01E-04 | 4.46E-02 | No |
| *ARL6IP6* | 3.54 | 4.05E-04 | 4.46E-02 | No |
| *BTN3A2* | 3.53 | 4.23E-04 | 4.57E-02 | No |
| *ZBTB22* | 3.52 | 4.25E-04 | 4.57E-02 | No |
| *MFSD6* | -3.52 | 4.29E-04 | 4.57E-02 | No |
| *TRIM10* | 3.52 | 4.29E-04 | 4.57E-02 | No |
| *PRMT7* | 3.52 | 4.35E-04 | 4.59E-02 | No |
| *UQCRC1* | 3.50 | 4.69E-04 | 4.90E-02 | No |

**Supplemental Table S7. Phenotype-based enrichment analysis of these 29 identified risk genes for PBC based on the GLAD4U database**

| **Gene Set** | **Description** | **Size** | **Expect** | **Ratio** | **P Value** |
| --- | --- | --- | --- | --- | --- |
| PA443464 | Autoimmune Diseases | 497 | 0.87 | 10.37 | 8.47E-08 |
| PA443888 | Diabetes Mellitus, Type 1 | 171 | 0.30 | 13.40 | 2.04E-04 |
| PA443780 | Connective Tissue Diseases | 372 | 0.65 | 7.70 | 3.99E-04 |
| PA444822 | Lupus Erythematosus, Systemic | 233 | 0.41 | 9.83 | 6.61E-04 |
| PA446552 | Machado-Joseph Disease | 27 | 0.05 | 42.42 | 1.00E-03 |
| PA444602 | Immune System Diseases | 739 | 1.29 | 4.65 | 1.41E-03 |
| PA165108310 | Dermatitis medicamentosa | 33 | 0.06 | 34.71 | 1.50E-03 |
| PA444797 | Liver Cirrhosis | 148 | 0.26 | 11.61 | 2.11E-03 |
| PA443433 | Arthritis, Juvenile Rheumatoid | 42 | 0.07 | 27.27 | 2.41E-03 |
| PA443652 | Celiac Disease | 163 | 0.28 | 10.54 | 2.77E-03 |

**Supplemental Table S8. PBC-associated risk genes matched in druggable gene categories**

| **Druggable Gene Category** | **Matching Gene Count** | **Matching Genes** | **Non-Matching Genes** |
| --- | --- | --- | --- |
| ENZYME | 9 | *CSNK2B, DDAH2, MANBA, IDUA, CARM1, UBE2D3, NAAA, CTSH, TSFM* | *LY6G5B, LY6G5C, IRF5, SOCS1, HLA-DMA, TCF19, KPNA4, MFSD6, MED1, DGKQ, FCRL3, SH2B3* |
| DRUGGABLE GENOME | 6 | *MANBA, MFSD6, FCRL3, CARM1, NAAA, CTSH* | *CSNK2B, LY6G5B, DDAH2, LY6G5C, IRF5, SOCS1, HLA-DMA, TCF19, KPNA4, MED1, IDUA, DGKQ, UBE2D3, SH2B3, TSFM* |
| KINASE | 4 | *CSNK2B, SOCS1, DGKQ, SH2B3* | *LY6G5B, DDAH2, LY6G5C, IRF5, HLA-DMA, TCF19, KPNA4, MANBA, MFSD6, MED1, IDUA, FCRL3, CARM1, UBE2D3, NAAA, CTSH, TSFM* |
| CLINICALLY ACTIONABLE | 3 | *SOCS1, HLA-DMA, SH2B3* | *CSNK2B, LY6G5B, DDAH2, LY6G5C, IRF5, TCF19, KPNA4, MANBA, MFSD6, MED1, IDUA, DGKQ, FCRL3, CARM1, UBE2D3, NAAA, CTSH, TSFM* |
| TRANSCRIPTION FACTOR | 3 | *IRF5, TCF19, MED1* | *CSNK2B, LY6G5B, DDAH2, LY6G5C, SOCS1, HLA-DMA, KPNA4, MANBA, MFSD6, IDUA, DGKQ, FCRL3, CARM1, UBE2D3, NAAA, SH2B3, CTSH, TSFM* |
| TRANSCRIPTION FACTOR BINDING | 3 | *CSNK2B, MED1, NAAA* | *LY6G5B, DDAH2, LY6G5C, IRF5, SOCS1, HLA-DMA, TCF19, KPNA4, MANBA, MFSD6, IDUA, DGKQ, FCRL3, CARM1, UBE2D3, SH2B3, CTSH, TSFM* |
| CELL SURFACE | 2 | *HLA-DMA, FCRL3* | *CSNK2B, LY6G5B, DDAH2, LY6G5C, IRF5, SOCS1, TCF19, KPNA4, MANBA, MFSD6, MED1, IDUA, DGKQ, CARM1, UBE2D3, NAAA, SH2B3, CTSH, TSFM* |
| EXTERNAL SIDE OF PLASMA MEMBRANE | 2 | *LY6G5B, LY6G5C* | *CSNK2B, DDAH2, IRF5, SOCS1, HLA-DMA, TCF19, KPNA4, MANBA, MFSD6, MED1, IDUA, DGKQ, FCRL3, CARM1, UBE2D3, NAAA, SH2B3, CTSH, TSFM* |
| DNA REPAIR | 1 | *UBE2D3* | *CSNK2B, LY6G5B, DDAH2, LY6G5C, IRF5, SOCS1, HLA-DMA, TCF19, KPNA4, MANBA, MFSD6, MED1, IDUA, DGKQ, FCRL3, CARM1, NAAA, SH2B3, CTSH, TSFM* |
| METHYL TRANSFERASE | 1 | *CARM1* | *CSNK2B, LY6G5B, DDAH2, LY6G5C, IRF5, SOCS1, HLA-DMA, TCF19, KPNA4, MANBA, MFSD6, MED1, IDUA, DGKQ, FCRL3, UBE2D3, NAAA, SH2B3, CTSH, TSFM* |

**Supplemental Table S9. MAGMA gene-property analysis identifies liver single cells based on the Mouse Cell Atlas**

| **VARIABLE** | **BETA** | **BETA STD** | **SE** | **P value** |
| --- | --- | --- | --- | --- |
| Dendritic_cell_Cst3_high | 0.39 | 0.04 | 0.15 | 5.80E-03 |
| T_cell_Trbc2_high | 0.40 | 0.04 | 0.16 | 7.10E-03 |
| Macrophage_Chil3_high | 0.35 | 0.03 | 0.16 | 1.67E-02 |
| T_cell_Gzma_high | 0.25 | 0.02 | 0.13 | 3.04E-02 |
| Dendritic_cell_Siglech_high | 0.18 | 0.02 | 0.14 | 8.96E-02 |

**Supplemental Table S10. Results of PBC-associated liver cell types by using Rolypoly-based integrative analysis of combining GWAS summary data with scRNA-seq data.**

| **Cell types** | **Bootstrap estimate** | **Bootstrap error** | **T value** | **P value** |
| --- | --- | --- | --- | --- |
| Cholangiocytes | 0.0119 | 0.0073 | 1.65 | 0.05 |
| Stellate cells | 0.0030 | 0.0031 | 0.95 | 0.17 |
| Sinusoidal endothelial cells | 0.0027 | 0.0053 | 0.52 | 0.302 |
| Inflammatory monocyte/macrophages | 0.0027 | 0.0065 | 0.41 | 0.339 |
| NK-like cells | 0.0018 | 0.0065 | 0.28 | 0.391 |
| Mature B cells | 0.0011 | 0.0044 | 0.24 | 0.404 |
| γδT cells | 0.0005 | 0.0042 | 0.11 | 0.456 |
| Non-inflammatory macrophages | -0.0005 | 0.0048 | -0.10 | 0.54 |
| RBCs | -0.0020 | 0.0050 | -0.39 | 0.653 |
| Plasma cells | -0.0028 | 0.0039 | -0.71 | 0.761 |
| Portal endothelial cells | -0.0035 | 0.0047 | -0.76 | 0.775 |
| Hepatocytes | -0.0055 | 0.0060 | -0.92 | 0.821 |
| T cells | -0.0084 | 0.0077 | -1.08 | 0.86 |

**Supplemental Table S11. The percentage of expressed 27 genetic risk genes in all 13 distinct cell types in human liver tissues**

| **Gene name** | **Portal endothelial cells** | **Cholangiocytes** | **Non-inflammatory macrophages** | **T cells** | **Inflammatory monocyte/macrophages** | **NK-like cells** | **RBCs** | **γδT cells** | **sinusoidal endothelial cells** | **Mature B cells** | **Stellate cells** | **Plasma cells** | **hepatocytes** |
| --- | --- | --- | --- | --- | --- | --- | --- | --- | --- | --- | --- | --- | --- |
| *CSNK2B* | 28.49% | 28.34% | 40.00% | 22.85% | 32.85% | 26.88% | 23.08% | 65.63% | 36.12% | 18.75% | 29.73% | 35.19% | 12.14% |
| *LY6G5B* | 0.19% | 0.11% | 0.00% | 0.29% | 0.09% | 0.20% | 0.00% | 0.00% | 0.00% | 0.00% | 0.00% | 0.00% | 0.00% |
| *DDAH2* | 14.34% | 8.67% | 36.99% | 6.12% | 16.21% | 5.70% | 3.53% | 19.79% | 30.10% | 5.47% | 37.84% | 3.85% | 4.85% |
| *LY6G5C* | 2.68% | 0.32% | 0.27% | 2.68% | 1.54% | 2.24% | 0.32% | 1.04% | 2.34% | 0.00% | 0.00% | 0.00% | 0.81% |
| *IRF5* | 0.00% | 0.39% | 6.30% | 0.86% | 6.40% | 1.22% | 0.32% | 1.04% | 0.00% | 1.56% | 0.00% | 1.15% | 0.16% |
| *SOCS1* | 1.91% | 1.13% | 0.82% | 14.72% | 5.46% | 26.48% | 0.32% | 8.33% | 1.00% | 5.47% | 5.41% | 1.15% | 0.32% |
| *SYNGR1* | 2.29% | 2.57% | 0.55% | 1.53% | 6.31% | 5.50% | 3.53% | 4.17% | 2.34% | 0.78% | 8.11% | 2.88% | 0.00% |
| *C6orf48* | 28.49% | 13.50% | 40.27% | 42.54% | 39.85% | 47.86% | 4.49% | 70.83% | 50.17% | 45.31% | 51.35% | 15.96% | 5.02% |
| *HLA-DMA* | 8.60% | 1.09% | 48.49% | 9.94% | 34.90% | 6.52% | 2.88% | 13.54% | 9.03% | 50.78% | 8.11% | 6.92% | 1.46% |
| *SMC4* | 9.37% | 4.97% | 5.75% | 7.17% | 5.29% | 8.55% | 8.33% | 71.88% | 7.02% | 3.91% | 2.70% | 31.15% | 4.85% |
| *TCF19* | 0.19% | 0.07% | 0.27% | 0.96% | 0.43% | 0.81% | 2.24% | 19.79% | 1.00% | 0.00% | 0.00% | 5.38% | 0.32% |
| *ORMDL3* | 1.34% | 22.38% | 0.82% | 10.42% | 4.61% | 9.37% | 18.91% | 8.33% | 2.34% | 0.78% | 0.00% | 7.50% | 11.65% |
| *KPNA4* | 9.56% | 9.73% | 12.60% | 8.99% | 10.84% | 8.35% | 6.41% | 13.54% | 11.37% | 2.34% | 2.70% | 5.38% | 3.40% |
| *MANBA* | 2.49% | 4.76% | 15.34% | 2.10% | 9.22% | 3.05% | 0.96% | 5.21% | 3.34% | 3.91% | 2.70% | 1.54% | 3.24% |
| *MFSD6* | 2.29% | 0.21% | 0.00% | 2.87% | 1.54% | 3.05% | 0.64% | 3.13% | 5.69% | 0.00% | 0.00% | 0.96% | 0.32% |
| *MED1* | 6.12% | 5.25% | 5.21% | 4.97% | 4.78% | 4.28% | 2.88% | 9.38% | 4.68% | 2.34% | 2.70% | 3.85% | 2.91% |
| *IDUA* | 0.76% | 0.63% | 0.55% | 0.57% | 0.94% | 1.22% | 0.64% | 0.00% | 1.34% | 0.00% | 2.70% | 0.58% | 0.49% |
| *DGKQ* | 0.96% | 1.41% | 0.82% | 1.63% | 1.96% | 2.24% | 0.96% | 2.08% | 1.00% | 0.78% | 0.00% | 0.19% | 1.13% |
| *FCRL3* | 0.00% | 0.04% | 0.27% | 2.77% | 1.96% | 3.87% | 0.32% | 3.13% | 0.00% | 5.47% | 2.70% | 0.58% | 0.16% |
| *ZCRB1* | 16.83% | 25.20% | 20.00% | 13.00% | 15.19% | 18.74% | 11.86% | 38.54% | 17.39% | 14.84% | 16.22% | 20.77% | 13.43% |
| *AGAP5* | 1.15% | 0.92% | 0.00% | 0.38% | 0.51% | 0.41% | 0.32% | 1.04% | 0.67% | 0.00% | 0.00% | 0.58% | 0.65% |
| *CARM1* | 4.02% | 6.91% | 4.66% | 1.05% | 3.75% | 1.63% | 6.09% | 0.00% | 5.35% | 0.78% | 2.70% | 6.35% | 4.85% |
| *UBE2D3* | 43.02% | 44.34% | 62.19% | 48.95% | 50.68% | 53.36% | 46.79% | 79.17% | 47.16% | 29.69% | 35.14% | 47.31% | 24.43% |
| *NAAA* | 5.35% | 9.87% | 24.38% | 7.27% | 16.55% | 2.44% | 4.17% | 3.13% | 3.68% | 7.03% | 8.11% | 3.65% | 3.88% |
| *SH2B3* | 12.81% | 3.00% | 26.58% | 2.87% | 17.06% | 3.67% | 1.60% | 2.08% | 7.36% | 2.34% | 2.70% | 0.96% | 1.62% |
| *CTSH* | 5.16% | 30.60% | 22.19% | 6.02% | 25.68% | 0.61% | 7.05% | 9.38% | 2.34% | 8.59% | 2.70% | 29.62% | 7.93% |
| *TTC34* | 0.19% | 0.00% | 0.27% | 0.00% | 0.26% | 0.20% | 0.00% | 0.00% | 0.00% | 0.00% | 0.00% | 0.00% | 0.00% |

**Supplemental Table S12. The significantly up-regulated DEGs among *ORMDL3^+^* cholangiocytes**

| **Gene** | **T score** | **Fold change** | **P value** | **FDR** |
| --- | --- | --- | --- | --- |
| *GALNT1* | 3.85 | 1.51 | 1.27E-04 | 1.86E-02 |
| *MPPED1* | 3.60 | 2.39 | 3.34E-04 | 3.53E-02 |
| *SLC25A30* | 3.58 | 1.60 | 3.63E-04 | 3.60E-02 |
| *HERPUD2* | 3.55 | 2.05 | 4.14E-04 | 3.91E-02 |
| *TAF11* | 3.51 | 1.51 | 4.72E-04 | 4.26E-02 |
| *GSK3B* | 3.47 | 1.85 | 5.40E-04 | 4.68E-02 |

**Supplemental Table S13. The significantly down-regulated DEGs among *ORMDL3^+^* cholangiocytes**

| **Gene** | **T score** | **Fold change** | **P value** | **FDR** |
| --- | --- | --- | --- | --- |
| *LDHB* | -7.08 | 0.20 | 1.76E-12 | 1.48E-08 |
| *CLDN10* | -6.94 | 0.05 | 5.00E-12 | 2.82E-08 |
| *TACSTD2* | -6.81 | 0.08 | 1.23E-11 | 5.19E-08 |
| *S100A6* | -6.15 | 0.30 | 9.39E-10 | 3.17E-06 |
| *SPP1* | -5.99 | 0.20 | 2.39E-09 | 6.21E-06 |
| *FXYD2* | -5.98 | 0.21 | 2.57E-09 | 6.21E-06 |
| *KRT19* | -5.88 | 0.13 | 4.50E-09 | 9.51E-06 |
| *GSTP1* | -5.81 | 0.30 | 7.08E-09 | 1.33E-05 |
| *PKM* | -5.73 | 0.05 | 1.11E-08 | 1.87E-05 |
| *PLPP2* | -5.62 | 0.07 | 2.12E-08 | 2.99E-05 |
| *ARHGDIB* | -5.46 | 0.15 | 5.13E-08 | 5.95E-05 |
| *CYBA* | -5.47 | 0.29 | 4.89E-08 | 5.95E-05 |
| *S100A11* | -5.34 | 0.28 | 1.03E-07 | 1.05E-04 |
| *SPINT2* | -5.33 | 0.21 | 1.06E-07 | 1.05E-04 |
| *CD24* | -5.23 | 0.30 | 1.92E-07 | 1.71E-04 |
| *LGALS3* | -5.18 | 0.19 | 2.34E-07 | 1.98E-04 |
| *COTL1* | -5.05 | 0.10 | 4.60E-07 | 3.38E-04 |
| *CLDN4* | -4.97 | 0.12 | 7.18E-07 | 4.67E-04 |
| *GYPC* | -4.91 | 0.17 | 9.85E-07 | 5.94E-04 |
| *LGALS2* | -4.84 | 0.14 | 1.34E-06 | 7.57E-04 |
| *HLA-DRA* | -4.77 | 0.23 | 1.99E-06 | 1.08E-03 |
| *C12orf75* | -4.76 | 0.21 | 2.07E-06 | 1.09E-03 |
| *FXYD5* | -4.69 | 0.13 | 2.91E-06 | 1.44E-03 |
| *CD74* | -4.62 | 0.52 | 4.16E-06 | 1.90E-03 |
| *RP11-386I14.4* | -4.56 | 0.08 | 5.41E-06 | 2.20E-03 |
| *HEXB* | -4.56 | 0.60 | 5.60E-06 | 2.20E-03 |
| *LCN2* | -4.57 | 0.37 | 5.09E-06 | 2.20E-03 |
| *RPS4Y1* | -4.56 | 0.28 | 5.44E-06 | 2.20E-03 |
| *PXYLP1* | -4.54 | 0.10 | 5.99E-06 | 2.30E-03 |
| *SMIM22* | -4.33 | 0.11 | 1.55E-05 | 4.44E-03 |
| *CDK7* | -4.30 | 0.19 | 1.78E-05 | 4.93E-03 |
| *ANXA2* | -4.26 | 0.60 | 2.20E-05 | 5.74E-03 |
| *CXCL8* | -4.23 | 0.12 | 2.43E-05 | 6.21E-03 |
| *SOD3* | -4.21 | 0.08 | 2.65E-05 | 6.58E-03 |
| *CCL3* | -4.20 | 0.08 | 2.77E-05 | 6.68E-03 |
| *TIMP2* | -4.19 | 0.18 | 2.85E-05 | 6.68E-03 |
| *LYZ* | -4.18 | 0.26 | 2.97E-05 | 6.87E-03 |
| *PDZK1IP1* | -4.18 | 0.23 | 3.03E-05 | 6.87E-03 |
| *CXCL1* | -4.15 | 0.19 | 3.35E-05 | 7.35E-03 |
| *EPCAM* | -4.15 | 0.20 | 3.47E-05 | 7.52E-03 |
| *VIM* | -4.12 | 0.30 | 3.88E-05 | 8.18E-03 |
| *KRT7* | -4.13 | 0.29 | 3.87E-05 | 8.18E-03 |
| *BAX* | -4.10 | 0.51 | 4.33E-05 | 9.03E-03 |
| *STK17B* | -4.07 | 0.08 | 4.84E-05 | 9.86E-03 |
| *PLAC8* | -4.07 | 0.08 | 4.84E-05 | 9.86E-03 |
| *PCAT19* | -4.05 | 0.14 | 5.29E-05 | 1.05E-02 |
| *TESC* | -3.89 | 0.24 | 1.01E-04 | 1.64E-02 |
| *TIMP1* | -3.88 | 0.59 | 1.08E-04 | 1.72E-02 |
| *PIGR* | -3.87 | 0.54 | 1.14E-04 | 1.79E-02 |
| *ANXA5* | -3.85 | 0.63 | 1.21E-04 | 1.84E-02 |
| *IGLC2* | -3.85 | 0.24 | 1.19E-04 | 1.84E-02 |
| *C19orf33* | -3.84 | 0.16 | 1.26E-04 | 1.86E-02 |
| *DAB2* | -3.82 | 0.24 | 1.35E-04 | 1.91E-02 |
| *TFF3* | -3.77 | 0.18 | 1.66E-04 | 2.19E-02 |
| *SFRP5* | -3.75 | 0.25 | 1.84E-04 | 2.30E-02 |
| *IFI16* | -3.74 | 0.22 | 1.91E-04 | 2.32E-02 |
| *FXYD3* | -3.74 | 0.12 | 1.89E-04 | 2.32E-02 |
| *PLLP* | -3.70 | 0.29 | 2.19E-04 | 2.55E-02 |
| *SLC5A1* | -3.65 | 0.19 | 2.69E-04 | 3.03E-02 |
| *SCGB3A1* | -3.60 | 0.15 | 3.20E-04 | 3.48E-02 |
| *SERPINB1* | -3.59 | 0.49 | 3.41E-04 | 3.54E-02 |
| *SRGN* | -3.58 | 0.38 | 3.48E-04 | 3.56E-02 |
| *VCAN* | -3.57 | 0.13 | 3.57E-04 | 3.59E-02 |
| *TPM4* | -3.56 | 0.32 | 3.76E-04 | 3.66E-02 |
| *GNG11* | -3.56 | 0.24 | 3.80E-04 | 3.67E-02 |
| *CHST4* | -3.55 | 0.25 | 3.92E-04 | 3.74E-02 |
| *FCN3* | -3.54 | 0.25 | 4.06E-04 | 3.85E-02 |
| *KLF4* | -3.53 | 0.19 | 4.18E-04 | 3.92E-02 |
| *IRF1* | -3.51 | 0.43 | 4.69E-04 | 4.26E-02 |
| *CTSS* | -3.48 | 0.52 | 5.10E-04 | 4.51E-02 |
| *C1QC* | -3.46 | 0.09 | 5.44E-04 | 4.69E-02 |

**Supplemental Table S14. Significantly KEGG pathways enriched by down-regulated DEGs among *ORMDL3^+^* cholangiocytes using the clusterProfiler tool**

| **ID** | **Description** | **Enrichment ratio** | **P value** | **FDR** |
| --- | --- | --- | --- | --- |
| hsa04640 | Hematopoietic cell lineage | 0.04 | 3.32E-10 | 1.07E-07 |
| hsa05140 | Leishmaniasis | 0.03 | 7.85E-10 | 1.27E-07 |
| hsa04061 | Viral protein interaction with cytokine and cytokine receptor | 0.03 | 1.90E-09 | 2.04E-07 |
| hsa05323 | Rheumatoid arthritis | 0.03 | 3.08E-09 | 2.49E-07 |
| hsa04514 | Cell adhesion molecules | 0.04 | 5.33E-08 | 3.44E-06 |
| hsa05144 | Malaria | 0.02 | 1.04E-07 | 5.61E-06 |
| hsa04060 | Cytokine-cytokine receptor interaction | 0.07 | 1.37E-07 | 6.31E-06 |
| hsa05321 | Inflammatory bowel disease | 0.02 | 3.42E-07 | 1.15E-05 |
| hsa04672 | Intestinal immune network for IgA production | 0.02 | 3.51E-07 | 1.15E-05 |
| hsa05340 | Primary immunodeficiency | 0.02 | 3.91E-07 | 1.15E-05 |
| hsa05150 | Staphylococcus aureus infection | 0.03 | 3.91E-07 | 1.15E-05 |
| hsa04062 | Chemokine signaling pathway | 0.05 | 6.47E-07 | 1.74E-05 |
| hsa05152 | Tuberculosis | 0.04 | 1.22E-06 | 3.03E-05 |
| hsa04658 | Th1 and Th2 cell differentiation | 0.03 | 1.52E-06 | 3.51E-05 |
| hsa05168 | Herpes simplex virus 1 infection | 0.09 | 2.03E-06 | 4.38E-05 |
| hsa05332 | Graft-versus-host disease | 0.02 | 2.42E-06 | 4.89E-05 |
| hsa05310 | Asthma | 0.01 | 2.64E-06 | 5.01E-05 |
| hsa04659 | Th17 cell differentiation | 0.03 | 5.63E-06 | 1.01E-04 |
| hsa04612 | Antigen processing and presentation | 0.02 | 1.58E-05 | 2.68E-04 |
| hsa04940 | Type I diabetes mellitus | 0.02 | 1.70E-05 | 2.74E-04 |
| hsa04512 | ECM-receptor interaction | 0.02 | 1.79E-05 | 2.76E-04 |
| hsa04145 | Phagosome | 0.04 | 3.16E-05 | 4.64E-04 |
| hsa04668 | TNF signaling pathway | 0.03 | 4.29E-05 | 5.77E-04 |
| hsa05145 | Toxoplasmosis | 0.03 | 4.29E-05 | 5.77E-04 |
| hsa05330 | Allograft rejection | 0.01 | 5.43E-05 | 7.02E-04 |
| hsa04670 | Leukocyte transendothelial migration | 0.03 | 6.20E-05 | 7.71E-04 |
| hsa05142 | Chagas disease | 0.03 | 1.16E-04 | 1.34E-03 |
| hsa05146 | Amoebiasis | 0.03 | 1.16E-04 | 1.34E-03 |
| hsa04510 | Focal adhesion | 0.04 | 1.37E-04 | 1.52E-03 |
| hsa05133 | Pertussis | 0.02 | 2.69E-04 | 2.90E-03 |
| hsa04064 | NF-kappa B signaling pathway | 0.02 | 4.11E-04 | 4.15E-03 |
| hsa04660 | T cell receptor signaling pathway | 0.02 | 4.11E-04 | 4.15E-03 |
| hsa04933 | AGE-RAGE signaling pathway in diabetic complications | 0.02 | 5.10E-04 | 4.99E-03 |
| hsa05410 | Hypertrophic cardiomyopathy | 0.02 | 5.56E-04 | 5.28E-03 |
| hsa04380 | Osteoclast differentiation | 0.03 | 5.93E-04 | 5.47E-03 |
| hsa04650 | Natural killer cell mediated cytotoxicity | 0.03 | 9.00E-04 | 8.08E-03 |
| hsa05166 | Human T-cell leukemia virus 1 infection | 0.04 | 1.03E-03 | 8.98E-03 |
| hsa05414 | Dilated cardiomyopathy | 0.02 | 1.49E-03 | 1.27E-02 |
| hsa05416 | Viral myocarditis | 0.02 | 1.69E-03 | 1.40E-02 |
| hsa05412 | Arrhythmogenic right ventricular cardiomyopathy | 0.02 | 2.20E-03 | 1.78E-02 |
| hsa04110 | Cell cycle | 0.03 | 3.25E-03 | 2.56E-02 |
| hsa04071 | Sphingolipid signaling pathway | 0.02 | 3.55E-03 | 2.73E-02 |
| hsa04151 | PI3K-Akt signaling pathway | 0.06 | 4.16E-03 | 3.13E-02 |
| hsa04015 | Rap1 signaling pathway | 0.04 | 4.32E-03 | 3.17E-02 |
| hsa04625 | C-type lectin receptor signaling pathway | 0.02 | 4.61E-03 | 3.31E-02 |
| hsa05418 | Fluid shear stress and atherosclerosis | 0.03 | 4.81E-03 | 3.38E-02 |
| hsa04657 | IL-17 signaling pathway | 0.02 | 5.47E-03 | 3.72E-02 |
| hsa04024 | cAMP signaling pathway | 0.04 | 5.53E-03 | 3.72E-02 |
| hsa04928 | Parathyroid hormone synthesis, secretion and action | 0.02 | 5.94E-03 | 3.92E-02 |
| hsa05169 | Epstein-Barr virus infection | 0.04 | 6.28E-03 | 4.06E-02 |
| hsa04020 | Calcium signaling pathway | 0.04 | 6.68E-03 | 4.23E-02 |
| hsa04973 | Carbohydrate digestion and absorption | 0.01 | 7.12E-03 | 4.43E-02 |
| hsa05202 | Transcriptional misregulation in cancer | 0.04 | 7.67E-03 | 4.68E-02 |
| hsa05320 | Autoimmune thyroid disease | 0.01 | 8.25E-03 | 4.93E-02 |
| hsa04810 | Regulation of actin cytoskeleton | 0.04 | 8.47E-03 | 4.98E-02 |

**Supplemental Table S15. Significantly KEGG pathways enriched by up-regulated DEGs among *ORMDL3^+^* cholangiocytes using the clusterProfiler tool.**

| **ID** | **Description** | **Enrichment ratio** | **P value** | **FDR** |
| --- | --- | --- | --- | --- |
| hsa05168 | Herpes simplex virus 1 infection | 0.17 | 4.32E-17 | 1.32E-14 |
| hsa05224 | Breast cancer | 0.04 | 3.13E-04 | 3.24E-02 |
| hsa05226 | Gastric cancer | 0.04 | 3.75E-04 | 3.24E-02 |
| hsa04550 | Signaling pathways regulating pluripotency of stem cells | 0.04 | 5.94E-04 | 3.24E-02 |
| hsa04310 | Wnt signaling pathway | 0.04 | 6.07E-04 | 3.24E-02 |
| hsa00140 | Steroid hormone biosynthesis | 0.02 | 6.80E-04 | 3.24E-02 |
| hsa04390 | Hippo signaling pathway | 0.04 | 7.44E-04 | 3.24E-02 |

**Supplemental Table S16. GO enrichment analysis according to biological process terms for 71 down-regulated DEGs among *ORMDL3^+^* cholangiocytes**

| **Gene Set** | **Description** | **Ratio** | **P Value** | **FDR** |
| --- | --- | --- | --- | --- |
| GO:0036230 | Granulocyte Activation | 8.60 | 1.12E-12 | 9.54E-10 |
| GO:0002446 | Neutrophil Mediated Immunity | 7.71 | 1.30E-10 | 5.53E-08 |
| GO:0007162 | Negative Regulation Of Cell Adhesion | 8.36 | 1.13E-06 | 3.20E-04 |
| GO:0010959 | Regulation Of Metal Ion Transport | 5.97 | 1.75E-05 | 3.46E-03 |
| GO:0043900 | Regulation Of Multi-Organism Process | 5.86 | 2.03E-05 | 3.46E-03 |
| GO:0048144 | Fibroblast Proliferation | 14.22 | 2.61E-05 | 3.70E-03 |
| GO:0022407 | Regulation Of Cell-Cell Adhesion | 4.99 | 1.86E-04 | 1.95E-02 |
| GO:0097066 | Response To Thyroid Hormone | 26.54 | 1.90E-04 | 1.95E-02 |
| GO:0002521 | Leukocyte Differentiation | 4.33 | 2.06E-04 | 1.95E-02 |
| GO:0051235 | Maintenance Of Location | 5.41 | 2.96E-04 | 2.50E-02 |
| GO:0071216 | Cellular Response To Biotic Stimulus | 6.46 | 3.24E-04 | 2.50E-02 |
| GO:0002237 | Response To Molecule Of Bacterial Origin | 5.07 | 4.40E-04 | 3.12E-02 |
| GO:0006959 | Humoral Immune Response | 5.92 | 5.12E-04 | 3.35E-02 |
| GO:0043270 | Positive Regulation Of Ion Transport | 5.55 | 7.17E-04 | 4.03E-02 |
| GO:0048871 | Multicellular Organismal Homeostasis | 4.07 | 7.28E-04 | 4.03E-02 |
| GO:0098542 | Defense Response To Other Organism | 4.04 | 7.59E-04 | 4.03E-02 |
| GO:0002764 | Immune Response-Regulating Signaling Pathway | 3.94 | 8.93E-04 | 4.47E-02 |
| GO:0030099 | Myeloid Cell Differentiation | 4.41 | 9.97E-04 | 4.71E-02 |
| GO:0070661 | Leukocyte Proliferation | 5.10 | 1.12E-03 | 4.99E-02 |

**Supplemental Table S17. GO enrichment analysis according to cellular component terms for 71 down-regulated DEGs among *ORMDL3^+^* cholangiocytes**

| **Gene Set** | **Description** | **Ratio** | **P Value** | **FDR** |
| --- | --- | --- | --- | --- |
| GO:0031983 | Vesicle Lumen | 8.21 | 7.04E-10 | 1.21E-07 |
| GO:0031012 | Extracellular Matrix | 4.78 | 4.83E-06 | 4.16E-04 |
| GO:0005775 | Vacuolar Lumen | 8.14 | 2.13E-05 | 1.22E-03 |
| GO:0070820 | Tertiary Granule | 7.27 | 1.61E-04 | 6.92E-03 |
| GO:0101002 | Ficolin-1-Rich Granule | 6.48 | 3.02E-04 | 1.04E-02 |
| GO:0005766 | Primary Lysosome | 6.38 | 1.07E-03 | 3.03E-02 |
| GO:0042581 | Specific Granule | 6.18 | 1.23E-03 | 3.03E-02 |

**Supplemental Table S18. GO enrichment analysis according to molecular function terms for 71 down-regulated DEGs among *ORMDL3^+^* cholangiocytes**

| **Gene Set** | **Description** | **Ratio** | **P Value** | **FDR** |
| --- | --- | --- | --- | --- |
| GO:0098641 | Cadherin Binding Involved In Cell-Cell Adhesion | 57.54 | 6.03E-07 | 1.06E-03 |
| GO:0098772 | Molecular Function Regulator | 3.11 | 1.13E-06 | 1.06E-03 |
| GO:0030234 | Enzyme Regulator Activity | 4.03 | 2.74E-06 | 1.71E-03 |
| GO:0044548 | S100 Protein Binding | 58.56 | 1.65E-05 | 7.72E-03 |
| GO:0023026 | MHC Class II Protein Complex Binding | 54.66 | 2.06E-05 | 7.72E-03 |
| GO:0098632 | Cell-Cell Adhesion Mediator Activity | 21.86 | 3.29E-05 | 1.01E-02 |
| GO:0023023 | MHC Protein Complex Binding | 43.15 | 4.33E-05 | 1.01E-02 |
| GO:0050840 | Extracellular Matrix Binding | 19.88 | 4.81E-05 | 1.01E-02 |
| GO:0004857 | Enzyme Inhibitor Activity | 6.06 | 4.84E-05 | 1.01E-02 |
| GO:0098631 | Cell Adhesion Mediator Activity | 18.53 | 6.35E-05 | 1.19E-02 |
| GO:0005125 | Cytokine Activity | 7.56 | 1.39E-04 | 2.37E-02 |
| GO:0017080 | Sodium Channel Regulator Activity | 23.43 | 2.81E-04 | 4.22E-02 |
| GO:0042802 | Identical Protein Binding | 2.58 | 2.93E-04 | 4.22E-02 |

**Supplemental Table S19. Disease-term enrichment analysis for 71 significantly down-regulated genes among *ORMLD3^+^* cholangiocytes based on the Disgenet database.** Note: There were six significant disease-terms overrepresented (FDR<0.05).

| **Gene Set** | **Description** | **Ratio** | **P Value** | **FDR** |
| --- | --- | --- | --- | --- |
| C0023893 | Liver Cirrhosis, Experimental | 5.72 | 4.00E-15 | 1.47E-11 |
| C4277682 | Chemical and Drug Induced Liver Injury | 7.66 | 5.28E-07 | 9.69E-04 |
| C0948089 | Acute Coronary Syndrome | 24.61 | 1.53E-06 | 1.87E-03 |
| C2609414 | Acute kidney injury | 13.54 | 4.71E-06 | 4.33E-03 |
| C0027626 | Neoplasm Invasiveness | 7.95 | 2.48E-05 | 1.82E-02 |
| C0024117 | Chronic Obstructive Airway Disease | 17.10 | 8.13E-05 | 4.98E-02 |
| C0034069 | Pulmonary Fibrosis | 10.03 | 1.32E-04 | 6.93E-02 |
| C0006663 | Calcinosis | 14.44 | 1.59E-04 | 7.30E-02 |
| C1458155 | Mammary Neoplasms | 3.63 | 3.48E-04 | 1.42E-01 |
| C0019202 | Hepatolenticular Degeneration | 20.30 | 4.08E-04 | 1.50E-01 |
| C0011853 | Diabetes Mellitus, Experimental | 7.59 | 4.85E-04 | 1.62E-01 |
| C0345967 | Malignant mesothelioma | 7.45 | 5.28E-04 | 1.62E-01 |
| C0022658 | Kidney Diseases | 7.00 | 7.02E-04 | 1.98E-01 |
| C0022650 | Kidney Calculi | 46.41 | 7.66E-04 | 2.01E-01 |
| C0011616 | Contact Dermatitis | 9.15 | 9.21E-04 | 2.25E-01 |
| C0018824 | Heart valve disease | 13.54 | 1.37E-03 | 3.14E-01 |
| C0008311 | Cholangitis | 32.48 | 1.62E-03 | 3.48E-01 |
| C0024667 | Animal Mammary Neoplasms | 5.68 | 1.79E-03 | 3.48E-01 |
| C0035126 | Reperfusion Injury | 7.64 | 1.80E-03 | 3.48E-01 |
| C1876165 | Copper-Overload Cirrhosis | 29.53 | 1.98E-03 | 3.63E-01 |
| C0018799 | Heart Diseases | 11.33 | 2.29E-03 | 3.94E-01 |
| C0003949 | Asbestosis | 27.07 | 2.36E-03 | 3.94E-01 |
| C0024668 | Mammary Neoplasms, Experimental | 5.24 | 2.55E-03 | 4.07E-01 |
| C0043094 | Weight Gain | 6.50 | 3.26E-03 | 4.99E-01 |
| C0027540 | Necrosis | 9.19 | 4.16E-03 | 6.12E-01 |
| C0021368 | Inflammation | 5.70 | 5.20E-03 | 7.04E-01 |
| C0027627 | Neoplasm Metastasis | 4.41 | 5.31E-03 | 7.04E-01 |
| C0007621 | Neoplastic Cell Transformation | 5.65 | 5.36E-03 | 7.04E-01 |
| C0020517 | Hypersensitivity | 7.73 | 6.76E-03 | 8.56E-01 |
